# Distinguishing causal from tagging enhancers using single-cell multiome data

**DOI:** 10.64898/2026.02.15.26346353

**Authors:** Elizabeth Dorans, Alkes L. Price

## Abstract

Methods that analyze single-cell RNA-seq+ATAC-seq multiome data have shown promise in linking enhancers to target genes by correlating chromatin accessibility with gene expression across cells. However, correlations among ATAC-seq peaks may induce non-causal *tagging* peak-gene links (analogous to tagging associations in GWAS); indeed, we confirm that tagging effects induced by peak co-accessibility are pervasive in peak-gene linking. We defined two scores for each ATAC-seq peak: *co-accessibility score*, the sum of squared correlations with each nearby peak; and *co-activity score*, the sum of squared correlations with each nearby gene. We compared these scores in 4 multiome data sets (spanning 86k cells and 6 immune/blood cell types) and determined that co-accessibility score and co-activity score were strongly correlated across peaks (*r* = 0.57-0.73); these correlations were not explained by read depth, cell subtypes, or measurement noise, but are consistent with tagging. Indeed, non-causal peak-gene correlations were strongly correlated to a peak’s tagging correlation with a causal peak in CRISPRi data (*r* = 0.92). We further determined that causal peak-gene associations are concentrated in specific functional categories of peaks, by regressing co-activity scores on stratified co-accessibility scores (S-CASC): e.g. 2.91x (s.e. 0.67) enrichment for peaks closest to a gene’s TSS and 1.41x (s.e. 0.11) enrichment for peaks overlapping H3K27ac marks. Co-accessibility scores were substantially driven by the number of transcription factor binding sites (TFBS) within a peak, and peak-peak correlations were substantially driven by the number of TFBS pairs within the two peaks with a shared TF. These effects were concentrated in a small number of *pioneer* TFs, which activate repressed chromatin regions. Consistent with widespread tagging, peak-gene links that we fine-mapped using SuSiE significantly outperformed marginal peak-gene links in evaluation sets derived from CRISPRi and eQTL data. We provide examples demonstrating the impact of tagging effects at specific peaks and genes implicated in GWAS of blood cell traits. Our findings underscore the importance of accounting for tagging effects when linking enhancers to target genes.

## Introduction

Mapping gene regulatory architectures is critical for the functional interpretation of GWAS discoveries and understanding the circuitry driving key developmental and disease processes^1–10^. ‘Peak-gene linking methods,’ which predict regulatory relationships from single-cell RNA-seq+ATAC-seq multiome data by correlating the accessibility of peaks of open chromatin in ATAC-seq data with genes in RNA-seq data, have shown particular promise in linking regulatory elements to their target genes^11–24^. Previously proposed peak-gene linking methods have attempted to account for false positive links induced by technical and biological confounders^12,13,15,19,20^ and gene co-expression^17^. Other studies have produced methods for mapping peak co-accessibility^11,14,25^ or inferred ‘modules’ of co-accessible peaks^16,26^ from single-cell ATAC-seq data, raising the question of how peak co-accessibility might impact peak-gene linking.

We hypothesized that correlations among ATAC-seq peaks may induce non-causal *tagging* peak-gene links, analogous to tagging associations in GWAS (**Figure 1**); in particular, we hypothesized that these correlations are induced by the binding of shared transcription factors (TFs) at multiple ATAC peaks, with patterns of co-accessibility reflecting dynamic cell types / states. Here, we show that tagging effects, induced by peak co-accessibility, are pervasive in peak-gene linking. We first define two scores for each ATAC-seq peak: *co-accessibility score*, quantifying peak-peak correlations, and *co-activity score*, quantifying peak-gene correlations. We investigate peak tagging by analyzing CRISPR data and by regressing co-activity scores on stratified co-accessibility scores (S-CASC) to disentangle causal from tagging effects and pinpoint categories of peaks harboring causal peak-gene links. We then assess TF binding activity as a potential biological mechanism underlying peak co-accessibility. We further investigate tagging peak-gene associations induced by gene-gene co-expression. Finally, we explore statistical fine-mapping^27^ as a method to distinguish causal from tagging peak-gene links, and we highlight several examples. Understanding the correlation structure among peaks and genes in single-cell multiome data is critical for accurate mapping of regulatory links and functional interpretation of GWAS discoveries.

**Figure 1.**
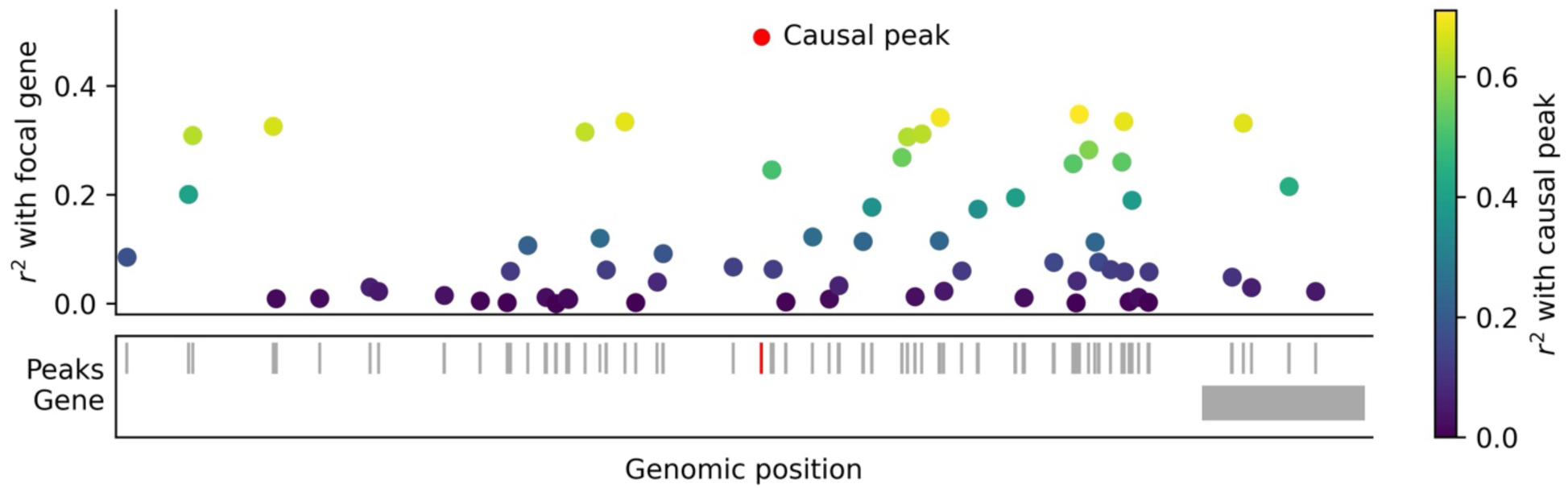
Toy example illustrating the effects of peak tagging on peak-gene links. We plot simulated squared peak-gene correlations for each peak <1Mb from the TSS of the focal gene. *r*^2^ values are reported as dots in the middle of each peak, with the causal peak colored in red and other peaks colored based on their squared correlation to the causal peak.

## Results

### Overview of methods

To quantify peak-peak and peak-gene correlations for each peak in single-cell multiome data, we define two scores for each peak: *co-accessibility score* (quantifying peak-peak correlations) and *co-activity score* (quantifying peak-gene correlations), each reflecting a sum of squared correlations. We assess the relationship between co-accessibility scores and co-activity scores across peaks. We propose a method, stratified co-accessibility score regression (S-CASC), to identify functional peak categories that are enriched for causal peak-gene associations. We note that co-accessibility scores can be viewed as an analogue of SNP LD scores^28^, co-activity scores can be viewed as an analogue of GWAS associations, and S-CASC can be viewed as an analogue of stratified LD score regression^4^.

In detail, we define the *co-accessibility score* of peak 𝑗 as 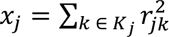, where 𝐾_j_ is the set of all *cis* peaks located <1Mb from focal peak 𝑗 (including peak 𝑗 itself) and 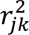 are squared peak-peak correlations (from scATAC-seq data) across metacells^14^. We define the *co-activity score* of peak 𝑗 as 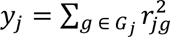, where 𝐺_j_ is the set of all *cis* genes with TSS <1Mb from focal peak 𝑗 and 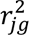 are squared peak-gene correlations (from scATAC-seq and scRNA-seq data) across metacells. In all analyses, we exclude peaks with distance <1kb to any TSS^22,29^ (thus focusing primarily on enhancers rather than promoters), and we correct each 𝑟^2^ term for upward bias^28^ (**Methods**). We assess whether peaks with higher co-accessibility scores tend to have higher co-activity scores, as would be expected if peak-gene correlations arise in part due to peak-peak tagging effects.

S-CASC determines that a category of peaks is enriched for causal peak-gene associations if peaks with high co-accessibility to that category have higher co-activity scores than peaks with low co-accessibility to that category. The S-CASC regression relies on the following model:

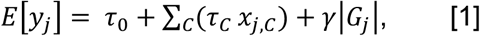

where 𝑦 is the co-activity score of peak 𝑗, 𝜏_0_ is an intercept term, 𝑥_j,𝐶_ is the co-accessibility score of peak 𝑗 with respect to peak category 𝐶 (defined as 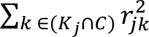), 𝜏_𝐶_ is the per-peak contribution to causal peak-gene effect variance for peak category 𝐶, and the covariate 2𝐺_j_2 is the number of genes with TSS <1Mb from peak 𝑗. We define the enrichment of a peak category as the proportion of causal peak-gene effect size variance from peaks in that category divided by the proportion of peaks in that category. We estimate standard errors by jackknifing across chromosomes; we then meta-analyze enrichments for each peak category across data set-cell type pairs (**Methods**). Due to peak measurement noise, we focus on the enrichment of a peak category, and not on total causal peak-gene effect size variance. We have released open-source software implementing S-CASC (**Code Availability**).

Although our primary focus is on peak-peak tagging effects, we also explore gene-gene tagging effects by defining the *gene co-expression score* of gene 𝑖 as 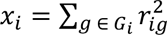, where 𝐺_𝑖_ is the set of all genes with TSS <1Mb from the TSS of focal gene *g* (including gene *g* itself) and 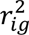 are squared gene-gene correlations across metacells; defining the *gene co-activity score* of gene 𝑖 as 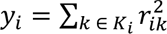, where 𝐾_𝑖_ is the set of all peaks located <1Mb from the TSS of focal gene 𝑖 and 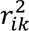 are squared gene-peak correlations across metacells; and identifying gene categories enriched for causal gene-peak associations using stratified co-expression score regression (**Methods** and **Code Availability**).

We analyzed 4 scRNA+ATAC-seq multiome data sets (Xu K562^30^, Satpathy K562^31^, SHARE-seq LCL^13^, and Luecken BMMC^32^) that each spanned 1-4 blood or immune cell types (total of 7 data set-cell type pairs, spanning 6 cell types and 86k cells after QC) (**Table 1**, **Supplementary Table 1** and **Methods**; see **Data Availability**). We have publicly released peak co-accessibility and co-activity scores and gene co-expression and co-activity scores for each data set-cell type pair analyzed in this study (**Data Availability**).

**Table 1.** Overview of single-cell multiome data sets. We report the cell types profiled and number of cells passing QC for each single-cell multiome data set. *: TF ChIP-seq data is available for that cell type. †: CRISPR data is available for that cell type. Further details are provided in **Supplementary Table 1**.

| Data set | Cell type(s) | Number of cells |
| --- | --- | --- |
| Xu<sup>30</sup> | K562*† | 7,861 |
| Satpathy<sup>31</sup> | K562*† | 5,359 |
| SHARE-seq LCL<sup>13</sup> | lymphoblastoid cell line (LCL)* | 17,644 |
| Luecken BMMC<sup>32</sup> | T, B, myeloid, erythroid | 55,321 |
| All | 6 cell types | 86,185 cells |

### Co-accessibility between peaks induces tagging peak-gene associations

We assessed the relationship between co-accessibility scores and co-activity scores across peaks, for each of the 7 data set-cell type pairs (**Table 1**). Results for the Xu-K562^30^ data set are reported in **Figure 2a** and **Supplementary Table 2**, and results for all other data set-cell type pairs are reported in **Supplementary Figure 1** and **Supplementary Table 2**. Co-accessibility scores and co-activity scores were strongly correlated (*r* = 0.57-0.73 in each data set-cell type pair), and these correlations remained strong after conditioning on the number of genes with TSS <1Mb from the focal peak (*r* = 0.48-0.69 in each data set-cell type pair). This result is potentially consistent with the hypothesis that co-accessibility between peaks induces tagging peak-gene associations (“tagging hypothesis”), but it is important to rule out other explanations (see below). Analyses using correlations across single cells instead of metacells produced similar findings (**Supplementary Figure 2**).

**Figure 2.**
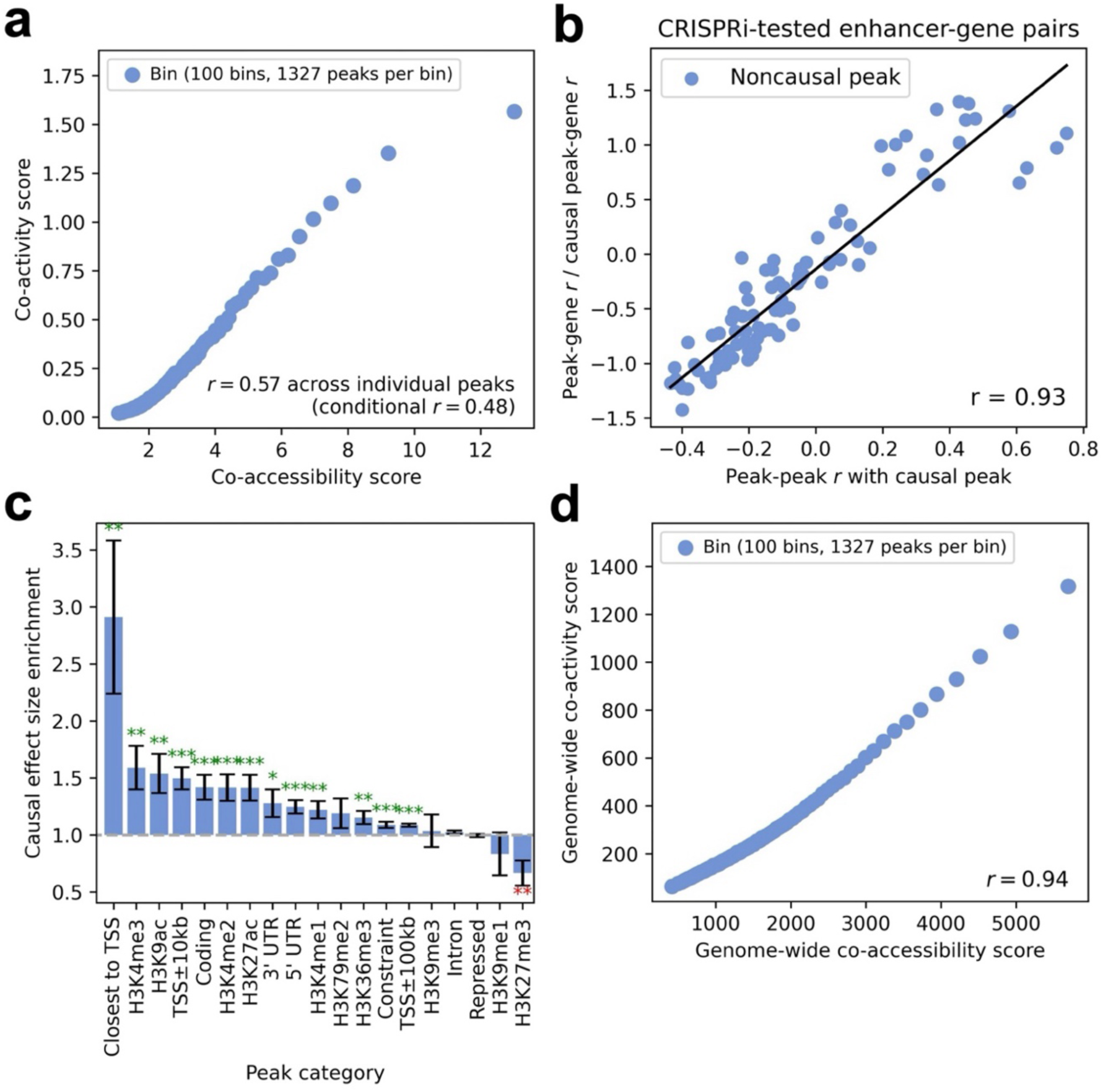
Relationship between peak-peak co-accessibility and peak-gene co-activity. **a)** Relationship between co-accessibility score and co-activity score across 132,736 peaks in the Xu K562^30^ data set, computed using peaks and genes within the *cis* window (<1Mb) of each focal peak. ‘Conditional *r*’ denotes correlation conditioned on number of genes <1Mb from the focal peak. **b)** Relationship between correlation with the CRISPR-validated causal peak and correlation with the target gene (relative to the causal peak’s correlation with the target gene) across non-causal peaks tested by CRISPRi, restricted to CRISPR positive-negative peaks where ArchR correctly identifies the causal peak-gene link. **c)** Causal effect size enrichment of 19 peak categories in stratified co-accessibility score regression. Bars and confidence intervals denote estimates and standard errors, respectively, meta-analyzed across data set-cell type pairs. Stars denote meta-analyzed p-values for significant enrichment (green) or depletion (red) (*: p < 0.05, **: p < 0.01, ***: p < 0.001). **d)** Relationship between genome-wide co-accessibility score and genome-wide co-activity score across 132,736 peaks in the Xu K562^30^ data set, computed using all peaks and genes for each focal peak. In **a** and **d,** peaks are partitioned equally into 100 bins (each represented by 1 point) by co-accessibility score and genome-wide co-accessibility score, respectively. Numerical results are reported in **Supplementary Table 2**, **Supplementary Table 4**, and **Supplementary Table 5**.

To evaluate the tagging hypothesis, we analyzed CRISPRi-tested peak-gene pairs^33–44^ in K562 cells, restricting to genes with both an experimentally validated causal (CRISPR-positive) peak that is identified by ArchR^14^ (with peak-gene correlation > 0.45) and at least one experimentally tested non-causal (CRISPR-negative) peak overlapping our single-cell data. We restricted this analysis to the 3 data set-cell type pairs most relevant to K562 cells: Xu-K562^30^, Satpathy-K562^31^, and Luecken-erythroid^32^ (**Table 1**). We measured the correlation (across metacells) of each CRISPR-negative peak with both the CRISPR-positive peak and the focal gene. Correlation with the CRISPR-positive peak was strongly correlated to correlation with the focal gene (*r* = 0.92; **Figure 2b**), corroborating the tagging hypothesis. Analyses using correlations across single cells instead of metacells produced similar findings (**Supplementary Figure 3**).

To further evaluate this hypothesis, we computed the enrichment of causal peak-gene effect size variance for 19 functional categories of peaks using S-CASC; the 19 functional categories of peaks included gene proximity, histone modifications, constraint, and other genic features (**Supplementary Table 3**). We observed significant enrichment of causal effect size variance for peak categories defined by gene proximity (closest to TSS, TSS±10kb, TSS±100kb), activating histone modifications (H3K4me3, H3K9ac, H3K4me2, H3K27ac, H3K4me1, H3K36me3), constraint, and other genic features (3’ UTR, 5’ UTR, coding) (ranging from 1.09±0.014 to 2.91±0.67), and significant depletion for the repressive histone modification H3K27me3 (0.67±0.11) (**Figure 2c**, **Supplementary Figure 4**, and **Supplementary Table 4**). These findings provide additional support for the tagging hypothesis, as alternative explanations for the correlation between co-accessibility scores and co-activity scores (see below) would not be expected to produce these functional enrichments. Analyses using correlations across single cells instead of metacells produced similar findings (**Supplementary Figure 5**). Analyses excluding the covariate for the number of genes <1Mb from each peak also produced similar findings. (**Supplementary Figure 6**).

We restricted most of our analyses to peaks/genes located <1Mb from the focal peak, but we also assessed the relationship (across focal peaks) between genome-wide co-accessibility scores and genome-wide co-activity scores, defined without this restriction (**Methods**). Genome-wide co-accessibility scores and genome-wide co-activity scores were very strongly correlated (**Figure 2d**, **Supplementary Figure 7** and **Supplementary Table 5**; *r* = 0.94-0.99 in each data set-cell type pair). Genome-wide co-accessibility scores were strongly correlated with co-accessibility scores restricted to <1Mb from focal peak (*r* = 0.73-0.86), and genome-wide co-activity scores were strongly correlated with co-activity scores restricted to <1Mb from focal peak (*r* = 0.58-0.67). Analyses using correlations across single cells instead of metacells produced similar findings (**Supplementary Figure 8**). This is consistent with peak tagging being driven by variation across cells in the abundance of particular transcription factors (TF) that bind at multiple sites across the genome (see below). This could involve the same copy of a TF binding to multiple nearby sites (*cis* TF effect), or different copies of the same TF binding to distal sites (*trans* TF effect), or a combination of *cis* and *trans* effects. The genome-wide correlation between co-accessibility and co-activity scores (**Figure 2d**) specifically implicates *trans* TF effects. On the other hand, our observation of higher peak-peak *r*^2^ at shorter genomic distances (**Supplementary Figure 9**) specifically implicates *cis* TF effects. We conclude that co-accessibility is driven by a combination of *cis* TF effects and *trans* TF effects.

We performed 5 secondary analyses to rule out alternative explanations for the correlation between co-accessibility scores and co-activity scores. First, to further assess the effects of biological causality, we performed a bivariate regression of co-activity score on co-accessibility score and a binary indicator of CRISPR-positive status (restricting to genes with CRISPR-positive and CRISPR-negative peaks), as well as univariate regressions of co-activity score on each predictor individually. We determined that co-accessibility score alone explained a much higher proportion of the observed variation in co-activity scores, and that the bivariate model did not substantially increase the proportion of variation explained (**Supplementary Figure 10**). Second, to assess peak read depth effects, we performed a bivariate regression of co-activity score on co-accessibility score and peak read depth, as well as univariate regressions of co-activity score on each predictor individually. We determined that co-accessibility score alone tended to explain a much higher proportion of the variance in co-activity scores (*r*^2^=0.34-0.58) vs. peak read depth alone (*r*^2^=0.0013-0.15), and that the bivariate model did not substantially increase the proportion of variation explained (*r*_2_=0.34-0.61; **Supplementary Figure 11**); we note that peak read depth explained *r*_2_=0.00046-0.12 of the variance in co-accessibility scores. Third, to assess cell subtype effects, we computed co-accessibility scores and co-activity scores using peak-peak and peak-gene correlations conditional on cell subtype proportions within each metacell of the Luecken^32^ T cell data set (for which major cell subtype annotations are available^32,45^). We determined that the correlation between co-accessibility scores and co-activity scores decreased but remained substantial (*r* = 0.51, vs. *r* = 0.62 for unconditioned correlation) (**Supplementary Figure 12**). This suggests that, although peak tagging is related to cell subtype / cell state-specific processes (see **Discussion**), major cell subtypes based on available annotations do not explain the bulk of the correlation between co-accessibility and co-activity. Fourth, to assess peak measurement noise effects, we introduced additional artificial noise for a subset of ATAC peaks and recomputed the correlation between co-accessibility scores and co-activity scores. We determined that adding noise did not substantially increase the correlation (**Supplementary Figure 13**). Fifth, we assessed the correlation between co-accessibility scores and co-activity scores when conditioning on the number of peaks located <1Mb from the focal peak (in addition to conditioning on the number of nearby genes; see above). We determined that the correlation remained strong (**Supplementary Figure 14**). We thus conclude that the correlation between co-accessibility scores and co-activity scores cannot be explained by biological causality (also see **Figure 2b**), peak read depth, major cell subtypes, varying peak measurement noise, or the number of nearby peaks.

We performed 3 additional secondary analyses. First, we estimated the proportion of gene expression variance explained by measured *cis-*peak accessibility using co-accessibility score regression (without stratifying by functional category). Our results suggested that the proportion of gene expression variance explained by measured *cis-*peak accessibility may exceed 0.5, but we caution that our estimates have limited interpretability (**Supplementary Figure 15**). Second, we assessed how much peak-peak correlation patterns vary across cell types by computing correlations across peaks between co-accessibility scores from each data set-cell type pair. We determined that co-accessibility scores are partially concordant across cell types (*r* = 0.08-0.35) and show stronger concordance in more similar cell types (**Supplementary Figure 16**), providing additional evidence that peak tagging is related to cell subtype / cell state-specific processes (see **Discussion**). Third, we recomputed standard errors and p-values for enrichments of peak categories using 200 jackknife blocks defined via *k*-means clustering on ATAC data, instead of via chromosomes. We determined that standard errors and p-values were similar using these two approaches (**Supplementary Table 6**).

These findings demonstrate the pervasive effects of peak-peak tagging on peak-gene associations.

### Co-accessibility between peaks is driven by transcription factor binding activity

We hypothesized that TF binding at multiple sites (including sites that do not impact gene expression in the focal cell type) could induce tagging correlations between pairs of ATAC-seq peaks—either via the same copy of a TF binding to multiple nearby sites (*cis* TF effect), or via different copies of the same TF binding to distal sites (*trans* TF effect), or via a combination of *cis* and *trans* effects—providing a mechanistic explanation for the relationship between peak tagging and cell subtypes / cell states (see **Discussion**). Under this hypothesis, *pioneer TFs*^46,47^ (a class of TFs with the ability to bind and activate closed chromatin) would be particularly powerful in driving tagging correlations. To evaluate the TF binding hypothesis, we analyzed transcription factor binding sites (TFBS) detected by ChIP-seq for 303 TFs in K562 cells and 148 TFs in GM12878 cells, including 9 known pioneer TFs in K562 and 8 known pioneer TFs in GM12878^46,47^ (**Supplementary Table 7** and **Methods**). For each ATAC-seq peak, we counted the number of TFs that bind within the peak. For each pair of ATAC-seq peaks, we counted the number of TFs that bind within both peaks. For each peak-gene pair, we counted the number of TFs that bind within both the peak and the promoter of the gene (defined as TSS±1kb^29^).

We computed the correlation across peaks between the number of TFs that bind within the peak and the co-accessibility score. Results for the Xu-K562^30^ data set are reported in **Figure 3a**, left panel and **Supplementary Table 8a**, and results for the Satpathy-K562^31^ and SHARE-seq-LCL^13^ data sets are reported in **Supplementary Figure 17a**, left panel, **Supplementary Figure 18a**, left panel, and **Supplementary Table 8a**. The number of TFs binding within a peak was moderately correlated to the co-accessibility score (*r* = 0.18-0.35 in each data set-cell type pair). To compare the effects of pioneer and non-pioneer TFs, we performed a bivariate linear regression of the co-accessibility score on the number of pioneer TFs that bind within a peak and the number of non-pioneer TFs that bind within a peak. We determined that the effect size per pioneer TFs was much larger than the effect size per non-pioneer TF (0.068±0.028 vs. 0.012±0.0047, p-value for difference = 0.028) (**Figure 3a**, right panel, **Supplementary Figure 17a**, right panel, **Supplementary Figure 18a**, right panel, **Supplementary Figure 19a**, right panel, and **Supplementary Table 8a**). These results are consistent with peak co-accessibility being driven by TF binding activity (particularly of pioneer TFs). Analyses using peak-peak correlations across single cells instead of metacells produced similar findings (**Supplementary Figure 20a**).

**Figure 3.**
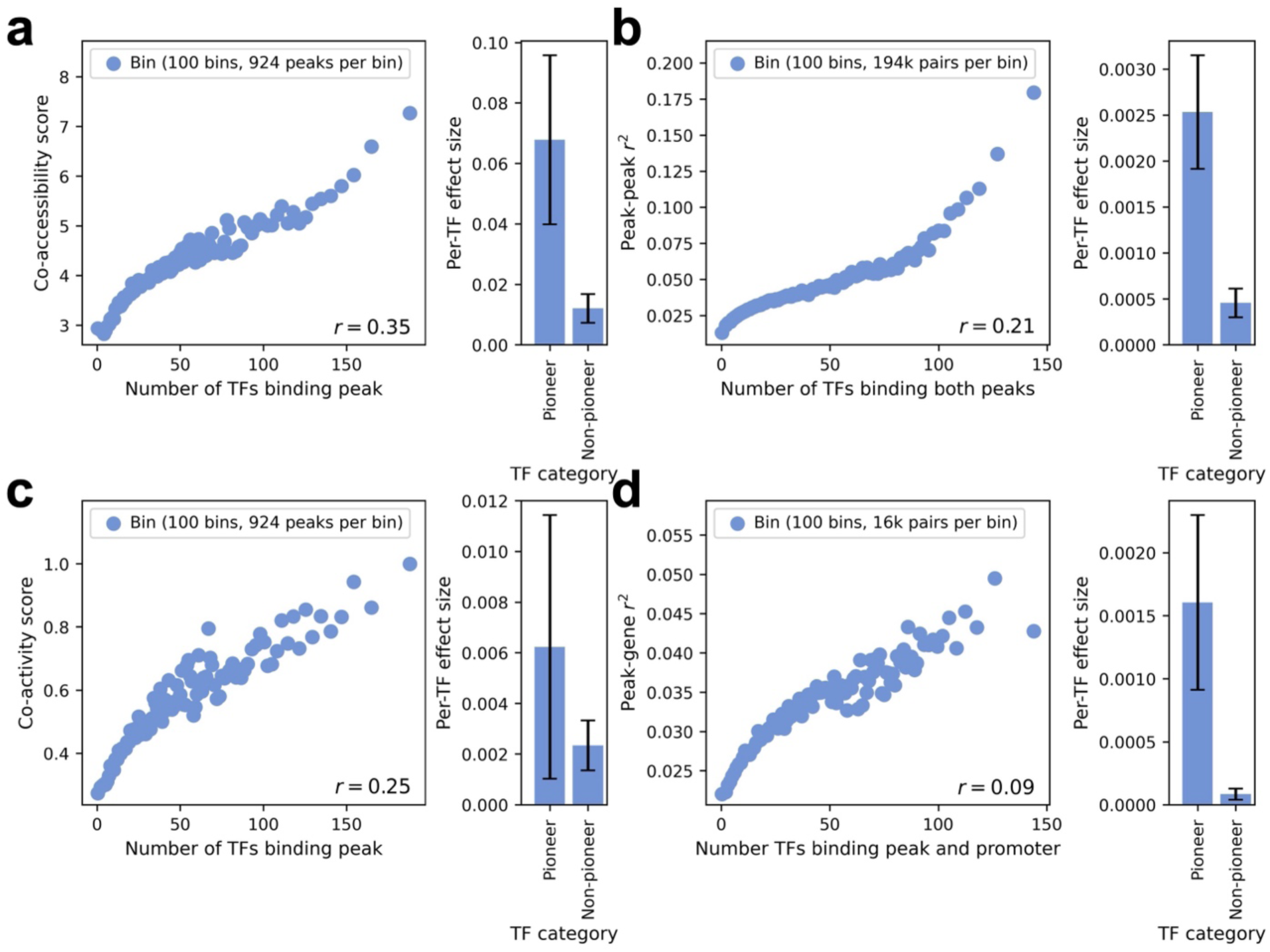
Peak co-accessibility and peak-gene co-activity are related to transcription factor binding activity. **a)** Relationship between number of TFs binding an ATAC peak and co-accessibility score (left). Per-TF effect for pioneer vs. non-pioneer TFs on co-accessibility score (right). **b)** Relationship between number of TFs binding both peaks in a peak-peak pair and squared peak-peak correlation (left). Per-TF effect for pioneer vs. non-pioneer TFs on squared peak-peak correlation (right). **c)** Relationship between number of TFs binding an ATAC peak and co-activity score (left). Per-TF effect for pioneer vs. non-pioneer TFs on co-activity score (right). **d)** Relationship between number of TFs binding both the peak and the gene (promoter) in a peak-gene pair and squared peak-gene correlation (left). Per-TF effect for pioneer vs. non-pioneer TFs on squared peak-gene correlation (right). In left subpanels of **a-d**, correlations are computed across 132,736 peaks in the Xu K562^30^ data set, and peaks (or peak-peak / peak-gene pairs) are partitioned equally into 100 bins (each represented by 1 point) by x-axis value. In right subpanels of **a-d**, regression effect sizes are meta-analyzed across 3 data set-cell type pairs (Xu K562^30^, Satpathy-K562^31^, and SHARE-seq-LCL^13^), and confidence intervals denote standard errors. Numerical results are reported in **Supplementary Table 8**.

We computed the correlation across peak-peak pairs between the number of TFs that bind within both peaks and the squared peak-peak correlation (across cells in scATAC-seq data). Results for the Xu-K562^30^ data set are reported in **Figure 3b**, left panel and **Supplementary Table 8b**, and results for the Satpathy-K562^31^ and SHARE-seq-LCL^13^ data sets are reported in **Supplementary Figure 17b**, left panel and **Supplementary Figure 18b**, left panel, and **Supplementary Table 8b**. The number of TFs that bind within both peaks was moderately correlated to the squared peak-peak correlation (*r* = 0.12-0.21 in each data set-cell type pair). To compare the effects of pioneer and non-pioneer TFs, we performed a bivariate linear regression of squared peak-peak correlation on the number of pioneer TFs and the number of non-pioneer TFs that bind within both peaks. We determined that the effect size per pioneer TF was much larger than the effect size per non-pioneer TF (0.0025±0.00062 vs. 0.00046±0.00016, p-value for difference = 2e-5) (**Figure 3b**, right panel, **Supplementary Figure 17b**, right panel, **Supplementary Figure 18b**, right panel, **Supplementary Figure 19b**, right panel, and **Supplementary Table 8b**). These results further corroborate the hypothesis that peak co-accessibility is driven by TF binding activity (particularly of pioneer TFs). Analyses using peak-peak correlations across single cells instead of metacells produced similar findings (**Supplementary Figure 20b**).

We computed the correlation across peaks between the number of TFs that bind within the peak and the co-activity score. Results for the Xu-K562^30^ data set are reported in **Figure 3c**, left panel and **Supplementary Table 8c**, and results for the Satpathy-K562^31^ and SHARE-seq LCL^13^ data sets are reported in **Supplementary Figure 17c**, left panel, **Supplementary Figure 18c**, left panel, and **Supplementary Table 8c**. The number of TFs binding within a peak was moderately correlated to the co-activity score (*r* = 0.14-0.25 in each data set-cell type pair). To compare the effects of pioneer and non-pioneer TFs, we performed a bivariate linear regression of co-activity score on the number of pioneer TFs that bind within a peak and the number of non-pioneer TFs that bind within a peak. We determined that the point estimate of the effect size per pioneer TF was larger than the point estimate of the effect size per non-pioneer TF, although the difference was not statistically significant due to noisy estimates (p-value for difference = 0.29) (**Figure 3c**, right panel, **Supplementary Figure 17c**, right panel, **Supplementary Figure 18c**, right panel, **Supplementary Figure 19c**, right panel, and **Supplementary Table 8c**). These results are consistent with peak-gene co-activity being impacted by TF-driven peak co-accessibility. Analyses using peak-gene correlations across single cells instead of metacells produced similar findings (**Supplementary Figure 20c**).

Finally, we computed the correlation across peak-gene pairs between the number of TFs that bind within both the peak and the promoter of the gene and the squared peak-gene correlation (across cells in scRNA-seq and ATAC-seq data). Results for the Xu-K562^30^ data set are reported in **Figure 3d**, left panel and **Supplementary Table 8d**, and results for the Satpathy-K562^31^ and SHARE-seq-LCL^13^ data sets are reported in **Supplementary Figure 17d**, left panel, **Supplementary Figure 17d**, left panel, and **Supplementary Table 8d**. The number of TFs binding within both the peak and the promoter of the gene was somewhat correlated to the squared peak-gene correlation (*r* = 0.03-0.09 in each data set-cell type pair). To compare the effects of pioneer and non-pioneer TFs, we performed a bivariate linear regression of squared peak-gene correlation on the number of pioneer TFs and the number of non-pioneer TFs that bind within both the peak and promoter of the gene. We determined that the effect size per pioneer TF was much larger than the effect size per non-pioneer TF (0.0016±0.00069 vs. 8.4e-5±4.4e-5, p-value for difference = 0.019) (**Figure 3d**, right panel, **Supplementary Figure 17d**, right panel, **Supplementary Figure 18d**, right panel, **Supplementary Figure 19d**, right panel, and **Supplementary Table 8d**). These results further corroborate the hypothesis that peak-gene co-activity is impacted by TF-driven peak co-accessibility. Analyses using peak-gene correlations across single cells instead of metacells produced similar findings (**Supplementary Figure 20d**).

We repeated the above analyses (**Figure 3a-d**) using genome-wide peak-peak and peak-gene pairs (and genome-wide co-accessibility and co-activity scores), instead of restricting to pairs with distance <1Mb. Correlations across peaks between the number of TFs that bind within a peak and either the co-accessibility score (**Figure 3a**) or the co-activity score (**Figure 3c**) became stronger when using genome-wide scores, with a larger and more significant difference for pioneer TFs vs. non-pioneer TFs (**Supplementary Figure 21a,c**). Correlations across peak-peak pairs (resp. peak-gene pairs) between the number of TFs that bind within both peaks (resp. the peak and the gene’s promoter) and the squared peak-peak correlation (**Figure 3b**) (resp. squared peak-gene correlation (**Figure 3d**)) were similar when using genome-wide pairs, with similar differences between pioneer TFs vs. non-pioneer TFs (**Supplementary Figure 21b,d**). These results further corroborate the *trans* TF binding mechanism of peak-peak tagging; as noted above, our observation of higher peak-peak *r*^2^ at shorter genomic distances (**Supplementary Figure 9**) also supports the *cis* TF binding mechanism.

We performed 3 secondary analyses. First, we repeated the bivariate regression of the co-accessibility score on the number of pioneer TFs that bind within a peak and the number of non-pioneer TFs that bind within a peak (**Figure 3a**, right panel) while adding covariates that bin TFs that bind each peak by their average binding and expression. We determined that the effect size per pioneer TF was still much larger than the effect size per non-pioneer TF (**Supplementary Figure 22**). Second, we repeated each bivariate regression comparing pioneer vs. non-pioneer TF effects (right panels of **Figure 3a-d**) using Perturb-multiome data to define pioneer TFs (TFs whose perturbation is associated with a significant change in peak accessibility or gene expression in scRNA+ATAC-seq multiome data^48^), and drew similar conclusions (**Supplementary Figure 23**). Third, we repeated the above analyses (**Figure 3a-d**) using sequence motif data^49^ instead of ChIP-seq data to quantify TF activity, and reached similar conclusions (**Supplementary Figures 24-28**). These analyses suggest that TF binding (particularly of pioneer TFs) at multiple sites may drive peak-peak tagging.

### Co-expression between genes induces tagging peak-gene associations

We shifted our focus to gene-gene tagging, a potential alternative source of non-causal peak-gene correlations, using gene co-expression scores and gene co-activity scores (defined above). We assessed the relationship between gene co-expression scores and gene co-activity scores across genes, for each of the 7 data set-cell type pairs (**Table 1**). Results for the Xu-K562^30^ data set are reported in **Figure 4a** and **Supplementary Table 9**, and results for all other data set-cell type pairs are reported in **Supplementary Figure 29** and **Supplementary Table 9**. Gene co-expression scores and gene co-activity scores were strongly correlated (*r* = 0.25-0.76 in each data set-cell type pair), and these correlations remained strong after conditioning on the number of peaks <1Mb from the focal gene (*r* = 0.26-0.65 in each data set-cell type pair). This result is potentially consistent with the hypothesis that co-expression between genes also induces tagging peak-gene associations (“gene tagging hypothesis”), in addition to tagging associations induced by co-accessibility between peaks. Analyses using correlations across single cells instead of metacells produced similar findings (**Supplementary Figure 30**).

**Figure 4.**
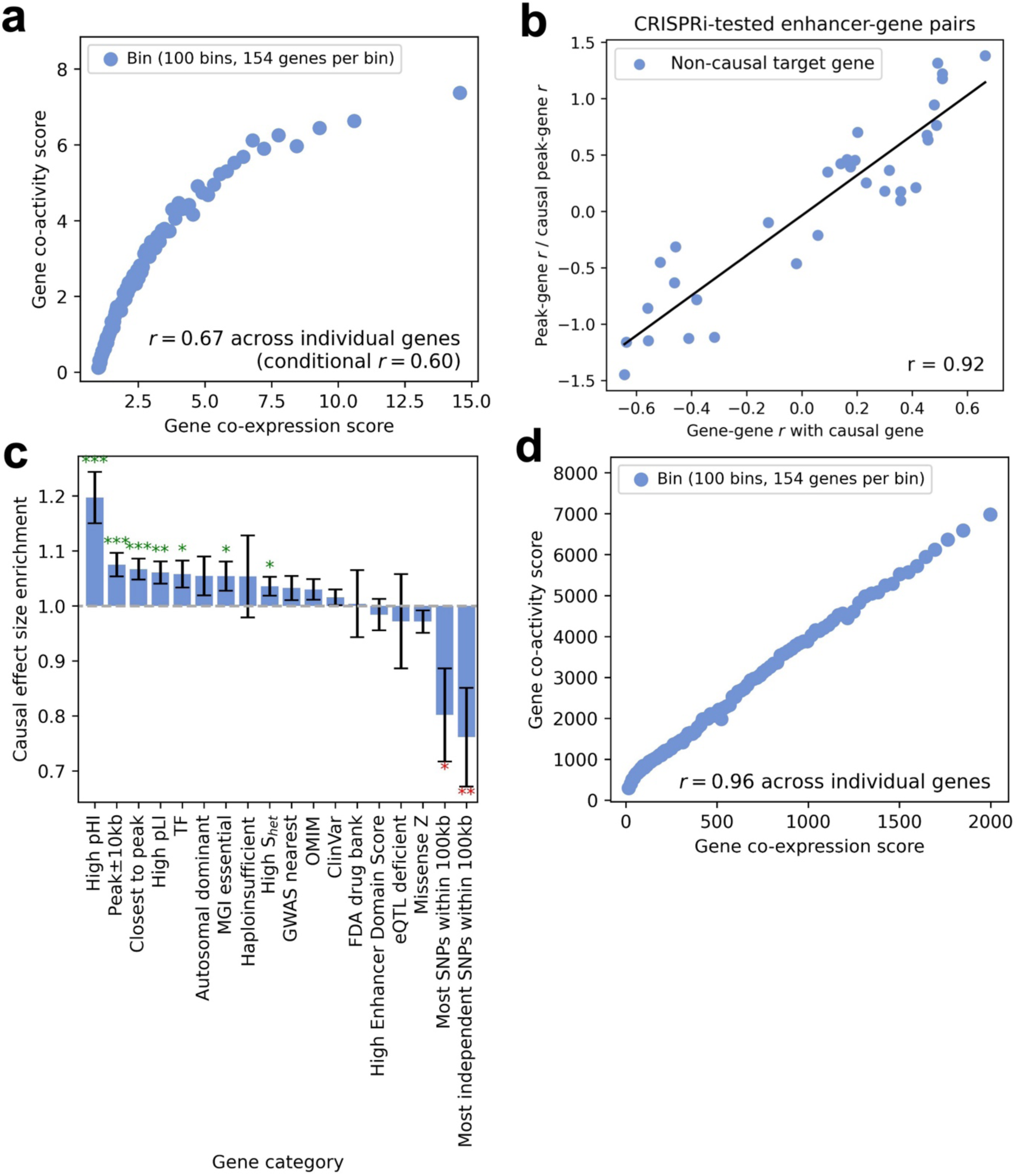
Relationship between gene-gene co-expression and peak-gene co-activity. **a)** Relationship between gene co-expression score and gene co-activity score, computed using genes and peaks within the *cis* window (<1Mb) of each focal gene, across 15,460 genes in the Xu K562^30^ data set. ‘Conditional *r*’ denotes correlation conditioned on number of peaks <1Mb from the focal gene. **b)** Relationship between correlation with the CRISPR-validated causal target gene and correlation with the causal peak (relative to the causal target gene’s correlation with the causal peak) across non-causal target genes tested by CRISPR, restricted to CRISPR positive-negative peaks where ArchR correctly identifies the causal peak-gene link. **c)** Causal effect size enrichment of 18 gene categories in stratified co-expression score regression. Bars and confidence intervals denote estimates and standard errors, respectively, meta-analyzed across data set-cell type pairs. Stars denote meta-analyzed p-values for significant enrichment (green) or depletion (red) (*: p < 0.05, **: p < 0.01, ***: p < 0.001). **d)** Relationship between genome-wide gene co-expression score and genome-wide gene co-activity score, computed using all peaks and genes for each focal gene, across 15,460 genes in the Xu K562^30^ data set. In **a** and **d,** genes are partitioned equally into 100 bins (each represented by 1 point) by gene co-expression score and genome-wide gene co-expression score, respectively. Numerical results are reported in **Supplementary Table 9**, **Supplementary Table 11**, and **Supplementary Table 12**.

To evaluate the gene tagging hypothesis, we analyzed CRISPRi-tested peak-gene pairs^33–44^ in K562 cells, analyzing multiple genes per peak (instead of multiple peaks per gene; **Figure 2b**). We restricted to peaks with both an experimentally validated target gene that is identified by ArchR^14^ and at least one experimentally tested non-target gene overlapping our single-cell data, restricting to the same 3 data set-cell type pairs most relevant to K562 cells (**Table 1**). We measured the correlation (across metacells) of each CRISPR-negative gene with both the CRISPR-positive gene and the focal peak. Correlation with the CRISPR-positive gene was strongly correlated to correlation with the focal peak (**Figure 4b**), corroborating the gene tagging hypothesis. Analyses using correlations across single cells instead of metacells produced similar findings (**Supplementary Figure 31**).

To further evaluate the gene tagging hypothesis, we computed the enrichment of causal peak-gene effect size variance for 18 functional categories of genes by modeling gene co-activity score as a function of co-expression scores stratified by functional gene category, analogous to S-CASC (**Methods**); the 18 functional categories of genes included various metrics for gene essentiality and disease relevance (**Supplementary Table 10**). We observed significant enrichment of causal effect size variance for gene categories defined by gene conservation (high pLI, high pHI) and proximity to peaks (peak±10kb) (ranging from 1.06±0.021 to 1.20±0.047), and significant depletion for genes with many nearby SNPs (e.g. most independent SNPs within 10kb, 0.76±0.090) (**Figure 4c**, **Supplementary Figure 32**, and **Supplementary Table 11**). These findings provide additional support for the gene tagging hypothesis. Analyses using correlations across single cells instead of metacells were generally underpowered (**Supplementary Figure 33**). Analyses excluding the covariate for the number of peaks <1Mb from each gene also produced similar findings (**Supplementary Figure 34**).

We also assessed the relationship (across focal genes) between genome-wide gene co-expression scores and genome-wide gene co-activity scores, defined without restricting to gene-gene or peak-gene pairs with distance <1Mb. Genome-wide gene co-expression scores and genome-wide gene co-activity scores were very strongly correlated (**Figure 4d**, **Supplementary Figure 35** and **Supplementary Table 12**; *r* = 0.72-0.99 in each data set-cell type pair). Analyses using correlations across single cells instead of metacells produced similar findings (**Supplementary Figure 36**).

Although TF binding within a peak explained substantial variance in peak co-accessibility (and peak co-activity) (**Figure 3a-b**), TF binding within a gene promoter explained very little variance in gene co-expression (and gene co-activity) (**Supplementary Table 13**). Instead, gene co-expression is driven by other biological factors^9,50,51^ (as well as gene read depth; see below). In particular, gene co-expression may be driven by peak co-accessibility, as hypothesized by ref.^26^. Indeed, we observed that the average gene-gene *r*^2^ for pairs of genes linked to highly co-accessible pairs of peaks was substantially higher (*r*^2^ *=* 0.28) than for all gene-gene pairs (*r*^2^ *=* 0.047; p-value for difference = 4e-15), supporting this hypothesis (**Supplementary Figure 37**).

We performed 5 secondary analyses to rule out alternative explanations for the correlation between gene co-expression scores and gene co-activity scores. First, to further assess the effects of biological causality, we performed a bivariate regression of gene co-activity score on gene co-expression score and a binary indicator of CRISPR-positive status (restricting to genes with CRISPR-positive and CRISPR-negative peaks), as well as univariate regressions of gene co-activity score on each predictor individually. We determined that gene co-expression score alone explained a much higher proportion of the observed variation in gene co-activity scores, and that the bivariate model did not substantially increase the proportion of variation explained (**Supplementary Figure 38**). Second, to assess gene read depth effects, we performed a bivariate regression of gene co-activity score on gene co-expression score and gene read depth, as well as univariate regressions of gene co-activity score on each predictor individually. We determined that gene co-expression score alone tended to explain a higher proportion of the variance in gene co-activity scores (*R*^2^=0.062-0.57) vs. gene read depth alone (*R*_2_=0.12-0.27), and that the bivariate model did not substantially increase the proportion of variation explained (*R*^2^=0.20-0.58; **Supplementary Figure 39**); we note that gene read depth explained *R*^2^=0.036-0.24 of the variance in gene co-expression scores. Third, to assess cell subtype effects, we computed gene co-expression scores and gene co-activity scores using gene-gene and peak-gene correlations conditional on cell subtype proportions within each metacell of the Luecken^32^ T cell data set. We determined that the correlation between gene co-expression scores and gene co-activity scores remained substantial (*r* = 0.66, vs. *r* = 0.57 for unconditioned correlation), suggesting that major cell subtypes do not explain this correlation (**Supplementary Figure 40**). Fourth, to assess gene measurement noise effects, we introduced additional artificial noise for a subset of genes and recomputed the correlation between gene co-expression scores and gene co-activity scores. We determined that adding noise did not substantially increase the correlation (**Supplementary Figure 41**). Fifth, we assessed the correlation between gene co-expression scores and gene co-activity scores when conditioning on the number of genes with TSS <1Mb from the focal gene’s TSS (in addition to conditioning on the number of nearby peaks; see above). We determined that the correlation remained strong (**Supplementary Figure 42**). We thus conclude that the correlation between gene co-expression scores and gene co-activity scores cannot be explained by biological causality (also see **Figure 4b**), gene read depth, major cell subtypes, varying gene measurement noise, or the number of nearby genes.

These findings demonstrate the pervasive effects of gene-gene tagging on peak-gene associations.

### Accounting for co-accessibility between peaks via fine-mapping improves peak-gene linking

We sought to assess whether peak-level fine-mapping could distinguish causal peaks from tagging peaks, analogous to SNP-level fine-mapping in GWAS^27,52,53^. For each target gene (in all 7 data set-cell type pairs; **Table 1**), we applied SuSiE^27^ to our multiome data (processed using ArchR^14^ or Signac^12,13^ pipelines) to compute posterior inclusion probabilities (PIPs) for each peak, and multiplied marginal peak-gene linking scores (squared peak-gene correlations from ArchR^14^ or Signac^12,13^) by PIPs to obtain fine-mapped peak-gene linking scores (**Data Availability**). We assumed either a single causal peak or multiple causal peaks per gene, and used either flat priors or functionally informed priors obtained from S-CASC (analogous to ref.^54^). We evaluated the fine-mapped peak-gene linking scores using evaluation sets derived from CRISPR^33–44^ and eQTL^55,56^ data (**Data Availability**), computing average enrichment across recall values^22^. Further details are provided in the **Methods** section. We caution that we view these fine-mapping analyses as a proof of concept that accounting for tagging can improve upon existing approaches, and not as a resolution of the state-of-the-art.

We first evaluated fine-mapped peak-gene linking scores using 448 gold-standard links validated by CRISPR^33–44^ (**Supplementary Table 14**). Results for ArchR^14^ are reported **Figure 5a**, **Supplementary Figure 43**, and **Supplementary Table 15**. Fine-mapped peak-gene linking scores substantially outperformed marginal peak-gene linking scores, particularly for single causal peak methods (average enrichment 2.48-2.51x vs. 1.1x for marginal, each p-value for difference < 8e-15); we observed little benefit from including functional priors. Results for Signac^12,13^ are reported in **Figure 5b**, **Supplementary Figure 44**, and **Supplementary Table 16**. Again, fine-mapped peak-gene linking scores substantially outperformed marginal peak-gene linking scores, particularly for single causal peak methods (average enrichment 3.49-3.50x vs. 2.2x for marginal, each p-value for difference < 2e-10), with little benefit from including functional priors.

**Figure 5.**
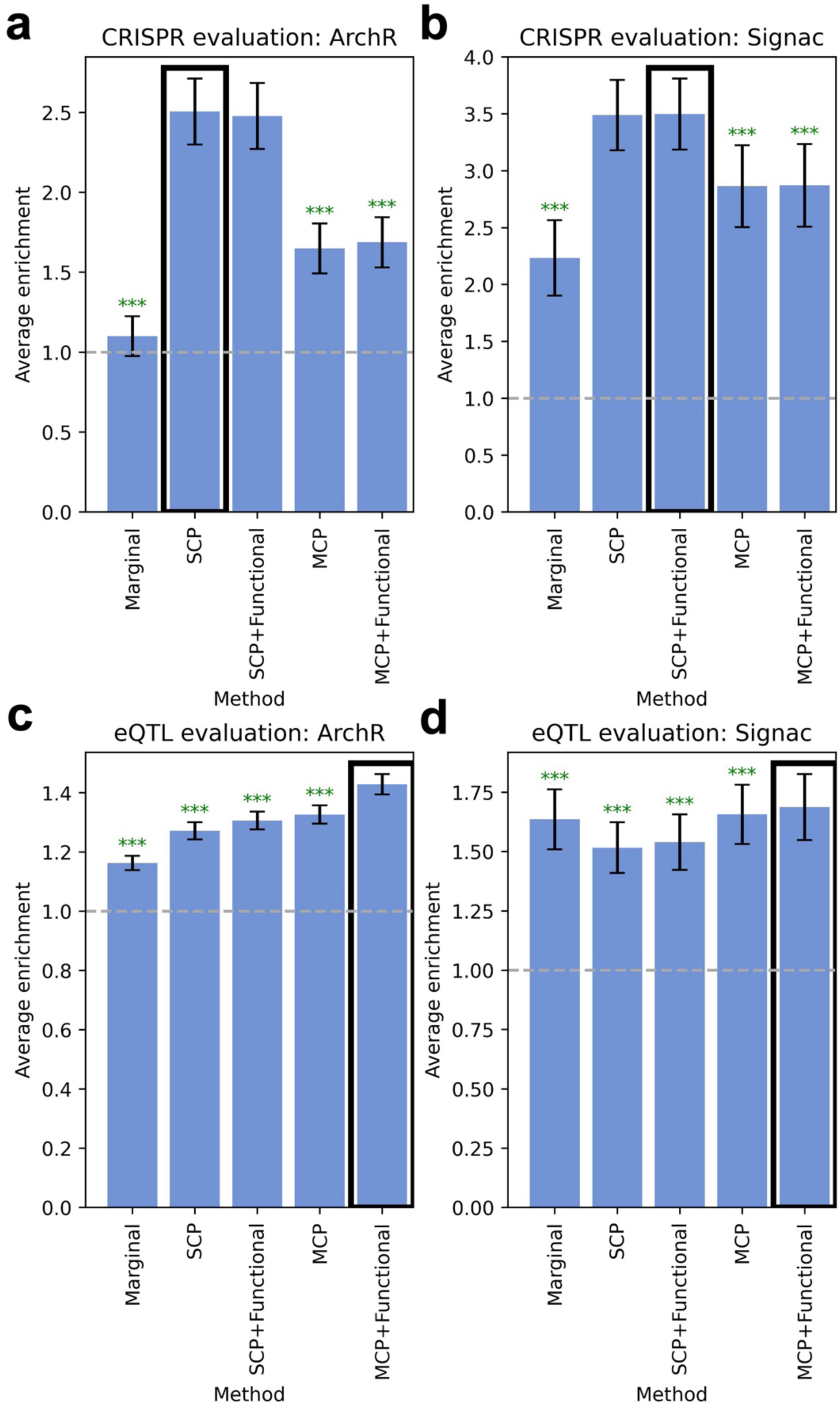
Fine-mapping improves peak-gene linking methods. **a)** Average enrichment across recall values of links predicted by ArchR vs. fine-mapped ArchR scores for 448 links validated by CRISPR. **b)** Average enrichment across recall values of links predicted by Signac vs. fine-mapped Signac scores for 448 links validated by CRISPR. **c)** Average enrichment across recall values of links predicted by ArchR vs. fine-mapped ArchR scores for 39,194 fine-mapped eSNP–eGene pairs attaining maximum PIP > 0.5 across GTEx tissues. **d)** Average enrichment across recall values of links predicted by Signac vs. fine-mapped Signac scores for 39,194 fine-mapped eSNP–eGene pairs attaining maximum PIP > 0.5 across GTEx tissues. Bars and confidence intervals denote estimates and standard errors, respectively, meta-analyzed across data set-cell type pairs. Stars denote meta-analyzed p-values for significant enrichment (green) or depletion (red) (*: p < 0.05, **: p < 0.01, ***: p < 0.001). Green asterisks denote significant underperformance of the focal method vs. the top-performing method (denoted with a black outline). Numerical results are reported in **Supplementary Tables 15-18**. Grey horizontal dashed lines mark 1.0x enrichment. SCP, single causal peak fine-mapping; SCP + Functional, single causal peak functionally informed fine-mapping; MCP, multiple causal peak fine-mapping; MCP + Functional, multiple causal peak functionally informed fine-mapping.

We next evaluated fine-mapped peak-gene linking scores using 39,194 predicted SNP-gene links implicated by fine-mapped eQTL (eSNP–eGene pairs with maximum PIP > 0.5 across GTEx tissues^55,56^). Results for ArchR^14^ are reported in **Figure 5c**, **Supplementary Figure 45**, and **Supplementary Table 17**. Fine-mapped peak-gene linking scores substantially outperformed marginal peak-gene linking scores, with functionally informed multiple causal peak fine-mapping performing best (average enrichment 1.43x vs. 1.16x for marginal, p-value for difference = 1e-48; all fine-mapped peak-gene linking scores outperform marginal, p-values for difference < 6e-8). Results for Signac^12,13^ are reported in **Figure 5d**, **Supplementary Figure 46**, and **Supplementary Table 18**. Functionally informed multiple causal peak fine-mapping again performed best, but with a much smaller improvement (average enrichment 1.69x vs. 1.64x for marginal, p-value for difference = 2e-5). Non-functionally informed multiple causal peak fine-mapping also slightly outperformed marginal peak-gene linking scores (p-value for difference = 0.0057), while single causal peak methods underperformed marginal peak-gene linking scores.

We also sought to assess whether gene-level fine-mapping could pinpoint target genes. For each peak, we applied SuSiE^27^ to our multiome data to compute PIPs for each gene, and multiplied marginal peak-gene linking scores (squared peak-gene correlation from ArchR^14^ or Signac^12,13^) by PIPs to obtain fine-mapped gene-peak linking scores, as above (**Methods**). We determined that both fine-mapped ArchR^14^ peak-gene linking scores and fine-mapped Signac^12,13^ peak-gene linking scores significantly outperformed marginal peak-gene linking scores, on both CRISPR^33–44^ and eQTL^55,56^ evaluation data (**Supplementary Figure 47**).

We performed 3 secondary analyses. First, we performed multiple causal peak fine-mapping of peak-gene correlations across metacells (from ArchR^14^) using peak-peak correlations computed across single cells (from Signac^12,13^), and vice versa. We determined that these approaches did not outperform estimating peak-peak correlations across the same data types (metacells or single cells, respectively) as the focal peak-gene linking method (**Supplementary Figure 48**). Second, we performed fine-mapping restricting to candidate peak-gene pairs with strong correlations instead of all peak-gene pairs. We determined that this approach did not outperform using all candidate peak-gene pairs in fine-mapping (**Supplementary Figure 49**). Third, we compared marginal and fine-mapped scores to a ‘top peak’ approach, retaining only peak-gene links for which the peak attains the highest correlation to the focal gene. We determined that the top-peak approach attained similar enrichment to single causal peak fine-mapping but attained very low recall, and is thus a less desirable approach (**Supplementary Figure 50**).

These initial efforts at fine-mapping peak-gene links using standard approaches^27,54^ suggest that accounting for peak-peak co-accessibility has substantial potential to improve peak-gene links.

### Peak-peak and gene-gene tagging impact peak-gene links at GWAS loci

We have shown that peak-peak tagging and gene-gene tagging effects are present in single-cell multiome data and induce tagging peak-gene associations (**Figure 2** and **Figure 4**, resp.).

Below, we dissect two examples illustrating peak-peak and gene-gene tagging effects in the Xu K562^30^ data set. We selected these examples from 90 peak-gene links (marginal ArchR linking score (*r*^2^) > 0.2025, corresponding to ArchR threshold *r* > 0.45^14,22^) involving a peak harboring a fine-mapped GWAS variant (PIP > 0.5 across 32 blood cell traits^57^; **Supplementary Table 19**), based on single causal variant fine-mapping (as multiple causal variant fine-mapping is not recommended in the absence of in-sample LD^54^) (**Supplementary Table 20**).

First, we dissect an example involving fine-mapping the causal peak for a given gene. ArchR linked the gene *RAP1GAP* to 16 peaks, including the ATAC peak spanning chr1:21687033-21688350 (assigned the highest marginal ArchR linking score among 171 peaks <1Mb from the *RAP1GAP* TSS) (**Figure 6a**, upper panel and **Supplementary Table 21**). The linked peak harbors the fine-mapped GWAS variant rs17423390, fine-mapped to high light scatter reticulocyte percentage of red blood cells (HLRP) with PIP = 0.63^57,58^. *RAP1GAP* has been well studied for its role in the differentiation and survival of erythroid cells (and other hematopoietic cell lineages)^59–63^. HLRP reflects erythroid cell maturity and is an established clinical indicator for anemia^64–66^. Therefore, *RAP1GAP* is a biologically plausible driver of this GWAS association, supporting the link between *RAP1GAP* and chr1:21687033-21688350; an erythroid/hematopoietic progenitor cell line such as K562^67,68^ is also a biologically plausible context for detecting this link. We measured the correlation of each other peak (<1Mb from the *RAP1GAP* TSS) with both chr1:21687033-21688350 and *RAP1GAP*. Squared correlation with chr1:21687033-21688350 was strongly correlated (*r* = 0.90) with linking score to *RAP1GAP* (**Figure 6a**, upper panel, **Supplementary Table 21**, and **Supplementary Figure 51**), suggesting the presence of peak-peak tagging. (We note that measured peak-peak *r*^2^ are likely underestimates of true biological values, due to peak measurement noise.)

**Figure 6.**
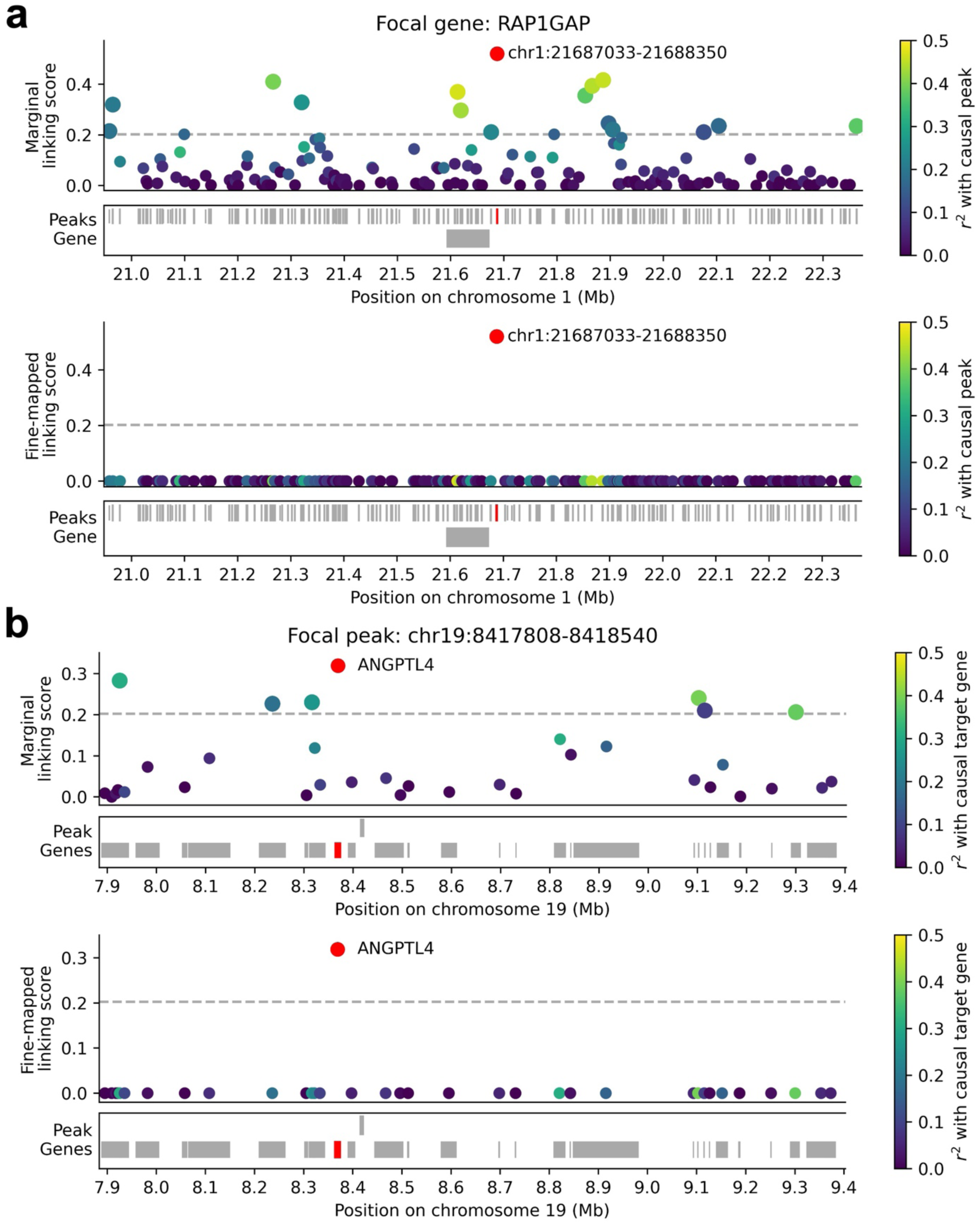
Examples of peak-peak and gene-gene tagging. **a)** Upper panel: Marginal linking score (squared ArchR peak-gene correlation) for each peak <1Mb from the TSS of the gene *RAP1GAP*. *r*^2^ values are reported as dots in the middle of each peak, with the top correlated peak colored in red and other peaks colored based on their squared correlation to the top correlated peak (non-focal genes not shown). Lower panel: Fine-mapped linking score (squared ArchR peak-gene correlation multiplied by fine-mapped PIP) for each peak <1Mb from the TSS of the gene *RAP1GAP*. **b)** Upper panel: Marginal linking score for each gene with TSS <1Mb from the peak spanning chr19:8417808-8418540. *r*^2^ values are reported as dots in the middle of each gene, with the top correlated gene colored in red and other genes colored based on their squared correlation to the top correlated gene (non-focal peaks not shown). Lower panel: Fine-mapped linking score for each gene with TSS <1Mb from the peak spanning chr19:8417808-8418540. In each panel, dashed line denotes linking score threshold corresponding to the ArchR linking threshold. Numerical results are reported in **Supplementary Table 21** and **Supplementary Table 23**.

We determined above that pioneer TFs were important for explaining peak co-accessibility and tagging peak-gene links (**Figure 3**). For each tagging peak, we thus measured the number of pioneer TFs binding both that tagging peak and the focal peak chr1:21687033-21688350 (of 6 total pioneer TFs^46,47^ that bind chr1:21687033-21688350). The number of shared pioneer TFs was moderately correlated to the tagging peak’s squared correlation with chr1:21687033-21688350 (*r* = 0.45) and the tagging peak’s linking score to *RAP1GAP* (*r* = 0.42) across peaks (**Supplementary Figure 52**). In particular, GATA1 is critical for hematopoiesis^69–71^ and has been shown to affect the accessibility of chr1:21687033-21688350 and the expression of *RAP1GAP* in Perturb-multiome experiments^48^. GATA1 binding within a peak was moderately correlated with squared correlation to chr1:21687033-21688350 (*r* = 0.35) and linking score to *RAP1GAP* (*r* = 0.32) across peaks (**Supplementary Table 22**). Other TFs were also moderately correlated with both chr1:21687033-21688350 and *RAP1GAP*, but were not implicated by Perturb-multiome^48^ (**Supplementary Table 22**). These findings provide additional support for this peak-gene link and for the TF-driven origins of peak-peak tagging.

We examined fine-mapped ArchR linking scores for *RAP1GAP.* Fine-mapped ArchR scores (assuming a single causal peak and flat priors) correctly prioritized the link to chr1:21687033-21688350 (the 4th closest peak) and assigned very low linking scores to other peaks (**Figure 6a**, lower panel and **Supplementary Table 21**), demonstrating the ability of fine-mapping to pinpoint causal peak-gene links. While the marginal ArchR method correctly prioritized the link to chr1:21687033-21688350, marginal ArchR linked 15 other peaks to *RAP1GAP* (**Figure 6a**, upper panel and **Supplementary Table 21**). Fine-mapping approaches assuming multiple causal peaks linked 3 additional peaks to *RAP1GAP*; while none harbor fine-mapped GWAS variants, these may be true causal peaks in this or other cellular contexts (**Supplementary Figure 53**). Functionally-informed approaches yielded highly similar results (**Supplementary Figure 53**). Overall, all fine-mapping approaches generated a smaller and thus ‘de-noised’ set of peaks relative to marginal peak-gene linking scores.

Second, we dissect an example involving fine-mapping the target gene for a given peak. ArchR linked the ATAC peak spanning chr19:8417808-8418540 to 7 genes, including the gene *ANGPTL4* (assigned the highest linking score among 35 genes with TSS <1 Mb from the focal peak) (**Figure 6b**, upper panel and **Supplementary Table 23**). The peak chr19:8417808-8418540 harbors the fine-mapped GWAS variant rs12150986, fine-mapped to mean platelet volume with PIP > 0.64^57,58^. *ANGPTL4* encodes an angiopoietin-like protein, a family of proteins well-studied for their role in hematopoiesis (production of erythroid and other blood cells from hematopoietic stem and progenitor cells)^72,73^, and has been broadly implicated in disease^74–76^. *ANGPTL4* has been shown to support the survival and expansion of hematopoietic progenitors^77,78^, with ANGPTL4 treatment linked to increased platelet production in mouse experiments^79^. Therefore, *ANGPTL4* is a biologically plausible driver of this GWAS association, supporting the link between *ANGPTL4* and chr19:8417808-8418540; a hematopoietic progenitor cell line such as K562^67,68^ is also a biologically plausible context for detecting this link. We measured the correlation (across metacells) of each other gene (with TSS <1Mb from the focal peak) with both *ANGPTL4* and the focal peak. Linking score to *ANGPTL4* was strongly correlated (*r* = 0.82) to squared correlation with the focal gene (**Figure 6b**, upper panel, **Supplementary Table 23**, and **Supplementary Figure 54**), suggesting the presence of gene-gene tagging. (We note that measured gene-gene *r*^2^ are likely underestimates of true biological values, due to peak measurement noise.)

We determined above that pioneer TF binding did not explain gene co-expression (see **Supplementary Table 13**) and that co-expression may instead be explained by gene read depth (see **Supplementary Figure 39**) and other biological factors ^9,50,51^ (including peak co-accessibility, see **Supplementary Figure 37**). We determined that gene read depth was only moderately correlated to the tagging gene’s squared correlation with *ANGPTL4* (*r =* 0.13) and the tagging gene’s linking score to chr19:8417808-8418540 (*r* = 0.29) (**Supplementary Figure 55**) across genes. We also determined that the average squared correlation with *ANGPTL4* was substantially higher for tagging genes strongly linked to peaks co-accessible with chr19:8417808-8418540 (*r*^2^ *=* 0.24) than for all tagging genes (*r*^2^ *=* 0.070) (**Supplementary Figure 56**), supporting the hypothesis that peak co-accessibility underlies gene co-expression^26^.

We examined fine-mapped ArchR linking scores for chr19:8417808-8418540 (from fine-mapping genes to a focal peak). Fine-mapped ArchR scores (assuming a single causal gene and flat priors) correctly prioritized the link to *ANGPTL4* (the 3rd closest gene) and assigned very low linking scores to other genes (**Figure 6b**, lower panel and **Supplementary Table 23**), demonstrating the ability of fine-mapping to pinpoint causal peak-gene links. While the marginal ArchR method correctly prioritized the link to *ANGPTL4*, marginal ArchR linked 6 other genes to chr19:8417808-8418540 (**Figure 6b**, upper panel and **Supplementary Table 23**). Fine-mapping approaches assuming multiple causal genes linked 2 additional genes to chr19:8417808-8418540 (*CERS4*, with known roles in ceramide synthesis and cancer progression^80,81^, and *OR7G1*, an olfactory receptor gene involved in odor perception^82^); while these genes seem less functionally relevant, they may be truly causal target genes in this or other cellular contexts (**Supplementary Figure 57**). Functionally-informed approaches yielded highly similar results (**Supplementary Figure 57**). Overall, all fine-mapping approaches generated a smaller and thus ‘de-noised’ set of genes relative to marginal peak-gene linking scores.

These examples demonstrate the effects of peak-peak and gene-gene tagging at specific peaks and genes implicated in GWAS.

## Discussion

In this study, we hypothesized that co-accessibility among scATAC-seq peaks may induce *tagging* peak-gene associations in single-cell multiome data (analogous to tagging associations in GWAS). We examined patterns of peak co-accessibility to assess this hypothesis. We determined that the extent of a peak’s correlation with nearby peaks is strongly related to the extent of its correlation to nearby genes, consistent with tagging and not with other explanations. CRISPRi data and a regression of co-activity scores on co-accessibility scores stratified by functional peak categories (S-CASC) provided additional support for the tagging hypothesis. We explored a TF-driven mechanism for peak tagging and determined that peak co-accessibility is strongly related to TF binding activity (with a particularly strong effect for pioneer TFs, which induce accessibility of closed chromatin). Indeed, although major cell subtypes based on available annotations do not explain the bulk of the correlation between co-accessibility and co-activity, we showed that peak tagging is related to cell subtype / cell state-specific processes (likely reflecting TF activity), and it is possible that tagging effects would largely disappear in the extreme limit of a perfectly homogeneous cell state across cells. We demonstrated that accounting for tagging effects using statistical fine-mapping improves peak-gene linking. We observed similar patterns when investigating gene-gene tagging effects, suggesting that gene co-expression can also impact peak-gene linking. Finally, we highlighted examples in which peak tagging or gene tagging obscures a causal peak-gene link, such that fine-mapping yields more precise predictions of peak-gene links. Overall, we find that tagging has a major impact on efforts to identify peak-gene links. We propose that tagging peak-gene links should be viewed as spurious links, because they implicate incorrect gene regulatory mechanisms (in contrast to tagging associations in GWAS, which are generally viewed as true associations because they implicate correct GWAS loci).

Our work has several implications for downstream analyses. First, researchers applying existing peak-gene linking methods should be aware that non-causal peak-gene links may be induced by tagging peaks or genes. Examining patterns of peak co-accessibility and gene co-expression in the focal data set or incorporating other types of data (e.g. transcription factor binding data or experimental follow-up via CRISPRi^33–43^ or base editing^63^) may help distinguish causal from tagging links. Second, researchers developing new single-cell peak-gene linking methods should consider how to explicitly model peak co-accessibility and/or gene co-expression to more effectively identify causal links; extensions of the fine-mapping approaches explored here^27^, as well as other approaches for causal inference^83–85^, could plausibly improve performance. In particular, peak-gene linking studies are often motivated by the functional interpretation of GWAS associations. A possible future method could jointly examine patterns of peak co-accessibility, gene co-expression, and GWAS summary statistics at a GWAS locus to nominate the causal peak-gene links driving the underlying genetic association.

Our work has several limitations and directions for future research. First, we primarily focused our analyses on blood/immune cell types (for which large and high-quality multiome data sets are available^13,30–32^); in particular, we focused our analyses of transcription factor binding patterns on the K562 and GM12878 cell lines^13,30,31^, for which abundant TF ChIP-seq data are available. While our findings were consistent in all cell types analyzed, exact patterns of peak co-accessibility and gene co-expression vary among cell types (**Supplementary Figure 16**); it will therefore be important to extend these analyses to other multiome data sets^23,24^. Second, in our fine-mapping analysis, we applied fine-mapping software designed for use on genotype data (which is generally highly accurate or well-imputed). While these analyses demonstrated in principle that accounting for tagging effects can improve peak-gene linking, extensions of the fine-mapping algorithm that explicitly model measurement noise in single-cell data may improve performance and should be further explored in future studies^86–88^; indeed, we present the fine-mapping approaches described here as a proof of concept rather than state-of-the-art methods (e.g. functionally informed priors do not always substantially improve performance). Likewise, due to noisy single-cell measurements, relative causal effect size enrichments from S-CASC are likely to be more reliable than absolute causal effect sizes variances. Third, a high fine-mapped linking score does not guarantee biological causality. While fine-mapping linking scores improve upon marginal linking scores (**Figure 5**), experimental validation^33–43,63^ is still needed to conclusively demonstrate a causal link. Fourth, we do not include a large set of existing correlation-based peak-gene linking methods in our analyses^19–21^. While our findings were highly consistent using linking scores computed from two foundational and distinct approaches (ArchR^14^ and Signac^12,13^), it would be valuable to perform these analyses using additional peak-gene linking methods. Fifth, we did not consider TF footprinting^89,90^, i.e. inferring TF binding sites from the strength and position of ATAC reads, in our TF analyses. TF footprinting requires high-resolution ATAC data, which can be challenging to obtain in a single-cell multiome data set, and achieves variable precision^89,91,92^. While our findings were consistent using ChIP-seq data (which provide context-specific, ‘gold standard’ readouts for a smaller set of TFs^89^) and DNA sequence motifs (which provide non-context-specific predictions for a larger set of TFs^49^) to define TF binding sites, it would be valuable to perform these analyses using TF footprinting data, particularly as higher quality multiome data sets amenable to precise footprinting become available^23,24^. Despite these limitations, we have shown that peak co-accessibility and gene co-expression, driven by TF activity and other biological factors, can induce non-causal peak-gene links in single-cell data, and that accounting for these ‘tagging’ effects is crucial for identifying causal peak-gene links.

## Acknowledgements

We thank Benedikt Geiger, Sasha Gusev, Gaspard Kerner, Soumya Raychaudhuri, Ben Strober, and Shamil Sunyaev for helpful discussions, and Gaspard Kerner for providing GWAS fine-mapping results. This research was funded by NIH grants U01 HG012009, R01 MH101244, R37 MH107649, R01 HG006399, R01 MH115676, and R01 HG013083.

## Methods

### Defining peak co-accessibility and peak co-activity

#### Computing co-accessibility scores

We defined the *co-accessibility score* of peak 𝑗 as 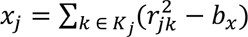, where 𝐾_j_ is the set of all *cis* peaks centered <1Mb from the center of peak 𝑗 (including peak 𝑗 itself) (average number of *cis* peaks per focal peak = 124-147 across data set-cell type pairs; **Supplementary Table 2**), 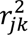 are squared peak-peak correlations (from scATAC-seq data) across metacells, and 𝑏_𝑥_ is a term that captures expected bias in measured peak-peak 𝑟^2^ due to finite sample size (see below). We defined *genome-wide co-accessibility score* analogously, letting 𝐾_j_ be the set of all peaks genome-wide. Co-accessibility scores can be viewed as an analogue of SNP LD scores^28^. We computed peak-peak correlations across metacells using the ArchR^14^ function addCoAccessibility (with parameter maxDist = 1e+06) followed by getCoAccessibility (with parameter corCutOff = −1.5, ensuring that all peak-peak correlations ∈ [−1,1] are returned). We excluded peaks overlapping any promoter region (defined as <1kb to any gene’s TSS^22,29^) from all analyses.

#### Bias correction

We computed the expected bias 𝑏_𝑥_ (see above) for a given data set-cell type pair as follows: we first randomly selected 100,000 peak-peak pairs and computed 𝑎_𝑥_, the average peak-peak 𝑟^2^ across the selected peak-peak pairs. We then generated a randomly downsampled ATAC matrix (containing 50% of metacells) and computed 𝑎_𝑥, downsampled_, the average peak-peak 𝑟^2^ (measured in the 50% downsampled metacells) across the selected peak-peak pairs. Finally, we computed 𝑏_𝑥_ = 𝑎_𝑥, downsampled_ − 𝑎_𝑥_. (Because squared correlations measured across 50% of metacells should contain 2x the amount of upward bias due to noise, this value of 𝑏_𝑥_ provides an estimate of the amount of upward bias due to noise in all metacells.)

#### Computing co-activity scores

We defined the *co-activity score* of peak 𝑗 as 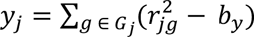, where 𝐺_j_is the set of all *cis* genes with TSS <1Mb from the center of the focal peak 𝑗 (average number of *cis* genes per focal peak = 8-12 across data set-cell type pairs; **Supplementary Table 2**), 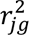 are squared peak-gene correlations (from scATAC-seq and scRNA-seq data) across metacells, and 𝑏_𝑦_ is a term that captures expected bias in measured peak-gene 𝑟^2^ due to finite sample size (analogous to bias correction procedure described above). We also defined *genome-wide co-activity score* as above, where 𝐺_j_is the set of all genes genome-wide. We computed peak-gene correlations across metacells using the ArchR^14^ function addPeak2GeneLinks (with parameter maxDist = 1e+06) followed by getPeak2GeneLinks (with parameters corCutOff = −1.5 and FDRCutOff = 1, ensuring that all peak-gene correlations ∈ [−1,1] are returned). We excluded peaks overlapping any promoter region (defined as <1kb to any gene’s TSS^22,29^) from all analyses.

### Stratified co-accessibility score regression

#### S-CASC model

We propose stratified co-accessibility score regression (S-CASC) as a method to identify functional peak categories that are enriched for causal peak-gene associations and assess the tagging hypothesis. The S-CASC regression relies on the following model:

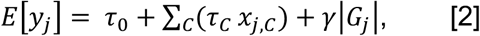

where 𝑦 is the co-activity score of peak 𝑗, 𝜏_0_ is an intercept term, 𝑥_j,𝐶_ is the co-accessibility score of peak 𝑗 with respect to peak category 𝐶, 𝜏_𝐶_ is the per-peak contribution to causal peak-gene effect variance for peak category 𝐶, and the covariate 2𝐺_j_2 is the number of genes with TSS <1Mb from peak 𝑗. 𝑥_j,𝐶_ is defined as 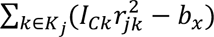, where 𝐾_j_ is the set of all *cis* peaks centered <1Mb from the center of peak 𝑗 (including peak 𝑗 itself) and not overlapping any promoter region, 𝐼_𝐶𝑘_ is the value of peak category 𝐶 for peak 𝑘 (for binary peak categories, 𝐼_𝐶𝑘_equals 1 if peak 𝑘 belongs to category 𝐶 and 0 otherwise; for probabilistic peak categories, 𝐼_𝐶𝑘_ ∈ [0,1]; see below), and other terms are defined as above. S-CASC can be viewed as an analogue of stratified LD score regression^4^.

#### Estimating per-peak causal effect size variance

We defined per-peak causal effect size variance for peak 𝑗 as 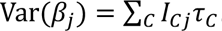 After computing co-activity scores (𝑦_j_) and stratified co-accessibility scores (𝑥_j,𝐶_), we performed the multiple regression in equation 2 (regressing 𝑦_j_on 𝑥_j,𝐶_) to obtain estimates 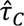 of 𝜏_𝐶_ for each peak category 𝐶. We then estimated per-peak causal effect size variances 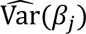 using 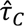.

#### Estimating causal effect size enrichment of a peak category

We defined the total causal effect size variance in peak category 𝐶 as 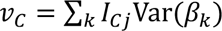 . We then defined the causal effect size enrichment of peak category 𝐶 as follows:

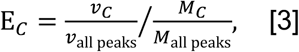

where 𝑣_all peaks_ is the total causal effect size variance of all peaks, 𝑀_𝐶_ is defined as 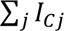 (for a binary peak category, 𝑀_𝐶_ is the number of peaks in category 𝐶), and 𝑀_all peaks_ is the total number of peaks. We use our estimates of 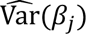 to estimate 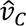 and 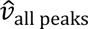. We then use these estimates to estimate 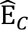 for each peak category 𝐶. (Due to peak measurement noise, we focus on 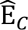 rather than 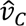 to assess the relative importance of each peak category.)

#### Computing jackknife standard errors

For a given data set-cell type pair, we estimated standard errors for each 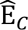 using a jackknife with blocks defined by chromosomes, analogous to previous work^4,28^. (We first obtained jackknifed estimates of 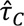, then used these estimates to obtain jackknifed estimates of 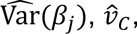, and 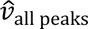, and 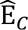.)

#### Computing meta-analyzed estimates and standard errors

We computed meta-analyzed 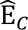 estimates and standard errors via random effect meta-analysis across data set-cell type pairs.

#### Defining peak categories

We implemented S-CASC using 19 functional peak categories (including 18 binary categories and 1 probabilistic peak annotation), in addition to a category defined by all peaks, as described in ref.^4^ For each binary category, we defined an ATAC peak belonging to that category as a peak overlapping any genomic region in that category.

The functional peak categories were defined as follows (further details are provided in **Supplementary Table 3**):

- 5 binary genomic categories (“Coding”, “3’ UTR”, “5’ UTR”, “Intron”, “Repressed”) from the S-LDSC^4^ baseline model.
- 10 binary categories defined by activating or repressive histone modifications (7 activating: H3K4me1, H3K4me2, H3K4me3, H3K9ac, H3K27ac, H3K36me3, H3K79me2; and 3 repressive: H3K27me3, H3K9me1, H3K9me3), as measured by histone ChIP-seq experiments in K562 cells publicly on the ENCODE portal (accessions are provided in **Supplementary Table 3**; for each experiment, we used the ‘pseudoreplicated peaks’ file in ‘bed narrowPeak’ format).
- 2 binary gene proximity-based categories (“TSS±10kb”, “TSS±100kb”) defined by any overlap between a peak and the union of genomic windows TSS±10kb (or TSS±100kb, respectively) across all genes.
- 1 binary category defined as “Closest to TSS”. For each gene, we identified the single ATAC peak closest to the focal gene’s TSS. We then defined the set of these peaks as the “Closest to TSS” category.
- 1 probabilistic constraint-based annotation (“Constraint”) defined by the proportion of bases in an ATAC peak overlapping any constrained region defined by ref.^93^

#### Genome-wide S-CASC

As stated above, to compute 𝑥_j,𝐶_, we restricted to the set of all *cis* peaks centered <1Mb from peak 𝑗. Computing 𝑥_j,𝐶_ without this restriction (and instead using all peaks genome-wide) yielded 𝑥_j,𝐶_ that were highly correlated across peak categories, leading to substantial multicollinearity in the S-CASC regression and inflated standard errors for 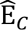, such that all 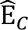 were statistically indistinguishable from 1.

### Gene co-expression and gene co-activity scores

#### Computing gene co-expression scores

We defined the *gene co-expression score* of gene 𝑖 as 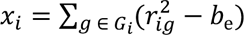, where 𝐺_𝑖_ is the set of all genes with TSS <1Mb from the TSS of focal gene 𝑖 (including gene 𝑖 itself) (average number of *cis* genes per focal gene = 31-44 across data set-cell type pairs; **Supplementary Table 9**), 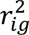 are squared gene-gene correlations (from scRNA-seq data) across metacells, and 𝑏_e_ is a term that captures expected bias in measured gene-gene 𝑟^2^ due to finite sample size (analogous to the bias correction procedure described above). We also defined *genome-wide gene co-expression score* as above, where 𝐺_𝑖_ is the set of all genes genome-wide. We computed gene-gene correlations using the cor() function on ArchR^14^ gene expression metacell matrices.

#### Computing gene co-activity scores

*\*We defined the *gene co-activity score* of gene 𝑖 as 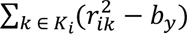, where 𝐾_𝑖_ is the set of all peaks centered <1Mb from the TSS of gene 𝑖 (average number of *cis* peaks = 64-102 across data set-cell type pairs; **Supplementary Table 9**), 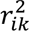 are squared gene-peak correlations (from scATAC-seq and scRNA-seq data) across metacells, and 𝑏_𝑦_ is a term that captures expected bias in measured peak-gene 𝑟^2^ due to finite sample size (analogous to the bias correction procedure described above; we note that the same 𝑏_𝑦_ value is used to compute peak co-activity scores and gene co-activity scores). We also defined *genome-wide gene co-activity score* analogously, letting 𝐾_𝑖_ be the set of all peaks genome-wide. We excluded peaks overlapping any promoter region (defined as <1kb to any gene’s TSS^22,29^) from all analyses. We computed peak-gene correlations across metacells as described above.

#### Stratified co-expression score regression model

We implemented stratified co-expression score regression as a method to identify functional gene categories that are enriched for causal peak-gene associations and assess the gene tagging hypothesis. This regression relies on a model analogous to the S-CASC model:

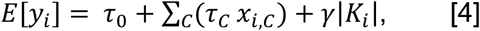

where 𝑦 is the gene co-activity score of gene 𝑖, 𝜏_0_ is an intercept term, 𝑥_𝑖,𝐶_ is the co-expression score of gene 𝑖 with respect to gene category 𝐶, 𝜏_𝐶_ is the per-peak contribution to causal peak-gene effect variance for gene category 𝐶, and the covariate |𝐾_𝑖_| is the number of peaks centered <1Mb from the TSS of gene 𝑖. 𝑥_𝑖,𝐶_ is defined as 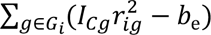, where 𝐺_𝑖_ is the set of all *cis* genes with TSS <1Mb from the TSS of gene 𝑖 (including gene 𝑖 itself), 𝐼_𝐶g_ is the value of gene category 𝐶 for gene 𝑖 (for binary gene categories, 1 if peak 𝑘 belongs to category 𝐶 and 0 otherwise), and other terms are defined as above.

#### Estimating per-gene causal effect sizes and causal effect size enrichment

We defined and estimated per-gene causal effect size variances, total causal effect size variance in each gene category, and causal effect size enrichment for each gene category, as described above. We also computed jackknife standard errors and meta-analyzed enrichments across data set-cell type pairs, as described above.

#### Defining gene categories

We implemented S-CESC using 18 functional gene categories, in addition to a category defined by all genes. These categories spanned 16 categories described in ref.^94^ and 2 peak proximity-based categories; all were binary categories.

The 16 functional gene categories from ref.^94^ are briefly described below (references and further details are provided in **Supplementary Table 10**; we excluded the gene category “Olfactory receptors”^94^ from our analyses because the Luecken BMMC^32^ data set did not include any genes in this category):

- 6 categories reflecting gene essentiality or selection against protein-altering variants (“High pLI”, “High pHI”, “High Shet”, “Haploinsufficient”, “Missense Z”, “MGI essential”).
- 3 categories reflecting clinical significance or pathogenicity (“OMIM”, “ClinVar”, “Autosomal dominant”).
- 2 categories defined by proximity to a large number of SNPs (“Most SNPs within 100kb”) or independent (*r* < 0.1) SNPs (“Most independent SNPs within 100kb”).
- 2 categories defined by the presence or absence of genetic associations (“GWAS nearest”, “eQTL-deficient”).
- 1 category defined by genes encoding transcription factors (“TFs”).
- 1 category defined by high predicted connectedness to enhancers (“High Enhancer Domain Score”).
- 1 category defined by established drug targets (“FDA drug bank”).

The 2 peak proximity-based categories are briefly described below:

- 1 category (“Peak±10kb”) defined by the overlap of a gene’s TSS with the 10kb genomic window surrounding any peak. (We note that the “Peak±100kb” category included most genes and was thus uninformative.)
- 1 category defined as “Closest to peak”. For each peak, we identified the single gene with TSS closest to the focal peak. We then defined the set of these genes as the “Closest to peak” category.

### Single-cell multiome data sets

We downloaded 4 publicly available scRNA+ATAC-seq multiome data sets (Xu K562^30^, Satpathy K562^31^, SHARE-seq LCL^13^, and Luecken BMMC^32^; **Table 1** and **Supplementary Table 1**) (see **Data Availability**) that spanned immune and blood cell types. For the Luecken BMMC^32^ data set, we analyzed major cell types that spanned >5,000 cells, yielding 7 data set-cell type pairs.

We obtained publicly available quality controlled gene expression and ATAC peak matrices for the Luecken BMMC data sets, and processed the Xu K562^30^, Satpathy K562^31^, SHARE-seq LCL^13^ data sets using both Seurat (v5.3.0)^95^ and ArchR (v1.0.3)^14^. We retained all cells from each data set passing quality control filters on total ATAC and RNA reads, nucleosome signal, and TSS enrichment, following ref.^22^ (**Supplementary Table 24**). We next identified ‘peaks’ of open chromatin in each data set using macs2^96^ and excluded non-standard chromosomes and blacklist regions. For the Luecken BMMC data set, which spans multiple cell types, we defined cell types using labels provided by the authors^32^. These steps yielded matrices of gene expression and ATAC peak counts in individual cells for each data set-cell type pair.

We pre-processed each data set-cell type pair using the ArchR^14^ pipeline. We first performed iterative LSI-based dimensionality reduction separately on RNA and ATAC peak matrices using addIterativeLSI() with default parameters. We then combined the RNA and ATAC LSI-based reductions into a joint dimension reduction using addCombinedDims() with default parameters. When computing peak-peak and peak-gene correlations (see above), ArchR^14^ generates metacell-level RNA and ATAC matrices from the joint dimension reduction, by defining metacells (low-level cell aggregates) using *k*-nearest neighbors and aggregating RNA and ATAC peak counts across single cells within each metacell. After computing peak-gene correlations, we extracted RNA and ATAC metacell matrices from the metadata of the output from peak-gene linking.

### Transcription factor binding site data

#### ChIP-seq data

We obtained publicly available ChIP-seq data for 303 transcription factors (TFs) in K562 cells and 148 TFs in GM12878 cells from the ENCODE portal, using the following search terms: “TF ChIP-seq” for assay title, “*Homo sapiens*” for organism, “K562” or “GM12878” for biosample, “Not perturbed” for perturbation, and “GRCh38” for genome assembly. We further restricted to assays with biosample summary either “*Homo sapiens* K562” or “*Homo sapiens* GM12878” (removing assays with “genetically modified using CRISPR” in the biosample summary). For each experiment, we downloaded the “IDR thresholded peaks” file in “bed narrowPeak” format, which contained quality-controlled ChIP-seq peaks (accessions are provided in **Supplementary Table 7**).

#### Defining TFBS for ATAC peaks

We analyzed transcription factor binding sites (TFBS) in 3 data set-cell type pairs (Xu K562^30,31^ and Satpathy K562^31^ for K562 TFs, SHARE-seq LCL^13^ for GM12878 TFs; **Table 1**). For a given TF and data set-cell type pair, we defined a TF binding in an ATAC peak as any overlap of the ATAC peak with a ChIP-seq peak for that TF in the corresponding cell type; we thus generated a binarized TFBS matrix for ATAC peaks in each data set-cell type pair (in which an entry of 1 for a given ATAC peak-TF pair indicates that ≥1 ChIP-seq peaks for the focal TF overlapped the focal ATAC peak, and an entry of 0 indicates that no ChIP-seq peaks for the focal TF overlapped the ATAC peak). We further computed the number of TFs that bind in each ATAC peak by summing binarized matrix entries across TFs. For a given peak-peak pair, we also computed the number of TFs that bind both peaks as the dot product between the binarized TFBS vectors for both peaks.

#### Defining TFBS for genes

For a given TF and data set-cell type pair, we also defined a TF binding a gene as any overlap of the gene’s promoter (TSS±1kb) with a ChIP-seq peak for that TF in the corresponding cell type; we thus generated a binarized TFBS matrix for genes in each data set-cell type pair. We further computed the number of TFs that bind in each gene’s promoter by summing binarized matrix entries across TFs. For a given peak-gene pair, we also computed the number of TFs that bind both the peak and the gene’s promoter as the dot product between the binarized TFBS vectors for the focal peak and gene.

#### Defining pioneer TFs

We obtained a list of 43 established pioneer TFs from refs.^46,47^ We intersected this set with the set of 303 TFs in our K562 ChIP-seq data and 148 TFs in our GM12878 ChIP-seq data to define 9 known pioneer TFs in K562 and 8 known pioneer TFs in GM12878. We define ‘non-pioneer’ TFs as TFs included in our ChIP-seq data but not included in the set of established pioneer TFs^46,47^; we note that the label ‘non-pioneer’ does not exclude the possibility that these TFs do exhibit pioneer activity in these or other cell types. We further note that the distinct sets of pioneer TFs in each cell type are a function of ChIP-seq data availability and do not necessarily reflect cell type-specific TF abundance or activity.

### Fine-mapping peak-gene links

#### Computing posterior inclusion probabilities

We performed peak-level fine-mapping using SuSiE^27^ (v0.14.2) to distinguish causal from tagging peaks. For a focal gene, we ran the function susie(), using as input standardized expression of the focal gene (“y” parameter) and standardized accessibility of candidate peaks (“X” parameter), to generate posterior inclusion probabilities (PIPs) for each candidate peak.

#### Fine-mapping approaches

We performed fine-mapping assuming either a single causal peak (with susie() parameter L = 1) or multiple causal peaks (with susie() parameter L = 10 or the number of candidate peaks, if there were <10 candidate peaks). We also performed fine-mapping assuming either flat priors or functionally informed priors, defined by per-peak causal effect size variances from S-CASC and supplied to the susie() parameter prior_weights (see above; analogous to ref.^54^). We thus defined 4 peak-gene fine-mapping approaches (single causal peak, single causal peak informed, multiple causal peak, multiple causal peak informed).

#### Defining marginal and fine-mapped linking scores

We define “marginal peak-gene linking score” as squared peak-gene correlation (from ArchR^14^ or Signac^12,13^; see below). We define “fine-mapped peak-gene linking score” as marginal peak-gene linking score multiplied by the fine-mapped PIP for the focal peak-gene pair. We computed marginal and fine-mapped linking scores for each candidate peak-gene link in each data set-cell type pair. We define “candidate peak-gene links” as the set of peak-gene pairs with peak-TSS distance <1Mb (for analyses involving the CRISPR^33–44^ evaluation set, we further restricted the set of “candidate peak-gene links” to peak-gene pairs tested by a CRISPR experiment^33–44^).

#### Generating marginal and fine-mapped linking scores from Signac

For each data set-cell type pair (**Table 1**), we generated separate fine-mapped scores for the ArchR^14^ peak-gene linking method (using metacell-level data pre-processed by ArchR^14^ and standardized across metacells) and the Signac^12,13^ peak-gene linking method (using single-cell-level data pre-processed by the Signac^12,13^ pipeline and standardized across single cells). To compute Signac linking scores, we first generated binarized ATAC matrices (in which an entry of 1 for a given peak-cell pair indicates that ≥1 ATAC fragments overlap the peak and an entry of 0 indicates that no ATAC fragments overlap the peak) and normalized gene expression matrices (by running SCTransfrom^86^ on the RNA counts matrix). We computed peak-gene correlations by running the Signac^12,13^ LinkPeaks() function (with parameters: 1e+06, pvalue_cutoff = 1, score_cutoff = 0; these values of “pvalue_cutoff” and “score_cutoff” ensured that all peak-peak correlations ∈ [−1,1] were returned).

#### Gene-level fine-mapping

We also performed gene-level fine-mapping using SuSiE^27^ to distinguish causal from tagging target genes. For a focal peak, we ran the function susie(), using as input standardized accessibility of the focal peak (“y” parameter) and standardized expression of candidate genes (“X” parameter), to generate posterior inclusion probabilities (PIPs) for each candidate gene. As above, we performed fine-mapping assuming either a single causal target gene or multiple causal target genes (with susie() parameter L = 1 or 10, respectively). We also performed fine-mapping assuming either flat priors or functionally informed priors, defined by per-peak causal effect size variances from stratified co-expression score regression. We defined “marginal peak-gene linking score” and “fine-mapped peak-gene linking score” as above.

### Evaluating fine-mapped peak-gene links

To assess the performance of peak-gene linking methods, we used an enrichment-based evaluation framework and peak-gene link evaluation sets described in ref.^22^ We defined 2 evaluation sets of peak-gene links derived from eQTL^55,56^ and CRISPR^33–44^ data (see **Data Availability**) (also described in ref.^22^). Brief descriptions of the evaluation sets are provided below:

The eQTL evaluation set is comprised of 39,194 fine-mapped eSNP-eGene pairs attaining a posterior inclusion probability (PIP) > 0.5 in any GTEx tissue^55,56^. Fine-mapping was conducted by ref.^56^ (by applying the SuSiE^27^ fine-mapping method to 49 GTEx tissues^55^). For each data set-cell type pair, we defined a peak-gene pair as a “true” link if the focal peak harbored any SNP with PIP > 0.5 to the focal gene; we classified all other peak-gene pairs as “false” links.

The CRISPR evaluation set (defined by ref.^44^) is comprised of 448 enhancer-gene links experimentally validated by CRISPR interference experiments^33–43^. Validated links are defined as those marked “TRUE” in the “Regulated” column of the combined CRISPR evaluation data set (see Data Availability). A full list of CRISPR studies included in this evaluation set is reported in **Supplementary Table 14**. For each data set-cell type pair, we defined a peak-gene pair as a “true” CRISPR link if the focal peak overlapped any enhancer with a validated causal effect on the focal gene; we defined a peak-gene pair as a “false” CRISPR link if the focal peak overlapped any genomic region experimentally tested for an effect on the focal gene and found to be non-causal.

We used an enrichment-based metric (also described in ref.^22^), *average enrichment across recall values*, to assess the ability of each linking method to prioritize (assign higher linking scores to) “true” links over “false” links (as defined by an evaluation set of peak-gene links).

For a given evaluation set and given method, we ranked candidate peak-gene links by linking score. At each unique linking score *c* assigned to any candidate peak-gene link, we computed enrichment and recall as follows:

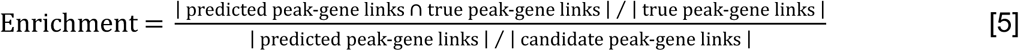

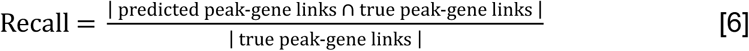

where “predicted peak-gene links” denotes the binarized set of peak-gene links attaining linking score ≥ *c*, “true peak-gene links” denotes the binarized set of “true” peak-gene links defined by the evaluation set (intersected with the set of “candidate peak-gene links”), and “candidate peak-gene links denotes the set of peak-gene pairs with peak-TSS distance <1Mb (as noted above, for analyses involving the CRISPR^33–44^ evaluation set, we further restricted the set of “candidate peak-gene links” to peak-gene pairs tested by a CRISPR experiment^33–44^). We restricted the set of “candidate peak-gene links” to links involving genes with ≥ 1 “true” link. Intuitively, equation 4 quantifies the extent of overlap between predicted peak-gene links and evaluation SNP-gene links relative to the overlap expected by chance.

For a given evaluation set and set of methods, we used equation 5 and equation 6 to construct enrichment-recall curves and observed 𝑅 ≤ 1, the maximum recall achieved by all methods (e.g. if ArchR and fine-mapped ArchR attained maximum recall values of 𝑅_1_ and 𝑅_2_, respectively, we defined 𝑅 as the minimum of 𝑅_1_ and 𝑅_2_. For each method, we then defined *average enrichment across recall values* (or *average enrichment*, shortened) as follows:

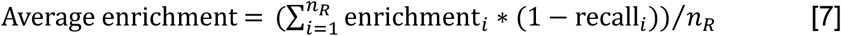

where 𝑛_𝑅_ denotes the total number of “true” evaluation links corresponding to recall 𝑅 and (recall_𝑖_, enrichment_𝑖_) for 𝑖 ∈ {1 … 𝑛} denote the values of each point on the enrichment-recall curve. Multiplying enrichment_𝑖_ by (1 − recall_𝑖_) upweights enrichments at lower recall values, prioritizing method performance at high score thresholds. Intuitively, this metric can be interpreted as a weighted area under the enrichment-recall curve.

We obtained standard errors on average enrichment for each method, as well as standard errors and p-values on the difference in average enrichment between two methods, by bootstrapping genes using 1,000 bootstrap iterations (as in ref.^22^).

## Data Availability

All single-cell multiome data sets analyzed are freely publicly available. The Xu K562^30^ data set is available at https://www.ebi.ac.uk/biostudies/arrayexpress/studies/E-MTAB-11264. The Satpathy K562^31^ data set is available in the IGVF portal (https://data.igvf.org/) (accession code IGVFDS2018URDY). The SHARE-Seq LCL^13^ and Luecken BMMC^32^ data sets are available at Gene Expression Omnibus (accession codes GSE140203 and GSE194122, respectively).

Peak co-accessibility and co-activity scores, gene co-expression and co-activity scores, and fine-mapped peak-gene linking scores have been made publicly available at https://doi.org/10.5281/zenodo.18643360.

TF ChIP-seq data sets are available on the ENCODE portal (https://www.encodeproject.org/). File accessions are listed in **Supplementary Table 7**. TF motif coordinates from ref.^49^ are available at https://resources.altius.org/~jvierstra/projects/motif-clustering/releases/v1.0/.

Fine-mapped GTEx eQTL data from ref.^56^ are available at https://www.finucanelab.org/data. GTEx eQTL summary statistics are available at https://www.gtexportal.org/home/downloads/adult-gtex/qtl. The CRISPR data set defined by ref.^44^ and derived from refs.^33–43^ is available at https://github.com/EngreitzLab/CRISPR_comparison/blob/main/resources/crispr_data/EPCrisprBenchmark_combined_data.training_K562.GRCh38.tsv.gz.

GWAS summary statistics for 32 blood cell traits were made available by ref.^97^ at https://alkesgroup.broadinstitute.org/.

Coordinates for the set of genes analyzed (from ref.^10^) are available at https://github.com/EngreitzLab/CRISPR_comparison/blob/main/resources/genome_annotations/CollapsedGeneBounds.hg38.bed.

## Code Availability

Code to compute peak-level and gene-level scores, run S-CASC, and perform peak-gene fine-mapping has been made publicly available at https://github.com/elizabethdorans/scTagging.

Tutorials for existing methods ArchR and Signac have been made publicly available at https://github.com/elizabethdorans/E2G_Method_Tutorials.

Code implementing ArchR^14^ is publicly available at https://github.com/GreenleafLab/ArchR. Code implementing Signac^12,13^ is publicly available at https://github.com/stuart-lab/signac. Code implementing SuSiE^27^ is publicly available at https://github.com/stephenslab/susieR. We used ArchR v1.0.2, and Signac v1.10.0, and SuSiE v0.14.2.

