## SupplementaryFigures1-57 for "Distinguishing causal from tagging enhancers using single-cell multiome data"

### Supplementary Figures

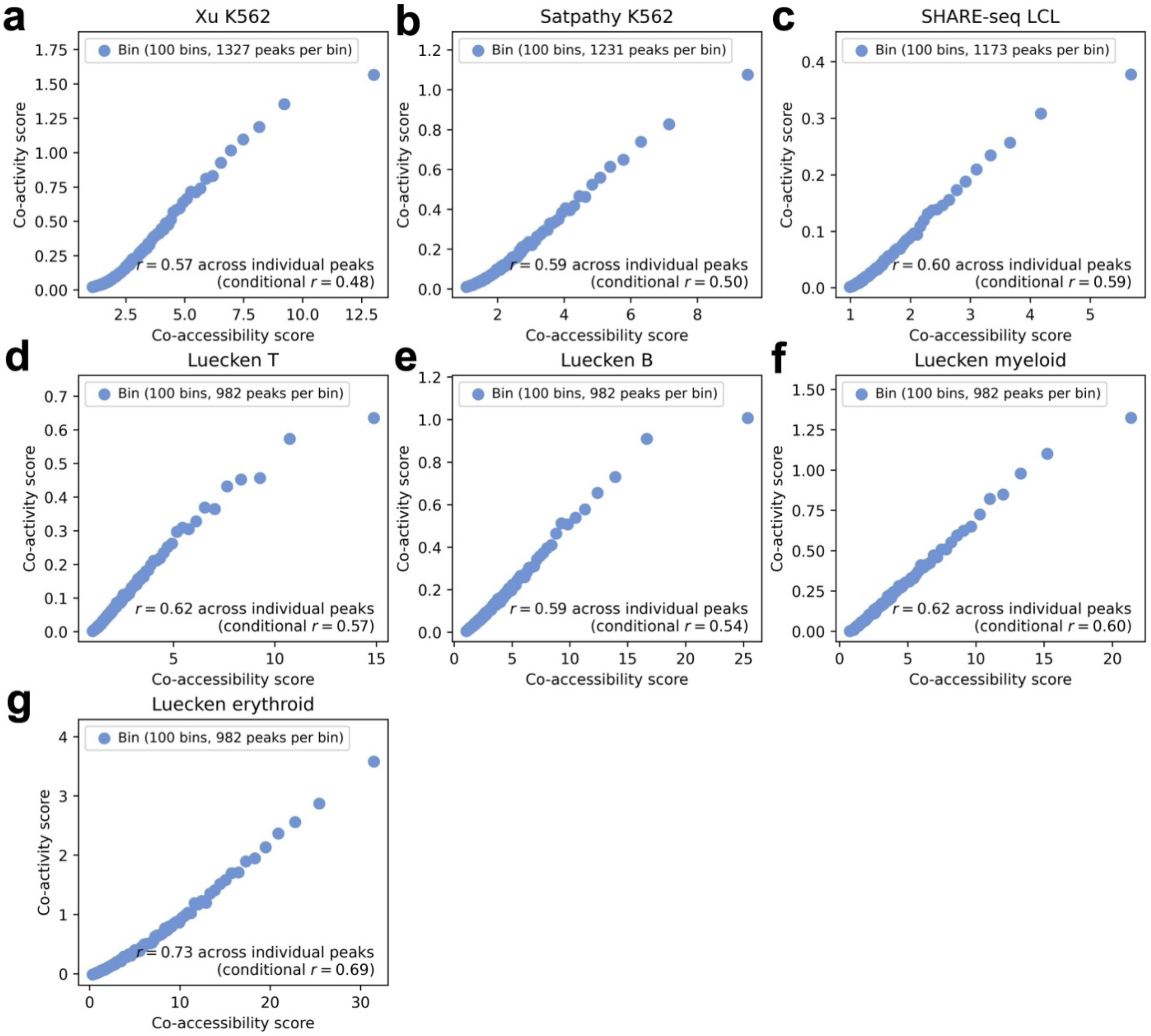


**Supplementary Figure 1. Relationship between co-accessibility score and co-activity score.** Relationship between co-accessibility score and co-activity score across peaks in the **a)** Xu K562, **b)** Satpathy K562, **c)** SHARE-seq LCL, **d)** Luecken T, **e)** Luecken B, **f)** Luecken myeloid, and **g)** Luecken erythroid data set-cell type pairs, computed using peaks and genes within the *cis* window (<1Mb) of each focal peak. 'Conditional *r*' denotes correlation conditioned on number of genes <1Mb from the focal peak. Peaks are partitioned equally into 100 bins (each represented by 1 point) by co-accessibility score. Numerical results are reported in **Supplementary Table 2**.

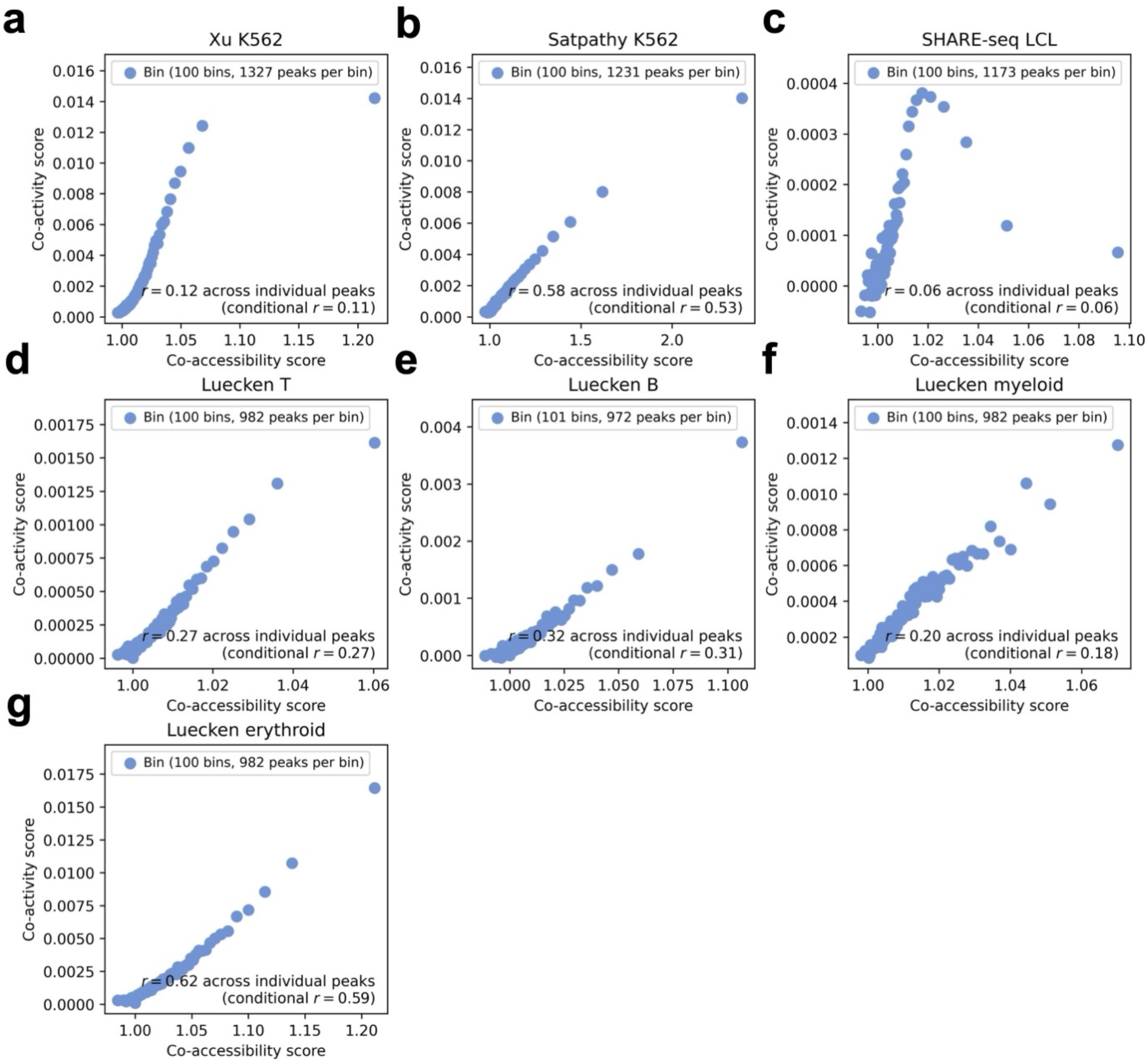


**Supplementary Figure 2. Relationship between co-accessibility score and co-activity score computed using correlations across single cells (instead of metacells).** Relationship between co-accessibility score and co-activity score across peaks in the **a)** Xu K562, **b)** Satpathy K562, **c)** SHARE-seq LCL, **d)** Luecken T, **e)** Luecken B, **f)** Luecken myeloid, and **g)** Luecken erythroid data set-cell type pairs, computed using peaks and genes within the *cis* window (<1Mb) of each focal peak, with peak-peak and peak-gene correlations computed across single cells (instead of metacells). 'Conditional *r*' denotes correlation conditioned on number of genes <1Mb from the focal peak. Peaks are partitioned equally into 100 bins (each represented by 1 point) by co-accessibility score.

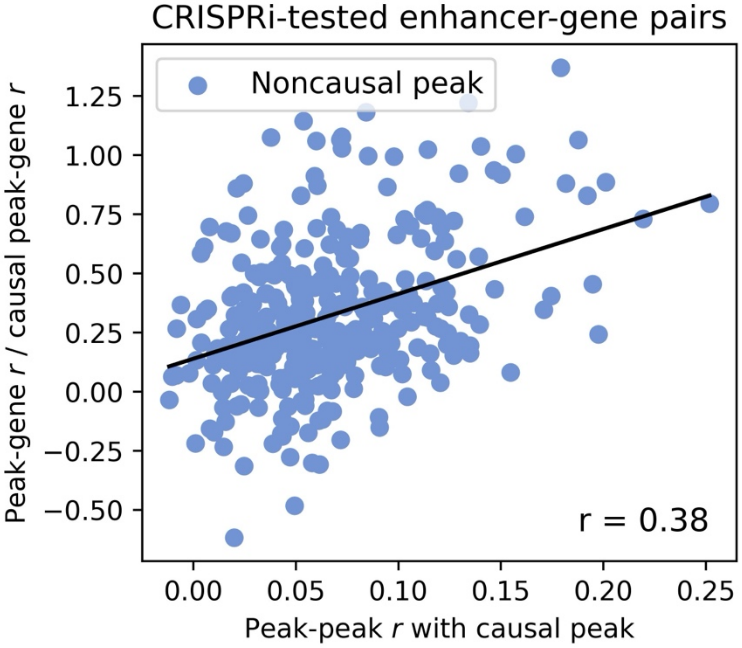


**Supplementary Figure 3. Relationship between non-causal peak-gene correlation and tagging peak correlation using correlations computed across single cells (instead of metacells).** Relationship between correlation with the CRISPR-validated causal peak and correlation with the target gene (relative to the causal peak's correlation with the target gene) across non-causal peaks tested by CRISPRi, with correlations computed across single cells (instead of metacells). We analyzed CRISPRi-tested peak-gene pairs, restricting to 28 genes with both an experimentally validated causal (CRISPR-positive) peak that is identified by Signac (with peak-gene correlation > 0.05 and p-value < 0.05) and at least one experimentally tested non-causal (CRISPR-negative) peak overlapping our single-cell data. We restricted this analysis to the 3 data set-cell type pairs most relevant to K562 cells: Xu K562, Satpathy K562, and Luecken erythroid. We measured the correlation of each CRISPR-negative peak with both the CRISPR-positive peak and the focal gene. Correlation with the CRISPR-positive peak was strongly correlated to correlation with the focal gene.

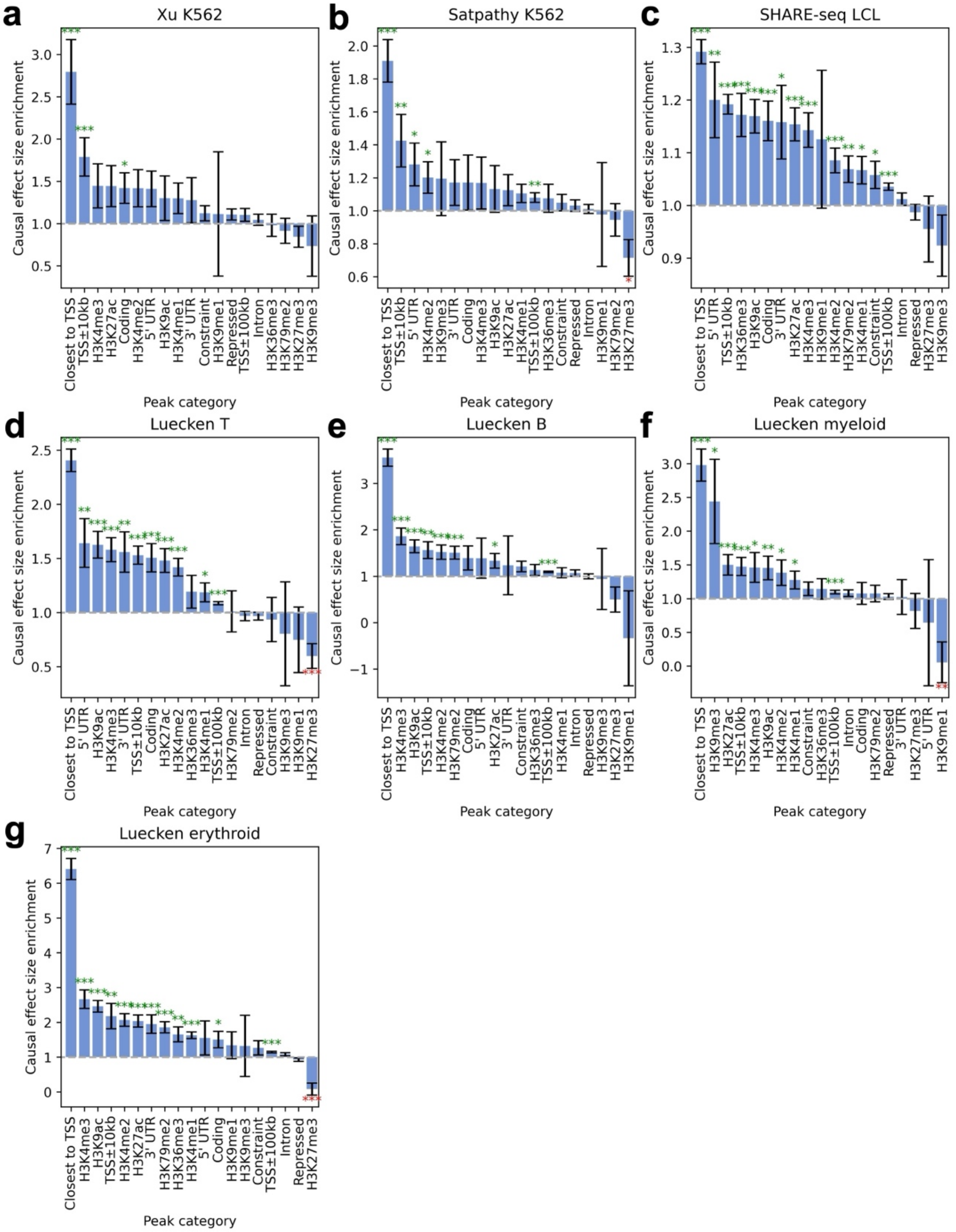


**Supplementary Figure 4. S-CASC causal effect size enrichments.** Causal effect size enrichment of 19 peak categories in stratified co-accessibility score regression in the **a)** Xu K562, **b)** Satpathy K562, **c)** SHARE-seq LCL, **d)** Luecken T, **e)** Luecken B, **f)** Luecken myeloid, and **g)** Luecken erythroid data set-cell type pairs. Bars and confidence intervals denote estimates and standard errors, respectively. Stars denote meta-analyzed p-values for significant enrichment (green) or depletion (red) (*: $p < 0.05$, **: $p < 0.01$, ***: $p < 0.001$). Numerical results are reported in **Supplementary Table 4**.

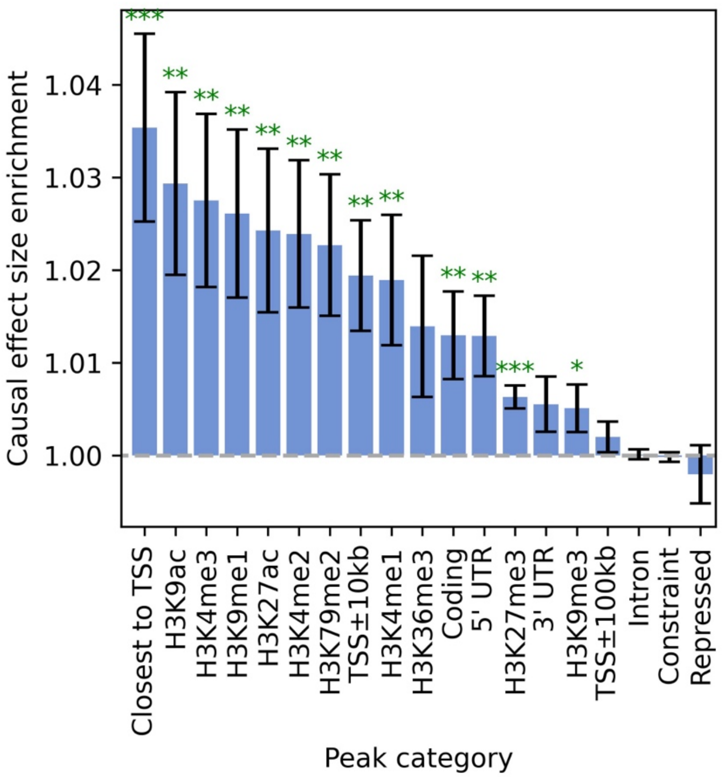


**Supplementary Figure 5. S-CASC causal effect size enrichments computed using correlations across single cells (instead of metacells).** Causal effect size enrichment of 19 peak categories in stratified co-accessibility score regression, implemented using peak-peak and peak-gene correlations computed across single cells (instead of metacells) and meta-analyzed across all 7 data set-cell type pairs. Results are qualitatively similar to **Figure 2c**, except for significant enrichments for the H3K27me3 and H3K9me1 categories (repressive histone marks) that were not observed at the metacell level. Bars and confidence intervals denote estimates and standard errors, respectively, meta-analyzed across data set-cell type pairs. Stars denote meta-analyzed p-values for significant enrichment (green) or depletion (red) (*: $p < 0.05$, **: $p < 0.01$, ***: $p < 0.001$).

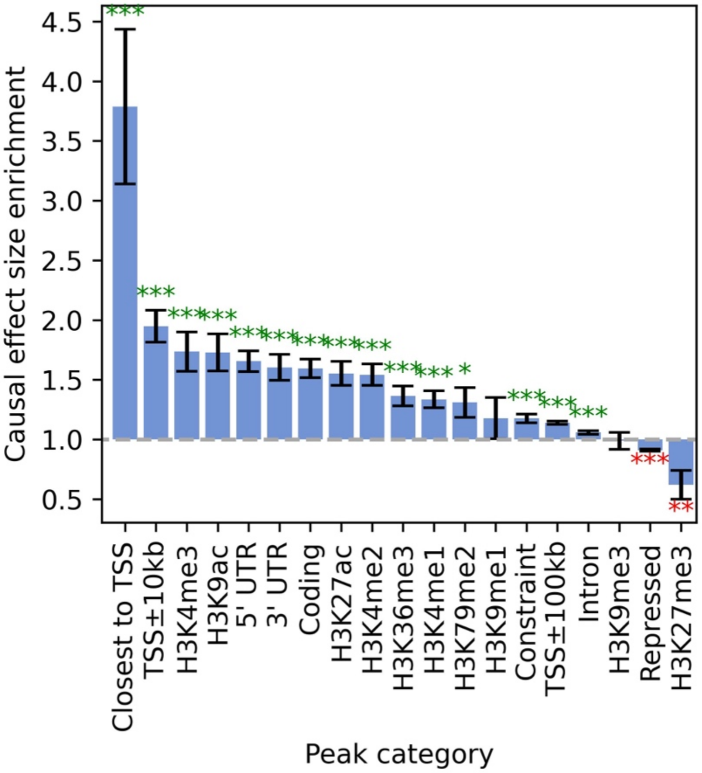


**Supplementary Figure 6. S-CASC causal effect size enrichments without conditioning on number of nearby genes.** Causal effect size enrichment of 19 peak categories in stratified co-accessibility score regression across peaks, implemented without the covariate for the number of genes <1Mb from each peak and meta-analyzed across all 7 data set-cell type pairs. Results are very similar to **Figure 2c**; differences include a small but significant enrichment for the "Intron" category and a small but significant depletion for the "Repressed" category (both nonsignificant in **Figure 2c**). Bars and confidence intervals denote estimates and standard errors, respectively, meta-analyzed across data set-cell type pairs. Stars denote meta-analyzed p-values for significant enrichment (green) or depletion (red) (*: $p < 0.05$, **: $p < 0.01$, ***: $p < 0.001$).

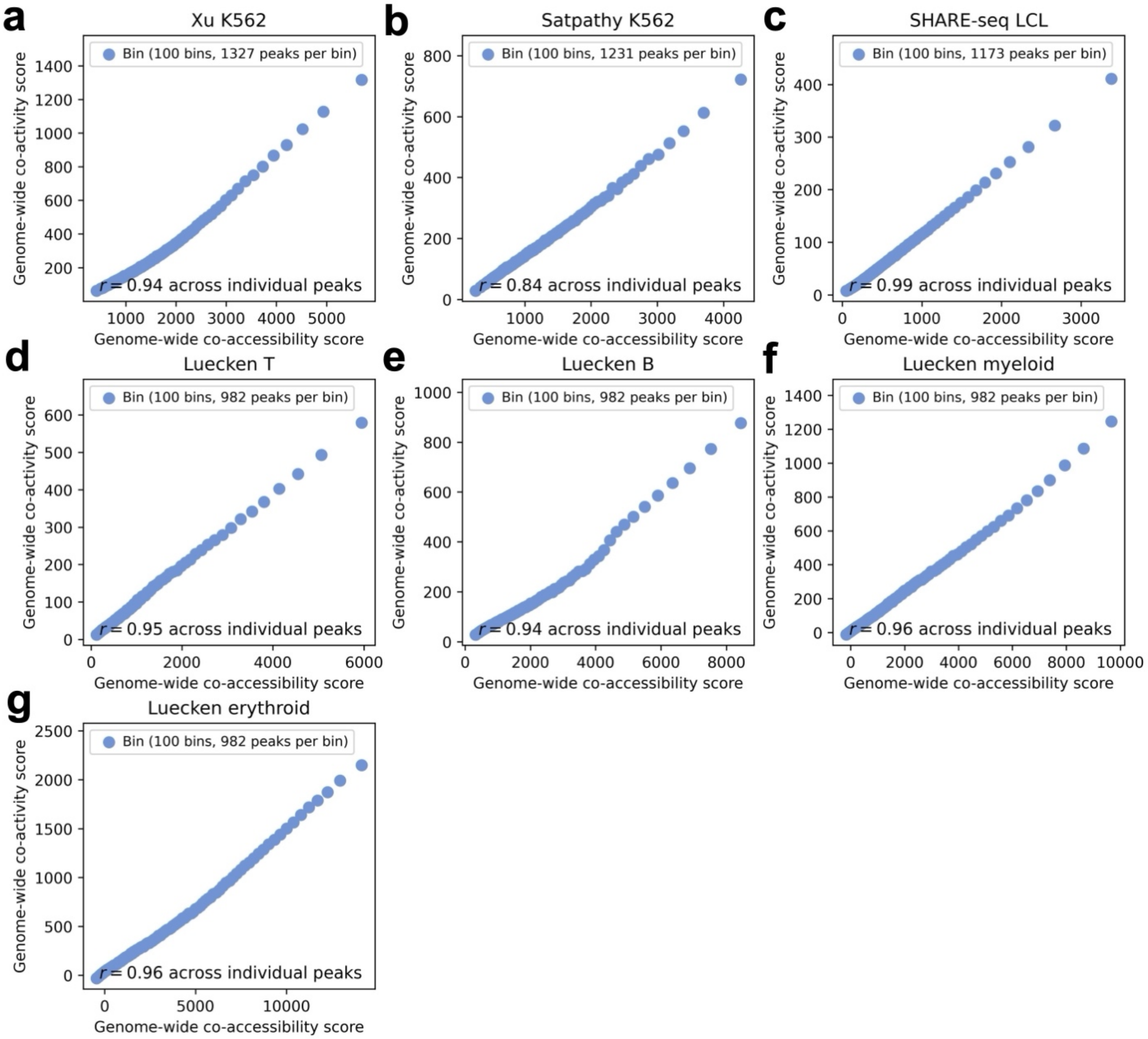


**Supplementary Figure 7. Relationship between genome-wide co-accessibility score and genome-wide co-activity score.** Relationship between genome-wide co-accessibility score and genome-wide co-activity score across peaks in the **a)** Xu K562, **b)** Satpathy K562, **c)** SHARE-seq LCL, **d)** Luecken T, **e)** Luecken B, **f)** Luecken myeloid, and **g)** Luecken erythroid data set-cell type pairs, computed using all peaks and genes genome-wide. Peaks are partitioned equally into 100 bins (each represented by 1 point) by co-accessibility score. Numerical results are reported in **Supplementary Table 5**.

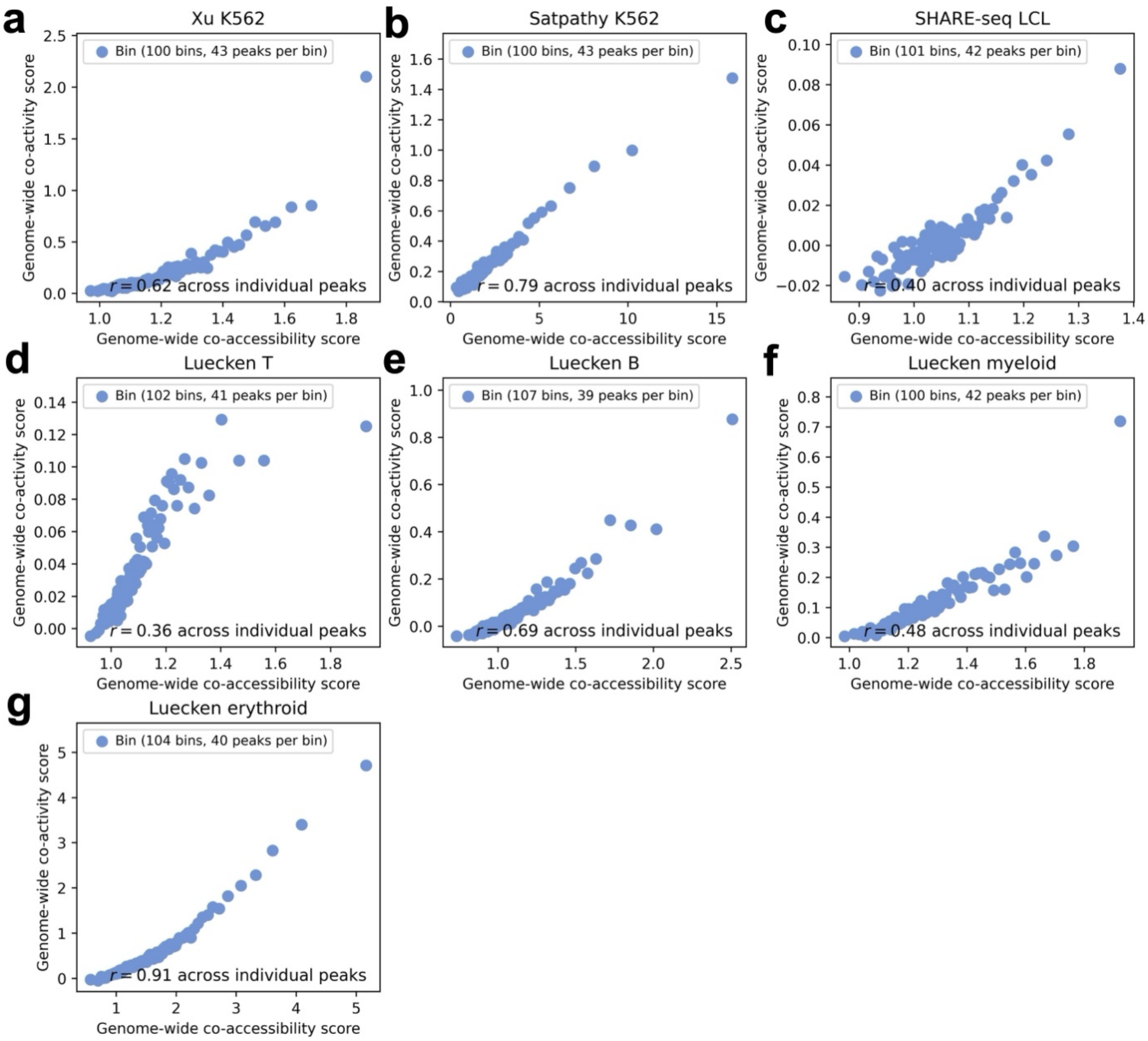


**Supplementary Figure 8. Relationship between genome-wide co-accessibility score and genome-wide co-activity score computed using correlations across single cells (instead of metacells).** Relationship between genome-wide co-accessibility score and genome-wide co-activity score (using a subset of 5000 peaks and 5000 genes) across peaks in the **a)** Xu K562, **b)** Satpathy K562, **c)** SHARE-seq LCL, **d)** Luecken T, **e)** Luecken B, **f)** Luecken myeloid, and **g)** Luecken erythroid data set-cell type pairs, computed using all peaks and genes genome-wide, with peak-peak and peak-gene correlations computed across single cells (instead of metacells). We downsampled to 5000 randomly selected peaks and 5000 randomly selected genes when computing scores (due to the high computational cost of computing correlations for every peak-peak, gene-gene, and peak-gene pair genome-wide across single cells). Peaks are partitioned equally into 100 bins (each represented by 1 point) by genome-wide co-accessibility score.

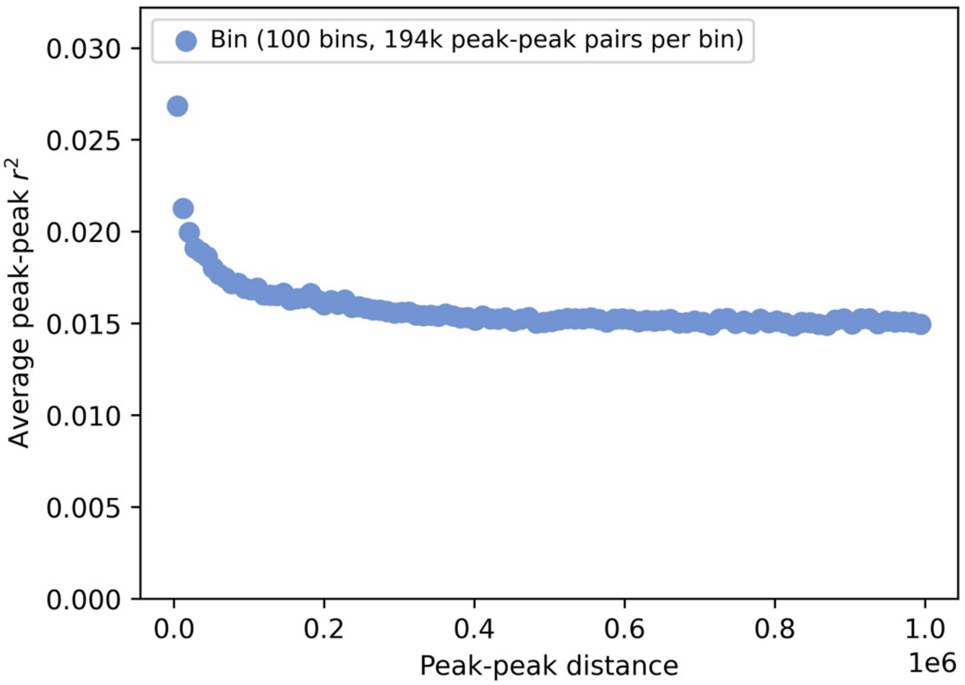


**Supplementary Figure 9. Average peak-peak $r^2$ at different peak-peak distances.** Relationship between average peak-peak squared correlation and peak-peak distance across 132,736 peaks in the Xu K562 data set. Peaks are partitioned equally into 100 bins (each represented by 1 point) by peak-peak distance. Peak-peak $r^2$ is moderately higher at shorter peak-peak distances, possibly implicating *cis* TF effects (the same copy of a TF binding to multiple nearby sites).

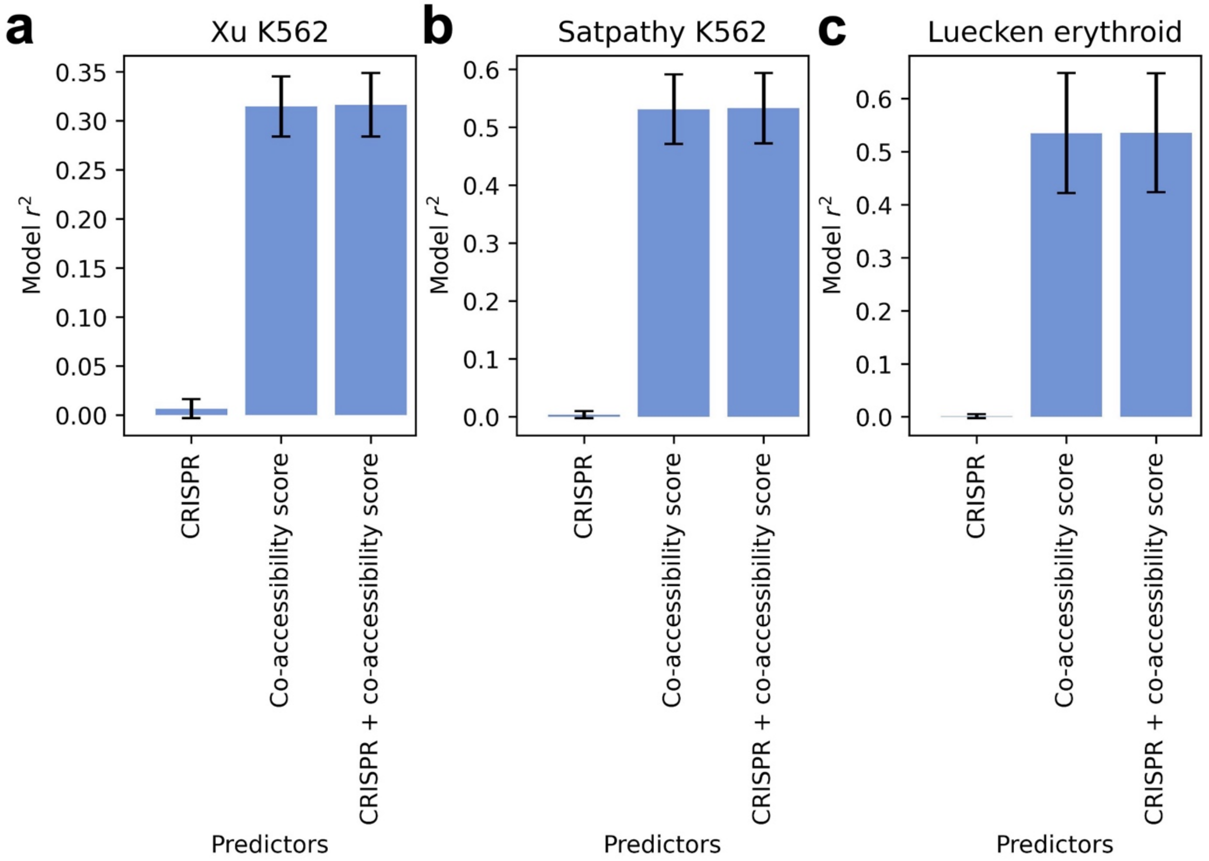


**Supplementary Figure 10. Regression of co-activity score on CRISPR status and co-accessibility score.** Model $r^2$ for linear regressions of co-activity score on CRISPR-positive status ("CRISPR"), co-accessibility score, or both, for the **a)** Xu K562, **b)** Satpathy K562, and **c)** Luecken erythroid data set-cell type pairs. Peaks in more 'active' regulatory regions (higher co-accessibility scores) may be more likely to truly affect gene expression. To assess the role of biological causality in driving the correlation between co-activity score and co-accessibility score, we performed a bivariate linear regression of co-activity score on co-accessibility score and a binary indicator of CRISPR-positive status (1 if a peak has at least one causal target gene validated by CRISPR, 0 otherwise) across peaks. We also performed univariate regressions of co-activity score on co-accessibility score and CRISPR-positive status individually. We restricted this analysis to the 3 data set-cell type pairs most relevant to K562 cells: Xu-K562, Satpathy-K562, and Luecken-erythroid. Within each data set-cell type pair, we further restricted to genes with ≥1 peak validated by CRISPR (CRISPR-positive) and ≥1 CRISPR-tested non-causal (CRISPR-negative) peak. Bars and confidence intervals denote estimates and standard errors, respectively.

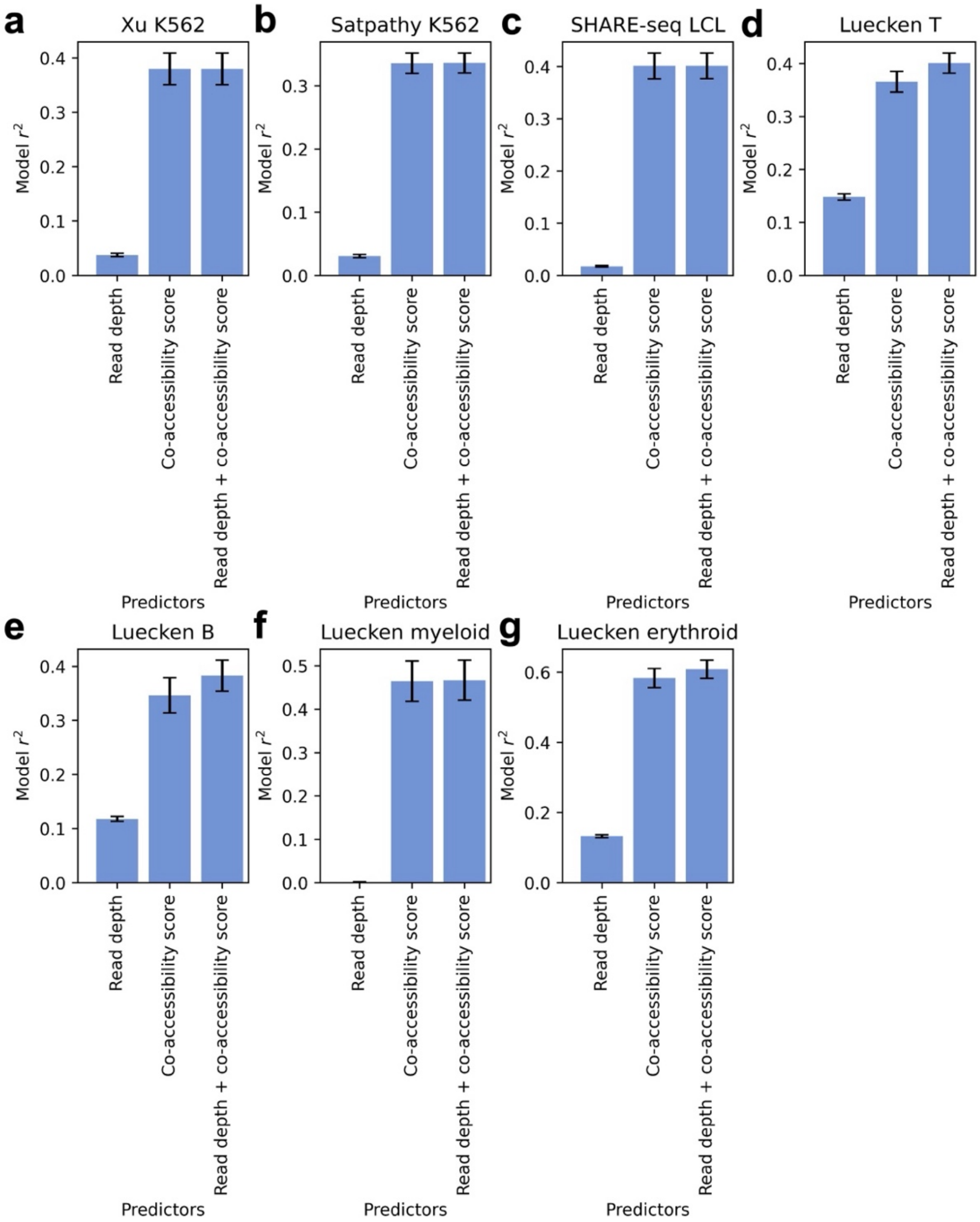


**Supplementary Figure 11**. **Regression of co-activity score on peak read depth and co-accessibility score.** Model $r^2$ for linear regressions of co-activity score on peak read depth, co-accessibility score, or both in the **a)** Xu K562, **b)** Satpathy K562, **c)** SHARE-seq LCL, **d)** Luecken T, **e)** Luecken B, **f)** Luecken myeloid, and **g)** Luecken erythroid data set-cell type pairs. Peaks with higher read depth may have higher measured correlations with other peaks and genes, generating higher co-accessibility scores and higher co-activity scores. To assess the role of read depth in driving the correlation between co-activity score and co-accessibility scores, we performed a bivariate linear regression of co-activity score on co-accessibility score and log(read depth) across peaks. We also performed univariate regressions of co-activity score on co-accessibility score and read depth individually. Bars and confidence intervals denote estimates and standard errors, respectively. (We note that log-transformed read depth attained higher $r^2$ than raw read depth.)

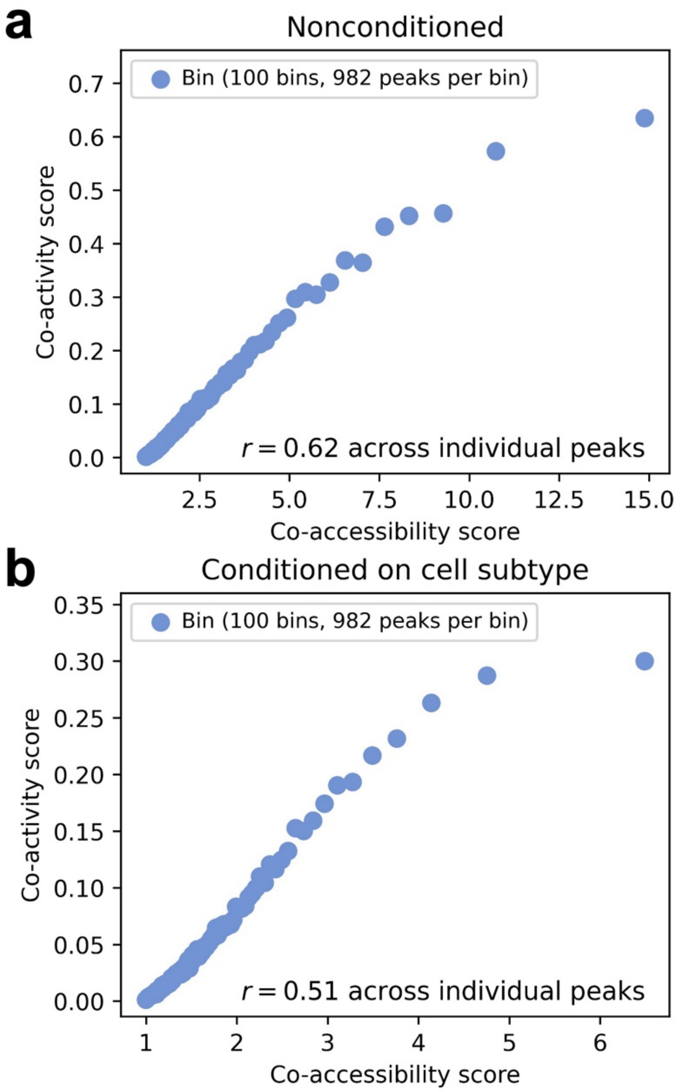


**Supplementary Figure 12**. **Relationship between co-accessibility score and co-activity score conditioned on cell subtype.** Relationship between co-accessibility score and co-activity score across peaks in the Luecken T cell data set computed using **a)** unconditioned peak-peak and peak-gene correlations and **b)** peak-peak and peak-gene correlations conditioned on cell subtype proportions. Cell subtypes may be associated with varying levels of average peak accessibility and gene expression, driving a correlation between co-accessibility score and co-activity score. To assess the role of cell subtypes, we analyzed T cells from the Luecken data set, which span 4 labeled T cell subtypes (activated CD4+, naive CD4+, CD8+, and naive CD8+; each represented by 1-12k cells). We separately residualized gene expression and ATAC peak accessibility by cell type proportions in each metacell (via linear regression across metacells), recomputed peak-peak correlations and peak-gene correlations using residualized data, and recomputed co-accessibility scores and co-activity scores by summing these conditional correlations. The correlation between co-activity score and co-accessibility score was lower after accounting for cell subtypes, suggesting that, while major cell subtypes do not fully explain the correlation, peak tagging may be related to cell subtype/state. Peaks are partitioned equally into 100 bins (each represented by 1 point) by co-accessibility score.

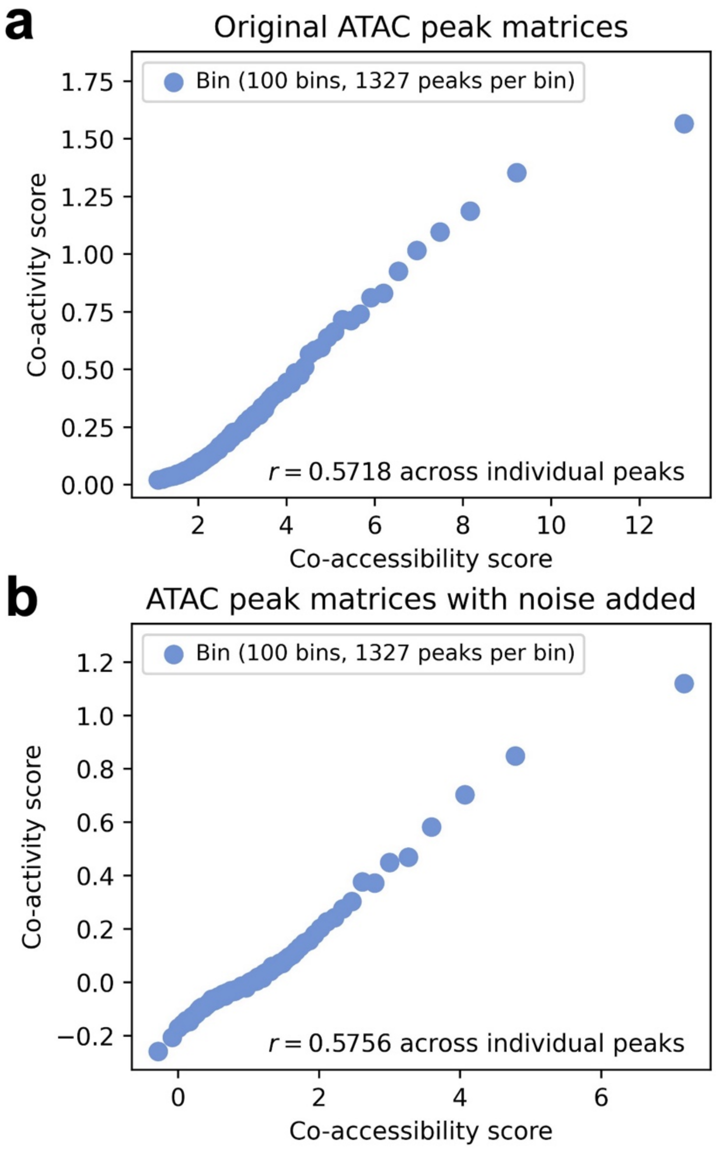


**Supplementary Figure 13**. **Relationship between co-accessibility score and co-activity score with ATAC measurement noise added.** Relationship between co-accessibility score and co-activity score across peaks in the Xu K562 data set computed using **a)** the original ATAC peak matrix and **b)** an ATAC peak matrix with added measurement noise. ATAC peaks with higher amounts of measurement noise may have lower measured correlations with other peaks and genes, possibly driving the correlation between co-activity score and co-accessibility score. Because it is difficult to directly assess the amount of measurement noise in ATAC data, we introduced additional artificial noise to 50% of ATAC peaks (by randomly setting 20% of entries equal to 0, retaining the original data for the remaining 50% of peaks), and recomputed co-accessibility and co-activity scores.

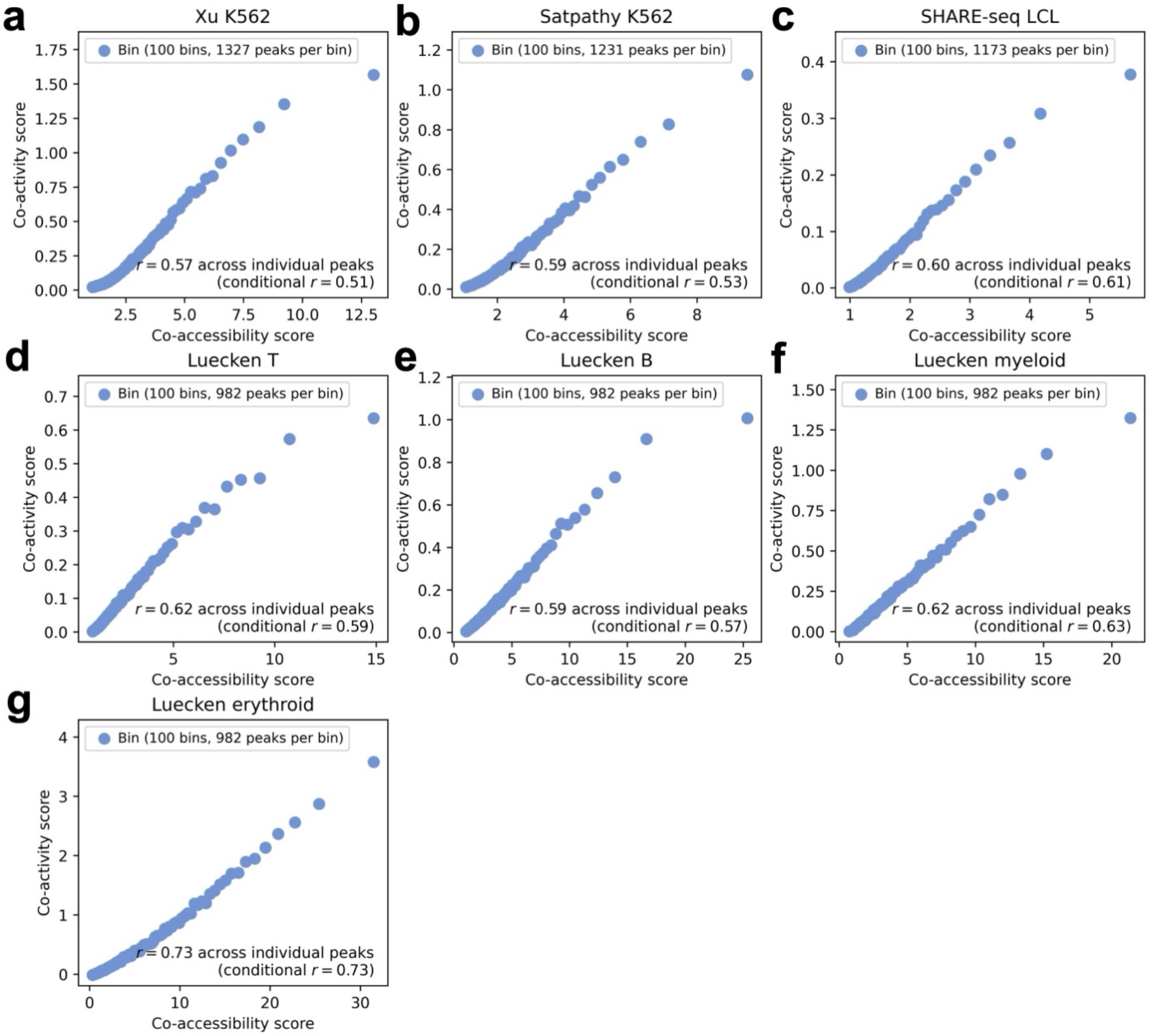


**Supplementary Figure 14. Relationship between co-accessibility score and co-activity score conditioned on number of nearby genes and peaks.** Relationship between co-accessibility score and co-activity score across peaks in the **a)** Xu K562, **b)** Satpathy K562, **c)** SHARE-seq LCL, **d)** Luecken T, **e)** Luecken B, **f)** Luecken myeloid, and **g)** Luecken erythroid data set-cell type pairs, computed using peaks and genes within the *cis* window (<1Mb) of each focal peak. 'Conditional *r*' denotes correlation conditioned on both the number of genes <1Mb from the focal peak and the number of peaks <1Mb from the focal peak (instead of only the number of genes <1Mb from the focal peak). Peaks are partitioned equally into 100 bins (each represented by 1 point) by co-accessibility score.

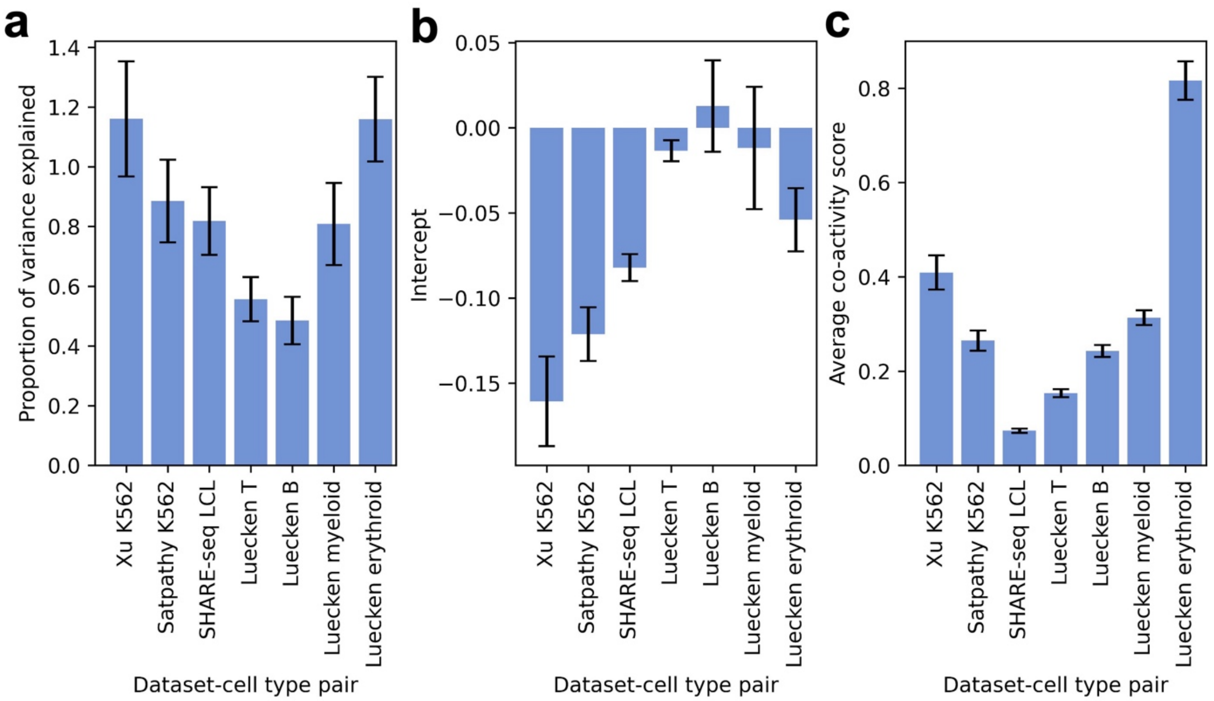


**Supplementary Figure 15. Estimates of proportion of gene expression variance explained by measured *cis*-peak accessibility from co-accessibility score regression. a)** Estimates of proportion of variance in gene expression explained by measured *cis*-peak accessibility, **b)** regression intercepts, and **c)** average co-activity score from co-accessibility score regression (performed without stratifying by functional category) in each data set-cell type pair. We regressed co-activity score on co-accessibility score across peaks (equivalent to performing stratified co-accessibility score regression with a single peak category defined by all peaks). We then estimated the proportion of variance explained by measured *cis*-peak accessibility as $\tau/k * M$, where $\tau$ is the slope of the regression, $k$ is the average number of genes <1Mb from the focal peak (across peaks), and $M$ is the average number of peaks <1Mb from the focal peak (across peaks). Estimating proportion of gene expression variance explained by peak accessibility is analogous to estimating heritability using LD score regression (LDSC)[28], with 2 changes: we divide by $k$ because co-activity score is the sum of $r^2$ across multiple genes instead of $r^2$ with a single trait as in ref.[28], and we do not divide by $N$ (number of metacells) because co-activity scores are sums of squared correlations and not $\chi^2$ statistics (which are equal to squared correlations multiplied by $N$) as in ref.[28] Importantly, we note that regression slopes may be overestimated when regression intercept is negative relative to the average value of co-activity score (leading to inflated estimates of proportion of gene expression variance explained); indeed, estimates in **a)** are largest for data set-cell type pairs with the most negative intercepts. The estimates in **a)** could be impacted by downward bias due to measurement noise or bias in either direction due to various forms of model misspecification, analogous to LDSC/S-LDSC[4,28,98]. Bars and confidence intervals denote estimates and standard errors, respectively.

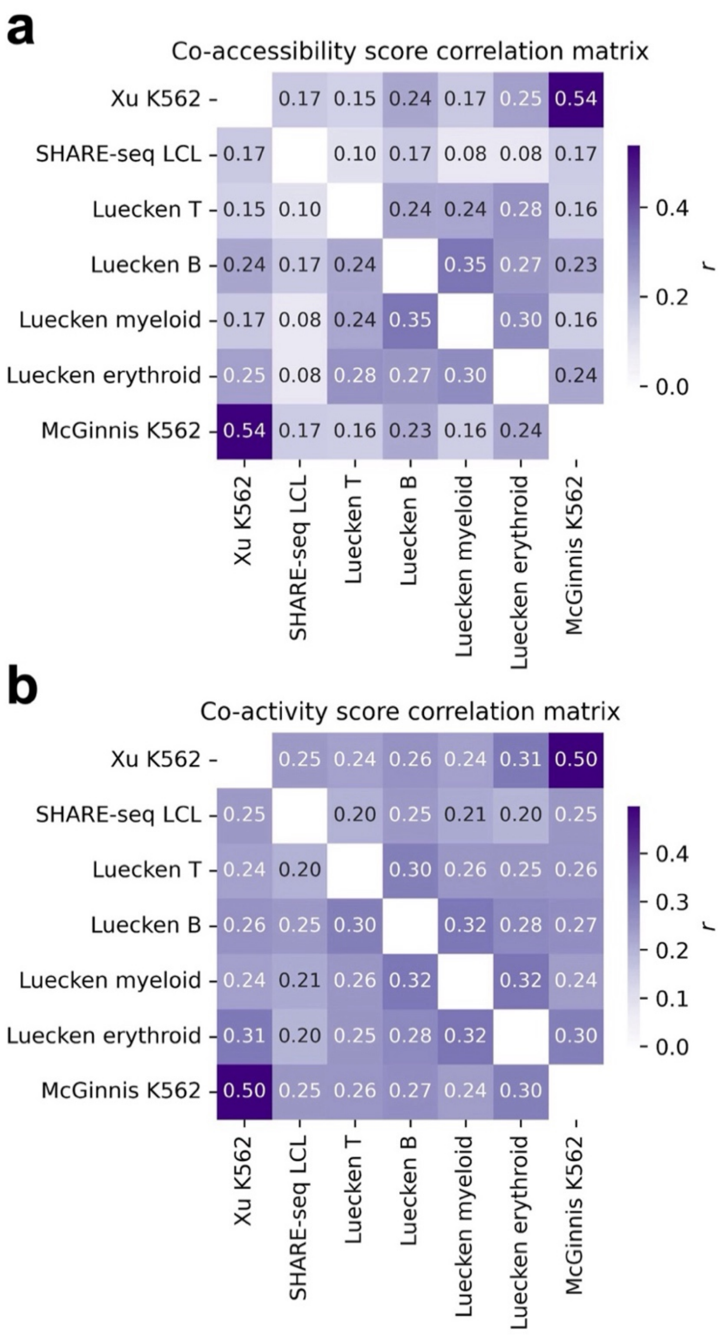


**Supplementary Figure 16. Correlations between co-accessibility scores and co-activity scores across data set-cell type pairs. a)** Pearson correlation matrix of co-accessibility scores computed in 7 data set-cell type pairs across ATAC peaks. **b)** Pearson correlation matrix of co-activity scores computed in 7 data set-cell type pairs across ATAC peaks. For each entry in **a)** and **b)** representing two data set-cell type pairs, correlation is computed across peaks called in both data set-cell type pairs (defined by any overlap between peaks in each data set-cell type pair). Scores tend to show stronger concordance in more similar cell types. For example: scores computed in K562 show the strongest concordance with each other, followed by scores computed in Luecken erythroid (the primary cell type most related to K562), and scores computed in SHARE-Seq LCL are more concordant with scores computed in Luecken B cells (the primary cell type from which LCL are derived) vs. other cell types.

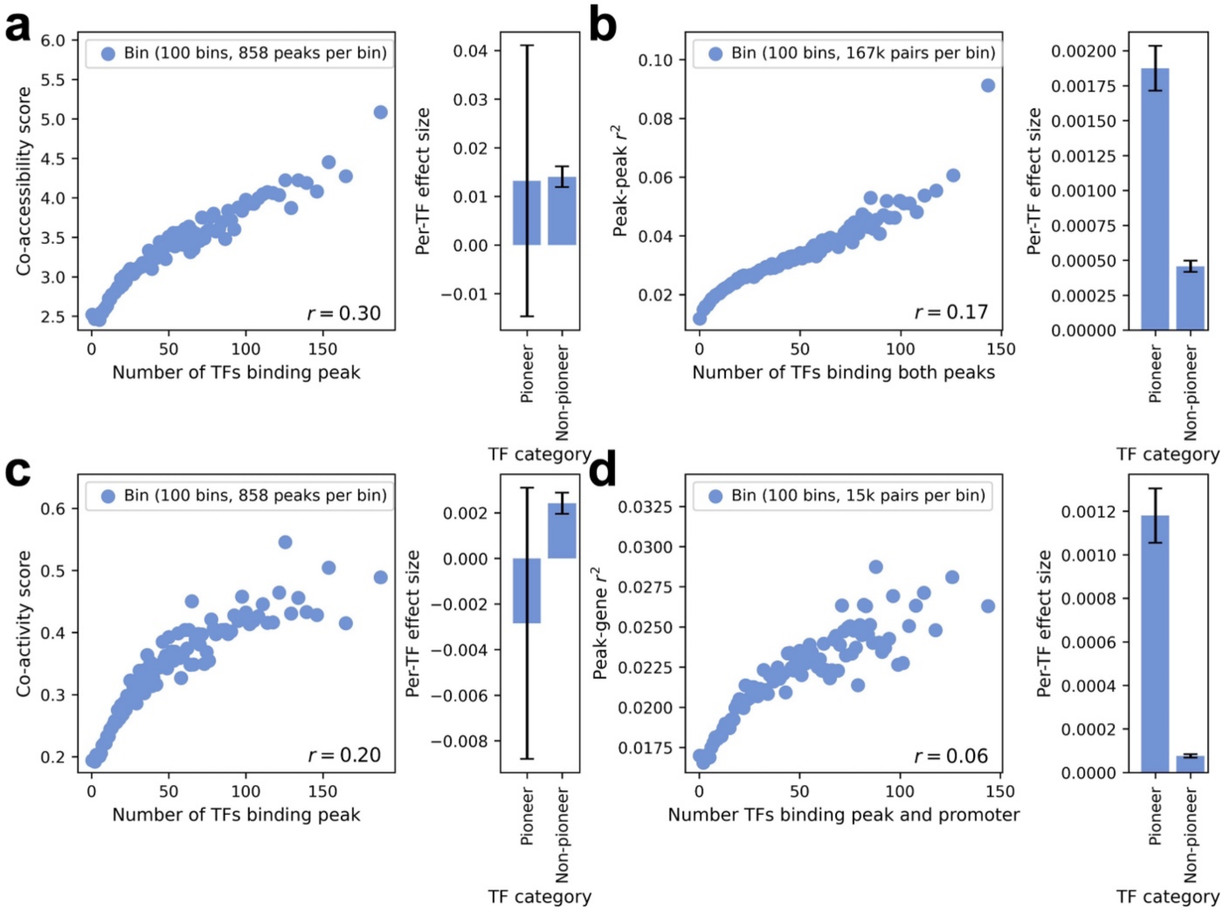


**Supplementary Figure 17. Relationship between peak-peak co-accessibility, peak-gene co-activity, and transcription factor binding activity in Satpathy K562 data set. a)** Relationship between number of TFs binding an ATAC peak and co-accessibility score (left). Per-TF effect for pioneer vs. non-pioneer TFs on co-accessibility score (right). **b)** Relationship between number of TFs binding both peaks in a peak-peak pair and squared peak-peak correlation (left). Per-TF effect for pioneer vs. non-pioneer TFs on squared peak-peak correlation (right). **c)** Relationship between number of TFs binding an ATAC peak and co-activity score (left). Per-TF effect for pioneer vs. non-pioneer TFs on co-activity score (right). **d)** Relationship between number of TFs binding both the peak and the gene (promoter) in a peak-gene pair and squared peak-gene correlation (left). Per-TF effect for pioneer vs. non-pioneer TFs on squared peak-gene correlation (right). Correlations and regression effect sizes are computed using 123,113 peaks in the Satpathy K562 data set. In left subpanels of **a-d**, peaks (or peak-peak / peak-gene pairs) are partitioned equally into 100 bins (each represented by 1 point) by x-axis value. In right subpanels of **a-d**, confidence intervals denote standard errors. Numerical results are reported in **Supplementary Table 8**.

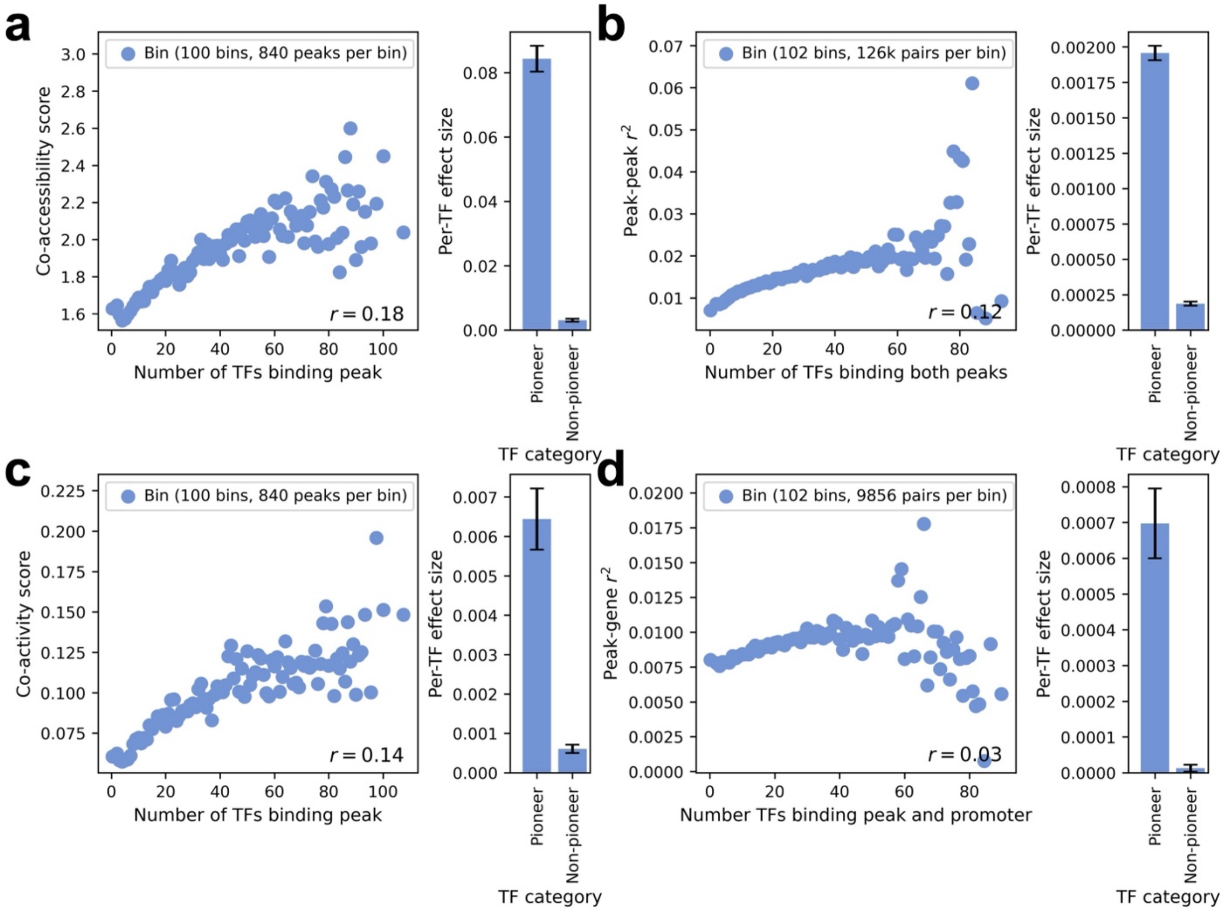


**Supplementary Figure 18. Relationship between peak-peak co-accessibility, peak-gene co-activity, and transcription factor binding activity in SHARE-seq LCL data set. a)** Relationship between number of TFs binding an ATAC peak and co-accessibility score (left). Per-TF effect for pioneer vs. non-pioneer TFs on co-accessibility score (right). **b)** Relationship between number of TFs binding both peaks in a peak-peak pair and squared peak-peak correlation (left). Per-TF effect for pioneer vs. non-pioneer TFs on squared peak-peak correlation (right). **c)** Relationship between number of TFs binding an ATAC peak and co-activity score (left). Per-TF effect for pioneer vs. non-pioneer TFs on co-activity score (right). **d)** Relationship between number of TFs binding both the peak and the gene (promoter) in a peak-gene pair and squared peak-gene correlation (left). Per-TF effect for pioneer vs. non-pioneer TFs on squared peak-gene correlation (right). Correlations and regression effect sizes are computed using 117,332 peaks in the SHARE-seq LCL data set. In left subpanels of **a-d**, peaks (or peak-peak / peak-gene pairs) are partitioned equally into 100 bins (each represented by 1 point) by x-axis value. In right subpanels of **a-d**, confidence intervals denote standard errors. Numerical results are reported in **Supplementary Table 8**.

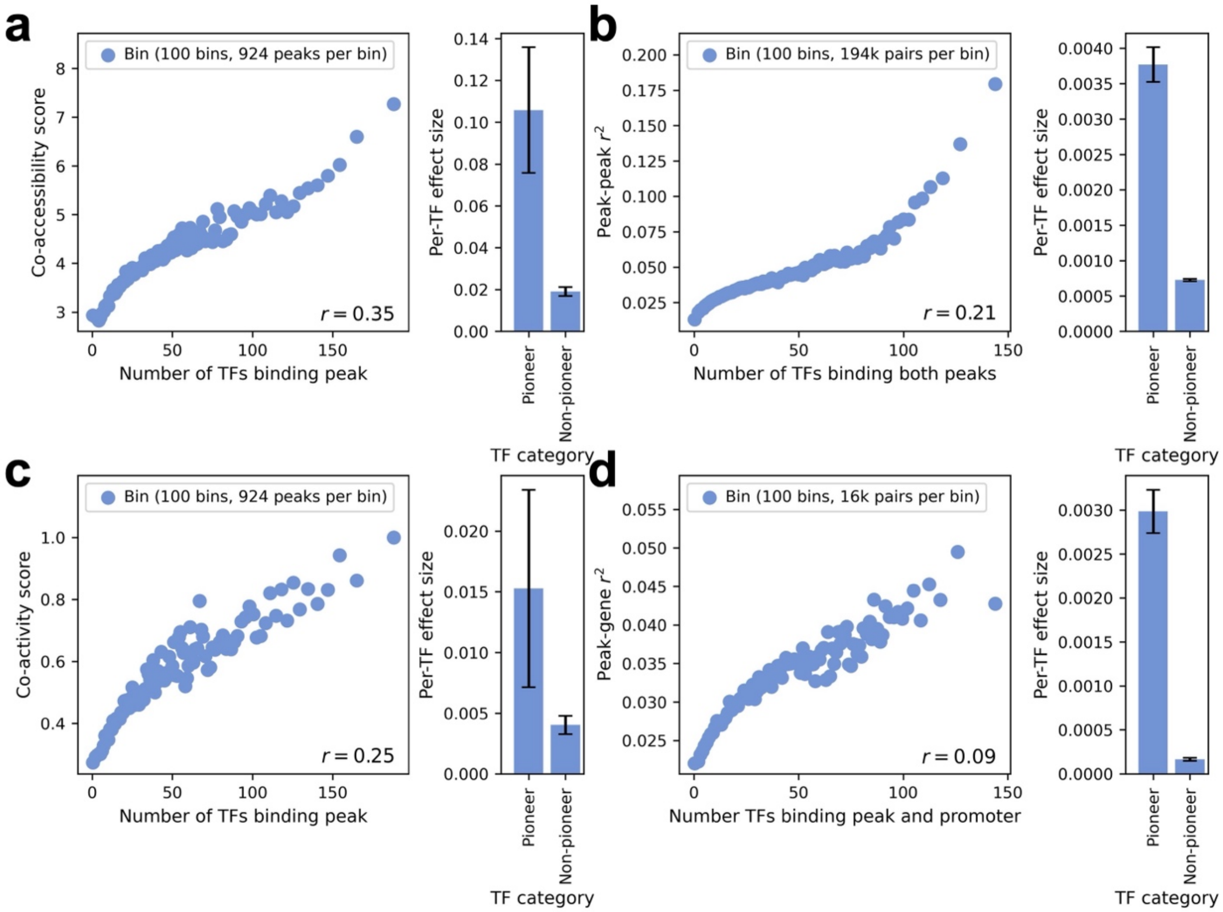


**Supplementary Figure 19. Relationship between peak-peak co-accessibility, peak-gene co-activity, and transcription factor binding activity in Xu K562 data set. a)** Relationship between number of TFs binding an ATAC peak and co-accessibility score (left). Per-TF effect for pioneer vs. non-pioneer TFs on co-accessibility score (right). **b)** Relationship between number of TFs binding both peaks in a peak-peak pair and squared peak-peak correlation (left). Per-TF effect for pioneer vs. non-pioneer TFs on squared peak-peak correlation (right). **c)** Relationship between number of TFs binding an ATAC peak and co-activity score (left). Per-TF effect for pioneer vs. non-pioneer TFs on co-activity score (right). **d)** Relationship between number of TFs binding both the peak and the gene (promoter) in a peak-gene pair and squared peak-gene correlation (left). Per-TF effect for pioneer vs. non-pioneer TFs on squared peak-gene correlation (right). Correlations and regression effect sizes are computed using 132,736 peaks in the Xu K562 data set. In left subpanels of **a-d**, peaks (or peak-peak / peak-gene pairs) are partitioned equally into 100 bins (each represented by 1 point) by x-axis value. In right subpanels of **a-d**, confidence intervals denote standard errors. Numerical results are reported in **Supplementary Table 8**.

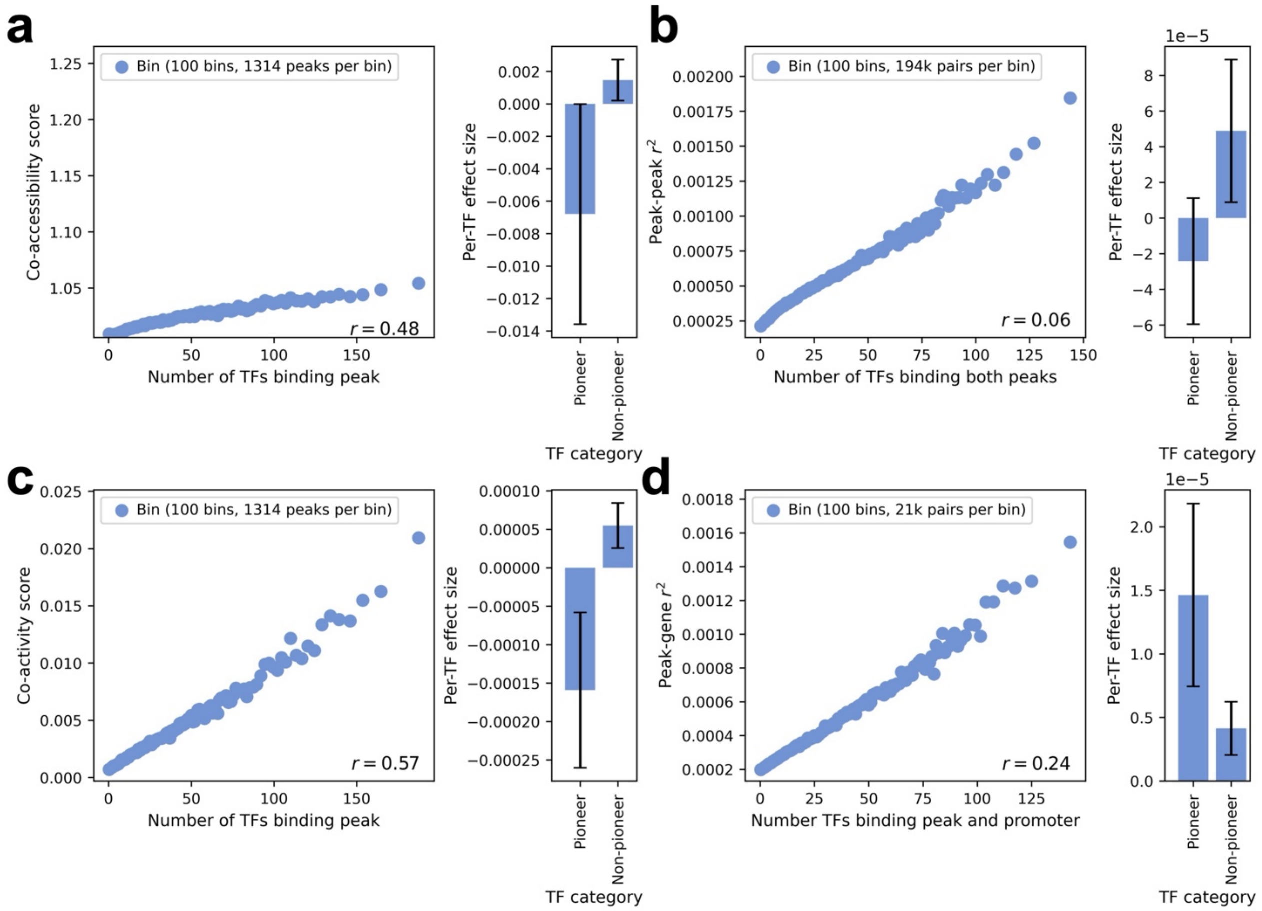


**Supplementary Figure 20. Relationship between peak-peak co-accessibility, peak-gene co-activity, and transcription factor binding activity using correlations across single cells (instead of metacells). a)** Relationship between number of TFs binding an ATAC peak and co-accessibility score (left). Per-TF effect for pioneer vs. non-pioneer TFs on co-accessibility score (right). **b)** Relationship between number of TFs binding both peaks in a peak-peak pair and squared peak-peak correlation (left). Per-TF effect for pioneer vs. non-pioneer TFs on squared peak-peak correlation (rit). **c)** Relationship between number of TFs binding an ATAC peak and co-activity score (left). Per-TF effect for pioneer vs. non-pioneer TFs on co-activity score (right). **d)** Relationship between number of TFs binding both the peak and the gene (promoter) in a peak-gene pair and squared peak-gene correlation (left). Per-TF effect for pioneer vs. non-pioneer TFs on squared peak-gene correlation (right). All peak-peak and peak-gene correlations are computed across single cells (instead of metacells). Analyses comparing per-TF effect size for pioneer vs. non-pioneer TFs (right subpanels of **a-d**) did not yield statistically significant differences (each p-value > 0.05). In left subpanels of **a-d**, correlations are computed across 132,736 peaks in the Xu K562 data set, and peaks (or peak-peak / peak-gene pairs) are partitioned equally into 100 bins (each represented by 1 point) by x-axis value. In right subpanels of **a-d**, regression effect sizes are meta-analyzed across 3 data set-cell type pairs (Xu K562, Satpathy-K562, and SHARE-seq-LCL), and confidence intervals denote standard errors.

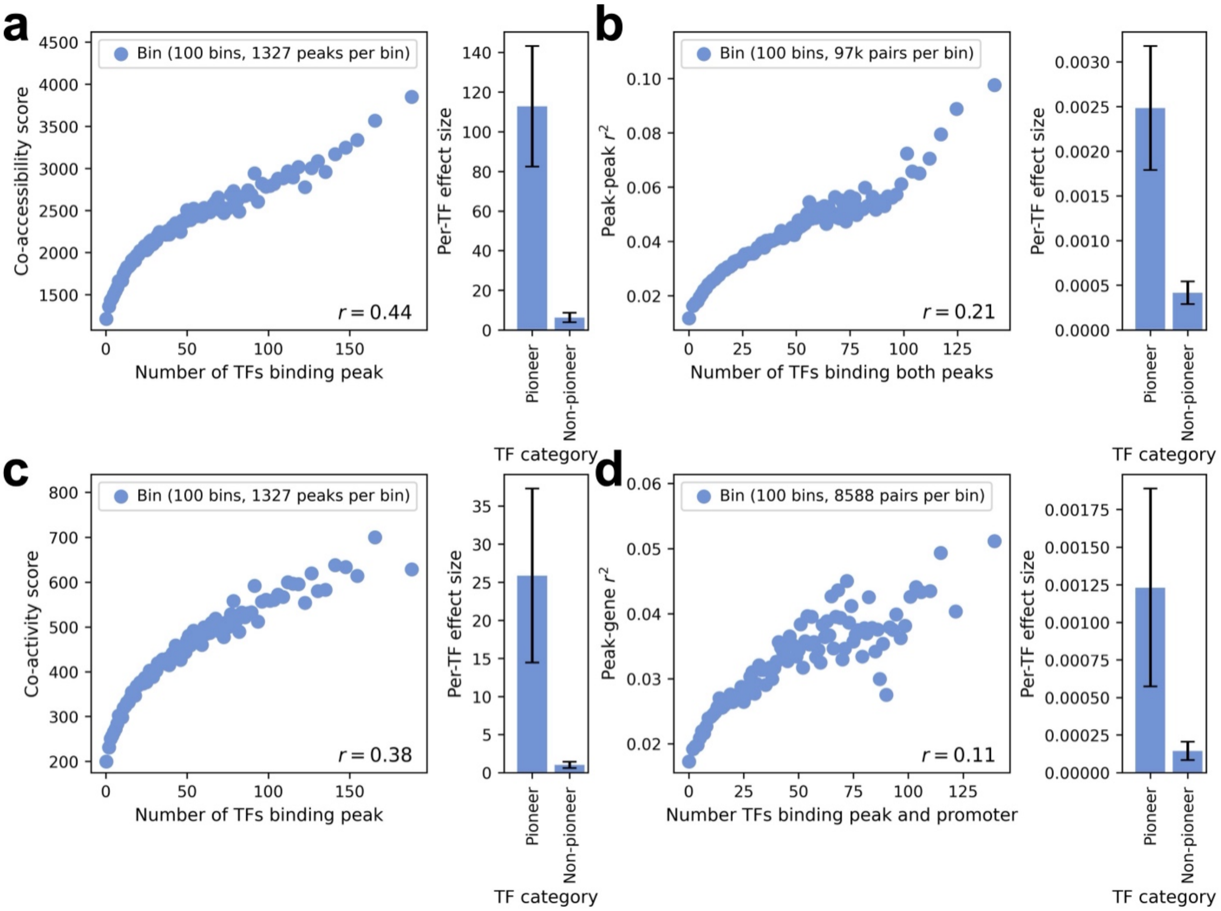


**Supplementary Figure 21. Relationship between genome-wide peak-peak co-accessibility, genome-wide peak-gene co-activity, and transcription factor binding activity. a)** Relationship between number of TFs binding an ATAC peak and genome-wide co-accessibility score (left). Per-TF effect for pioneer vs. non-pioneer TFs on genome-wide co-accessibility score (p-value for difference = 0.0002) (right). **b)** Relationship between number of TFs binding both peaks in a peak-peak pair and squared peak-peak correlation, across all peak-peak pairs (not restricted by peak-peak distance) from a random subset of 5,000 ATAC peaks (left). Per-TF effect for pioneer vs. non-pioneer TFs on squared peak-peak correlation, across all peak-peak pairs (not restricted by peak-peak distance) from a random subset of 5,000 ATAC peaks (p-value for difference = 0.0003) (right). **c)** Relationship between number of TFs binding an ATAC peak and genome-wide co-activity score (left). Per-TF effect for pioneer vs. non-pioneer TFs on genome-wide co-activity score (p-value for difference = 0.024) (right). **d)** Relationship between number of TFs binding both the peak and the gene (promoter) in a peak-gene pair and squared peak-gene correlation, across all peak-gene pairs (not restricted by peak-gene distance) from a random subset of 5,000 ATAC peaks and 5,000 genes (left). Per-TF effect for pioneer vs. non-pioneer TFs on squared peak-gene correlation, across all peak-gene pairs (not restricted by peak-gene distance) from a random subset of 5,000 ATAC peaks and 5,000 genes (p-value for difference = 0.070) (right). In left subpanels of **a-d**, correlations are computed across 132,736 peaks in the Xu K562 data set, and peaks (or peak-peak / peak-gene pairs) are partitioned equally into 100 bins (each represented by 1 point) by x-axis value. In right subpanels of **a-d**, regression effect sizes are meta-analyzed across 3 data set-cell type pairs (Xu K562, Satpathy-K562, and SHARE-seq-LCL), and confidence intervals denote standard errors.

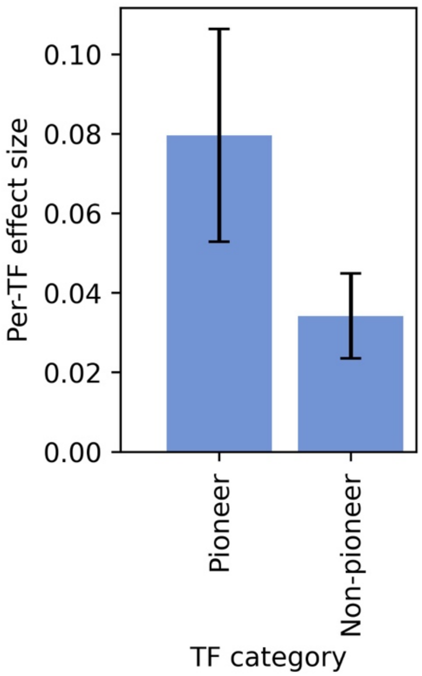


**Supplementary Figure 22. Pioneer vs. non-pioneer TF effects after conditioning on TF expression and binding.** Per-TF effect for pioneer vs. non-pioneer TFs on co-accessibility score in the Xu K562 data set after conditioning on overall TF expression and binding (p-value for difference = 0.034). We partitioned TFs equally into 10 bins by total expression (defined as expression of the focal TF in Xu K562 RNA-seq data) and 10 bins by total binding (defined as number of peaks in the Xu K562 data bound by the focal TF). We performed a linear regression of co-accessibility score on: 1 predictor for the number of pioneer TFs binding in a peak, 1 predictor for the number of non-pioneer TFs binding in a peak, 10 predictors for the number of TFs in each expression bin binding in a peak, and 10 predictors for the number of TFs in each binding bin binding in a peak. Plotted are the effect sizes for pioneer and non-pioneer TFs (conditioned on TF expression and binding information). Confidence intervals denote standard errors.

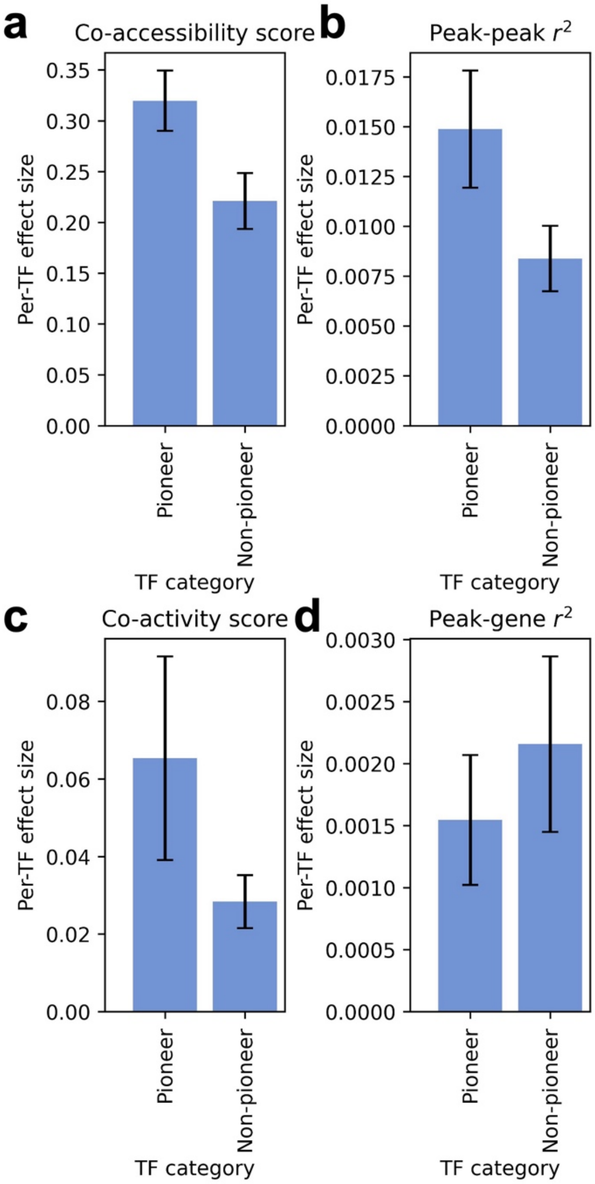


**Supplementary Figure 23. Pioneer vs. non-pioneer TF effects using Perturb-multiome data to define pioneer TFs.** Per-TF effect for pioneer vs. non-pioneer TFs on **a)** co-accessibility score (p-value for difference = 4e-12), **b)** squared peak-peak correlation (p-value for difference = 8e-7), **c)** co-activity score (p-value for difference = 0.067), **d)** squared peak-gene correlation (p-value for difference = 0.61). We used Perturb-multiome data from ref.[48] to define pioneer TFs as TFs whose perturbation is associated with a significant change in peak accessibility or gene expression in scRNA+ATAC-seq multiome data and repeated each bivariate regression comparing pioneer vs. non-pioneer TF effects (right panels of **Figure 3a-d**). Confidence intervals denote standard errors. Regression effect sizes are meta-analyzed across 2 data set-cell type pairs most similar to the HSPCs assayed in ref.[48] (Xu K562 and Satpathy K562). Bars and confidence intervals denote estimates and standard errors, respectively.

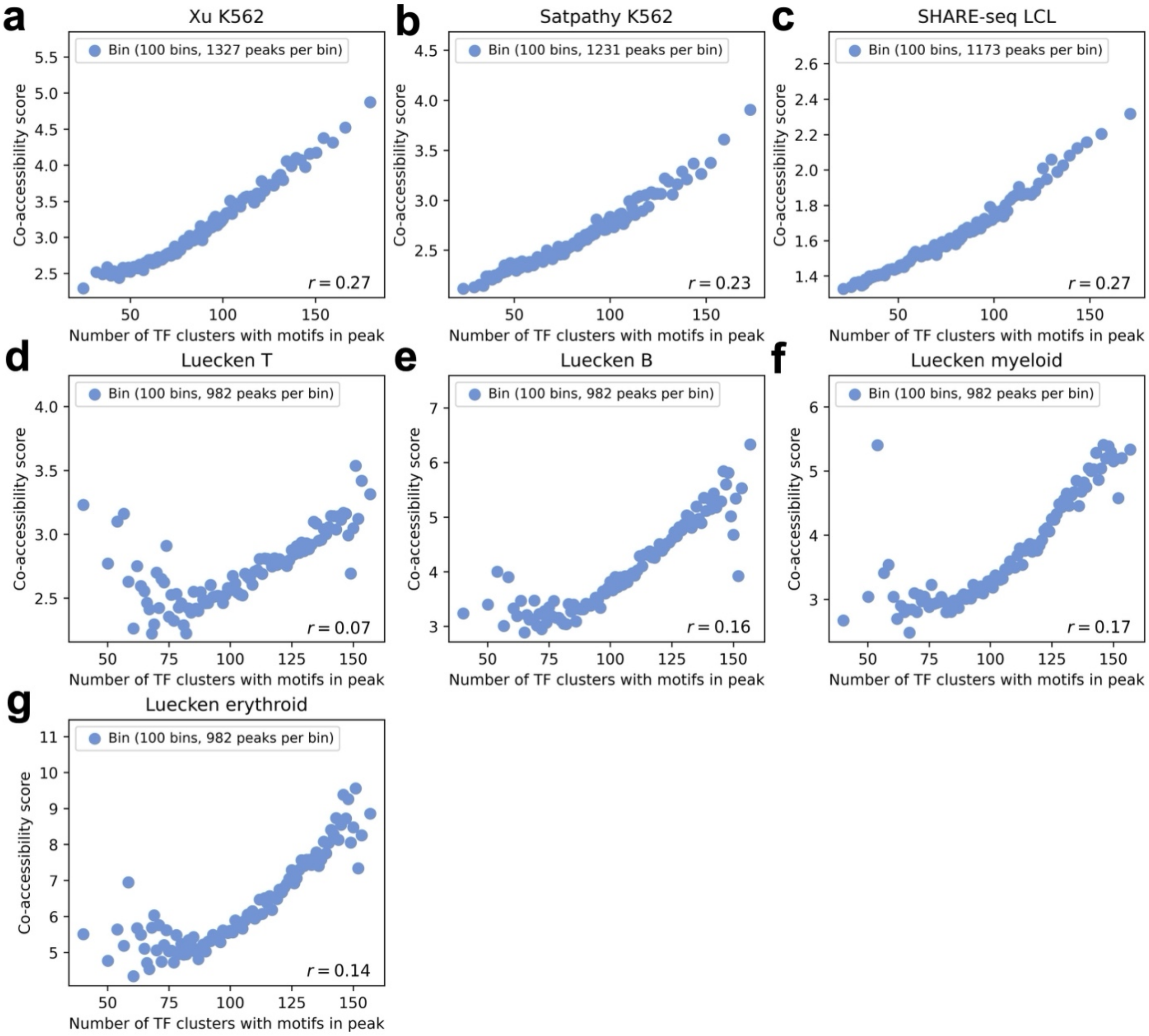


**Supplementary Figure 24. Relationship between co-accessibility score and transcription factor binding activity as assessed by transcription factor motifs. a)** Relationship between number of TF clusters with DNA sequence motifs in an ATAC peak and co-accessibility score across peaks in the **a)** Xu K562, **b)** Satpathy K562, **c)** SHARE-seq LCL, **d)** Luecken T, **e)** Luecken B, **f)** Luecken myeloid, and **g)** Luecken erythroid data set-cell type pairs. Peaks are partitioned equally into 100 bins (each represented by 1 point) by x-axis value. We obtained genomic coordinates for TF binding motifs (sites with predicted TF binding activity based on DNA sequence) from ref.[49], which grouped 2,179 TFs into 286 TF clusters with highly similar binding motifs. For a given TF cluster and data set-cell type pair, we computed the number of TF clusters with motifs in each ATAC peak (analogous to ChIP-seq analyses, using sequence motifs instead of ChIP-seq peaks to define genomic regions of TF activity).

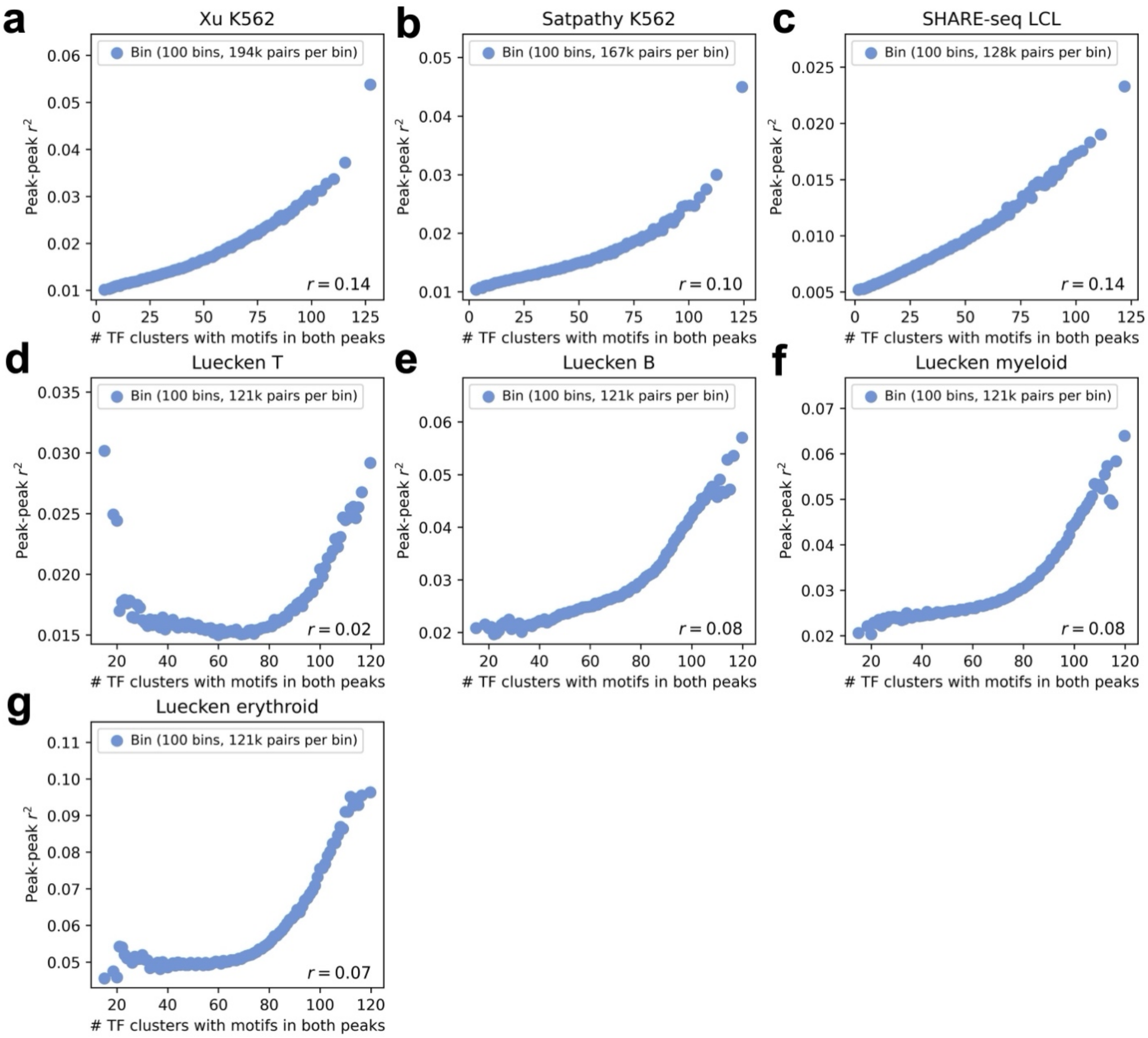


**Supplementary Figure 25. Relationship between squared peak-peak correlation and transcription factor binding activity as assessed by transcription factor motifs. a)** Relationship between number of TF clusters with DNA sequence motifs in both ATAC peaks in a peak-peak pair and peak-peak $r^2$, across peak-peak pairs in the **a)** Xu K562, **b)** Satpathy K562, **c)** SHARE-seq LCL, **d)** Luecken T, **e)** Luecken B, **f)** Luecken myeloid, and **g)** Luecken erythroid data set-cell type pairs. Peak-peak pairs are partitioned equally into 100 bins (each represented by 1 point) by x-axis value. We obtained genomic coordinates for TF binding motifs (sites with predicted TF binding activity based on DNA sequence) from ref.[49], which grouped 2,179 TFs into 286 TF clusters with highly similar binding motifs. For a given TF cluster and data set-cell type pair, we computed the number of TF clusters with motifs in each ATAC peak. For each peak-peak pair with peak-peak distance <1Mb, we then computed the number of TF clusters with motifs in both peaks (analogous to ChIP-seq analyses, using sequence motifs instead of ChIP-seq peaks to define genomic regions of TF activity).

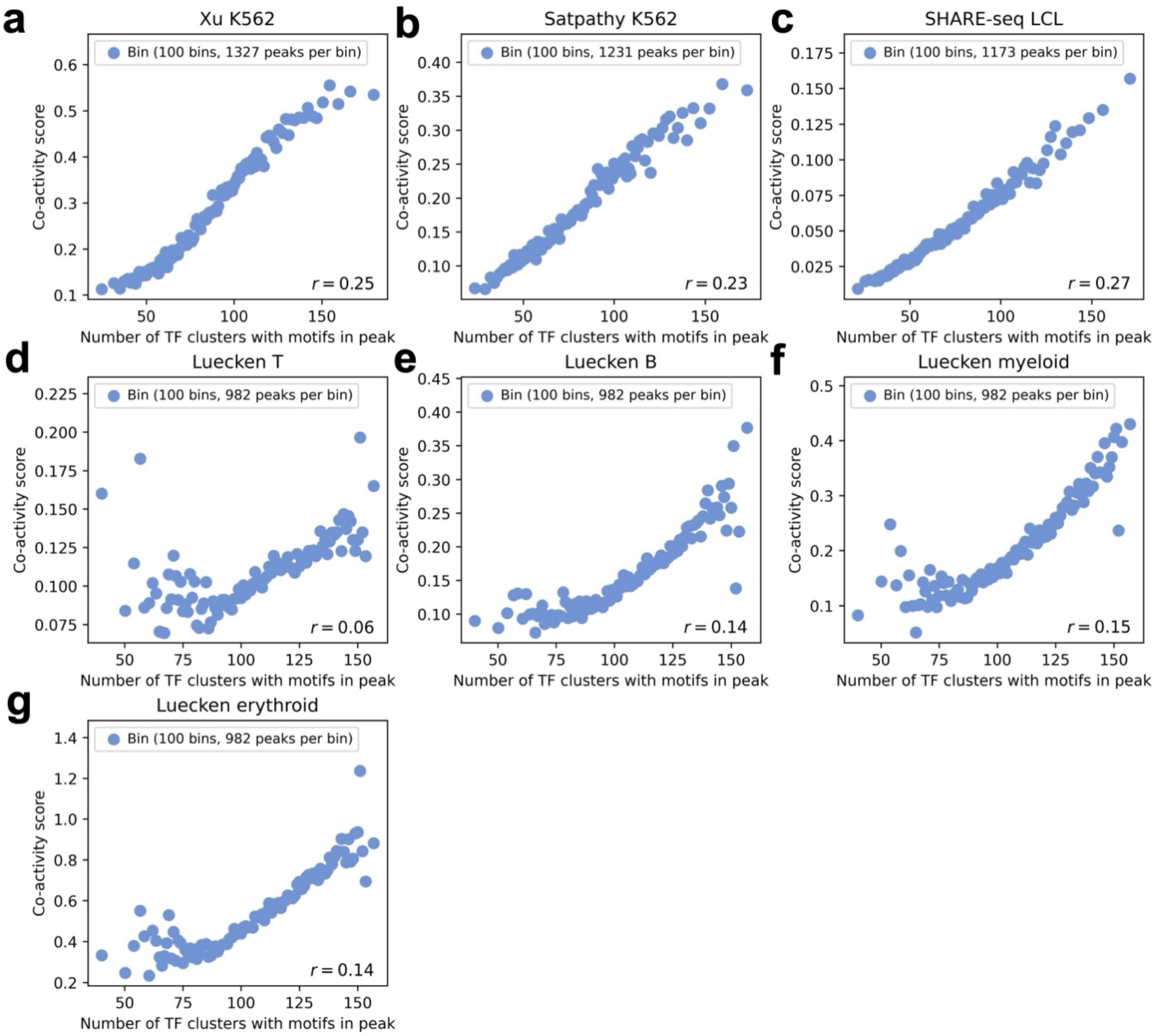


**Supplementary Figure 26. Relationship between co-activity score and transcription factor binding activity as assessed by transcription factor motifs. a)** Relationship between number of TF clusters with DNA sequence motifs in an ATAC peak and co-activity score across peaks in the **a)** Xu K562, **b)** Satpathy K562, **c)** SHARE-seq LCL, **d)** Luecken T, **e)** Luecken B, **f)** Luecken myeloid, and **g)** Luecken erythroid data set-cell type pairs. Peaks are partitioned equally into 100 bins (each represented by 1 point) by x-axis value. We obtained genomic coordinates for TF binding motifs (sites with predicted TF binding activity based on DNA sequence) from ref.[49], which grouped 2,179 TFs into 286 TF clusters with highly similar binding motifs. For a given TF cluster and data set-cell type pair, we computed the number of TF clusters with motifs in each ATAC peak and each gene's promoter (analogous to ChIP-seq analyses, using sequence motifs instead of ChIP-seq peaks to define genomic regions of TF activity).

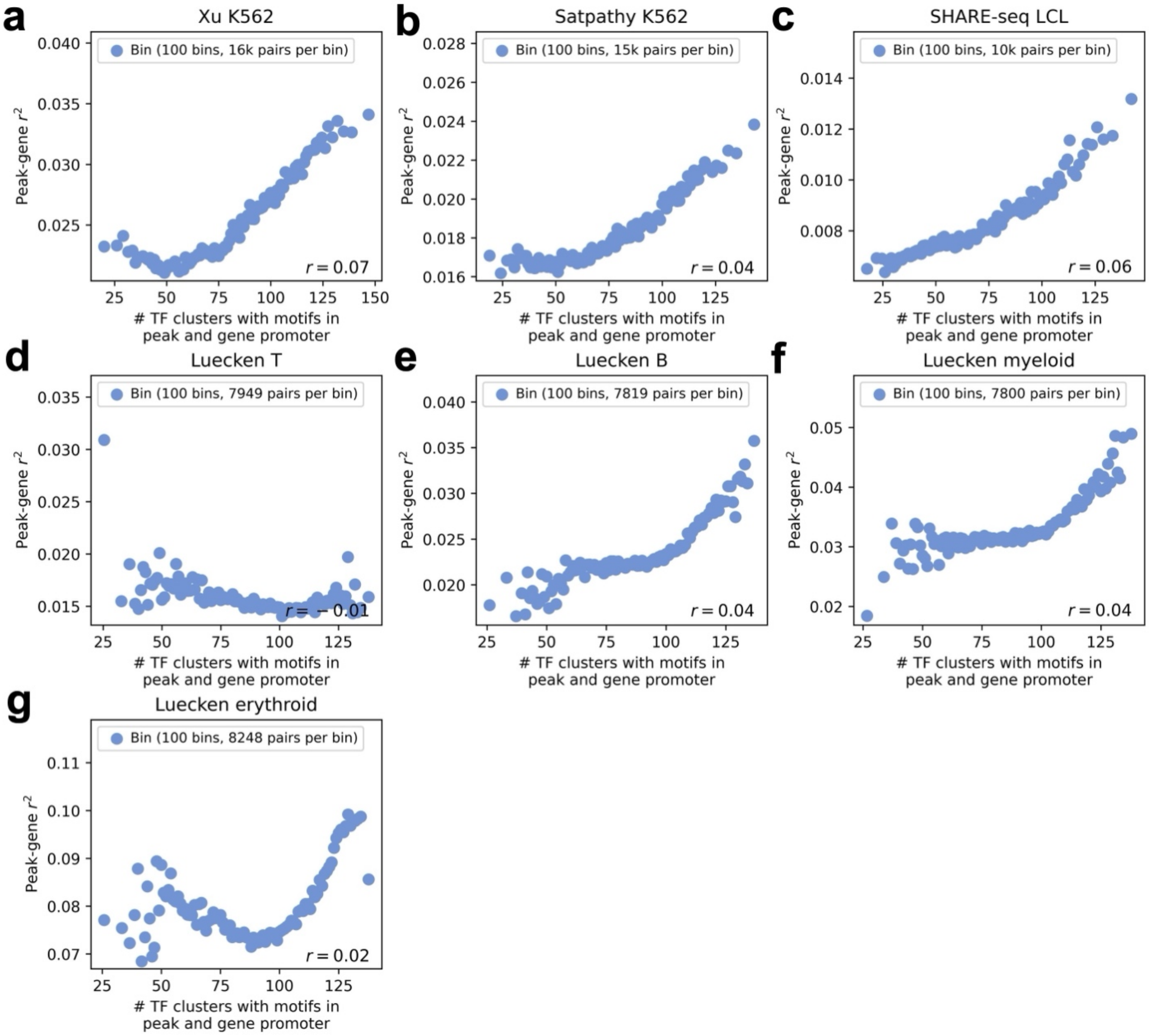


**Supplementary Figure 27. Relationship between squared peak-gene correlation and transcription factor binding activity as assessed by transcription factor motifs. a)** Relationship between number of TF clusters with DNA sequence motifs in both the ATAC peak and the gene's promoter for a peak-gene pair and peak-gene $r^2$, across peak-gene pairs in the **a)** Xu K562, **b)** Satpathy K562, **c)** SHARE-seq LCL, **d)** Luecken T, **e)** Luecken B, **f)** Luecken myeloid, and **g)** Luecken erythroid data set-cell type pairs. Peak-gene pairs are partitioned equally into 100 bins (each represented by 1 point) by x-axis value. We obtained genomic coordinates for TF binding motifs (sites with predicted TF binding activity based on DNA sequence) from ref.[49], which grouped 2,179 TFs into 286 TF clusters with highly similar binding motifs. For a given TF cluster and data set-cell type pair, we computed the number of TF clusters with motifs in each ATAC peak and each gene's promoter. For each peak-gene pair with peak-gene distance <1Mb, we then computed the number of TF clusters with motifs in both the peak and gene's promoter (analogous to ChIP-seq analyses, using sequence motifs instead of ChIP-seq peaks to define genomic regions of TF activity).

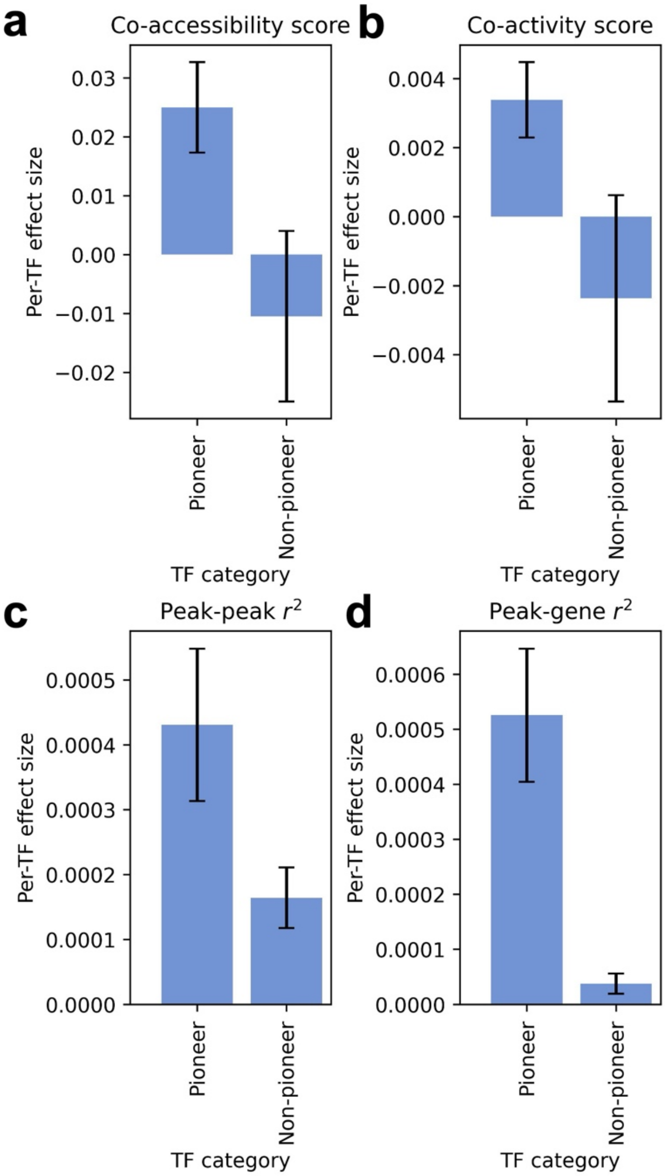


**Supplementary Figure 28. Pioneer vs. non-pioneer TF effects using transcription factor binding activity as assessed by transcription factor motifs.** Per-TF effect for pioneer vs. non-pioneer TF motifs on **a)** co-accessibility score (p-value for difference = 4e-12), **b)** squared peak-peak correlation (p-value for difference = 8e-7), **c)** co-activity score (p-value for difference = 0.067), **d)** squared peak-gene correlation (p-value for difference = 0.61). We obtained genomic coordinates for TF binding motifs (sites with predicted TF binding activity based on DNA sequence) from ref.[49], which grouped 2,179 TFs into 286 TF clusters with highly similar binding motifs. For a given TF cluster and data set-cell type pair, we computed the number of TF clusters with motifs in each ATAC peak and each gene's promoter. For each peak-gene pair with peak-gene distance <1Mb, we then computed the number of TF clusters with motifs in both the peak and gene's promoter (analogous to ChIP-seq analyses, using sequence motifs instead of ChIP-seq peaks to define genomic regions of TF activity). We defined pioneer TF motif clusters as clusters containing a TF from our main set of pioneer TFs (obtained from refs.[46,47]). Confidence intervals denote standard errors.

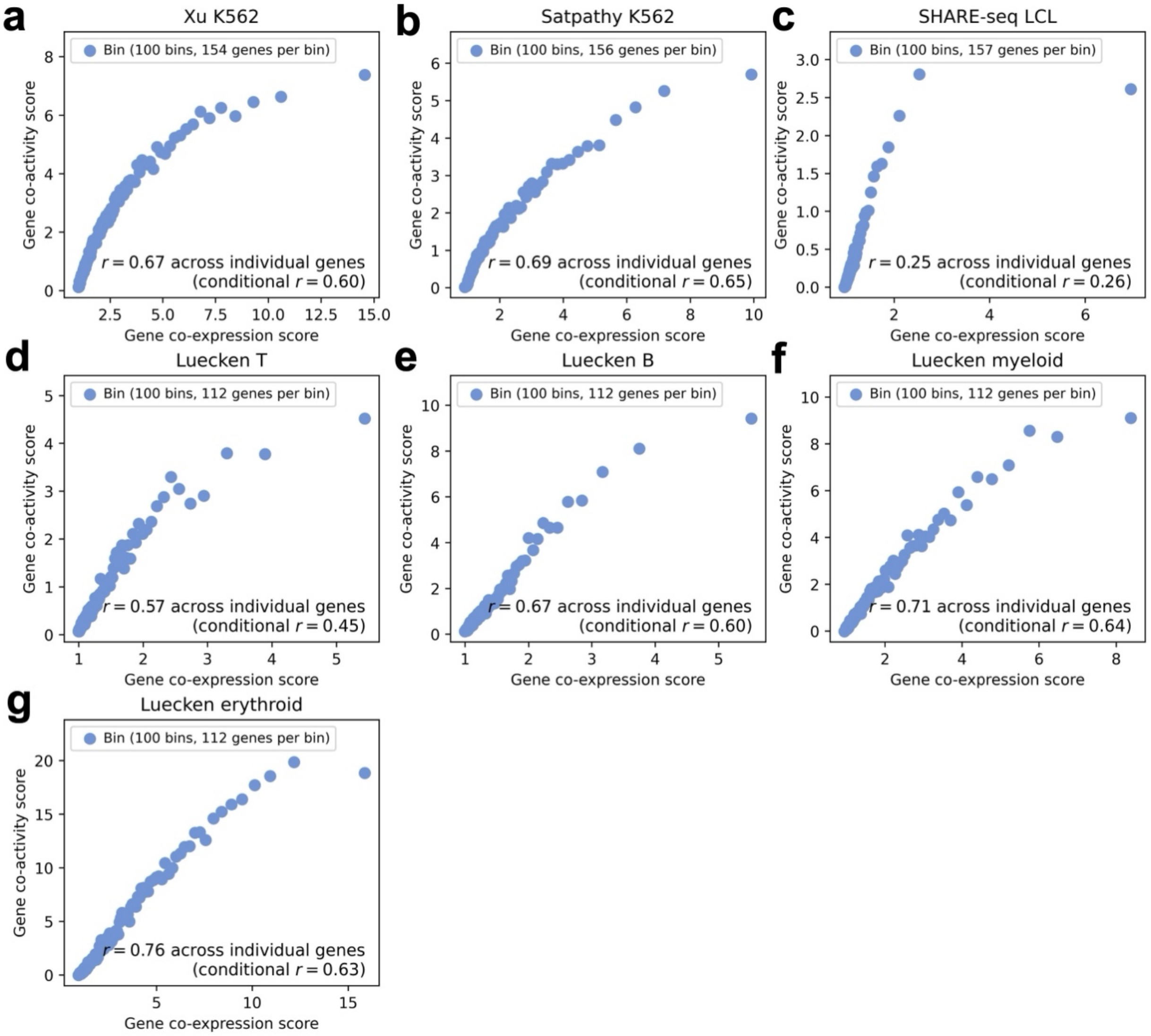


**Supplementary Figure 29. Relationship between gene co-expression score and gene co-activity score.** Relationship between gene co-expression score and gene co-activity score across genes in the **a)** Xu K562, **b)** Satpathy K562, **c)** SHARE-seq LCL, **d)** Luecken T, **e)** Luecken B, **f)** Luecken myeloid, and **g)** Luecken erythroid data set-cell type pairs, computed using peaks and genes within the *cis* window (<1Mb) of each focal gene. ‘Conditional *r*’ denotes correlation conditioned on number of peaks <1Mb from the focal gene. Genes are partitioned equally into 100 bins (each represented by 1 point) by co-expression score. Numerical results are reported in **Supplementary Table 9**.

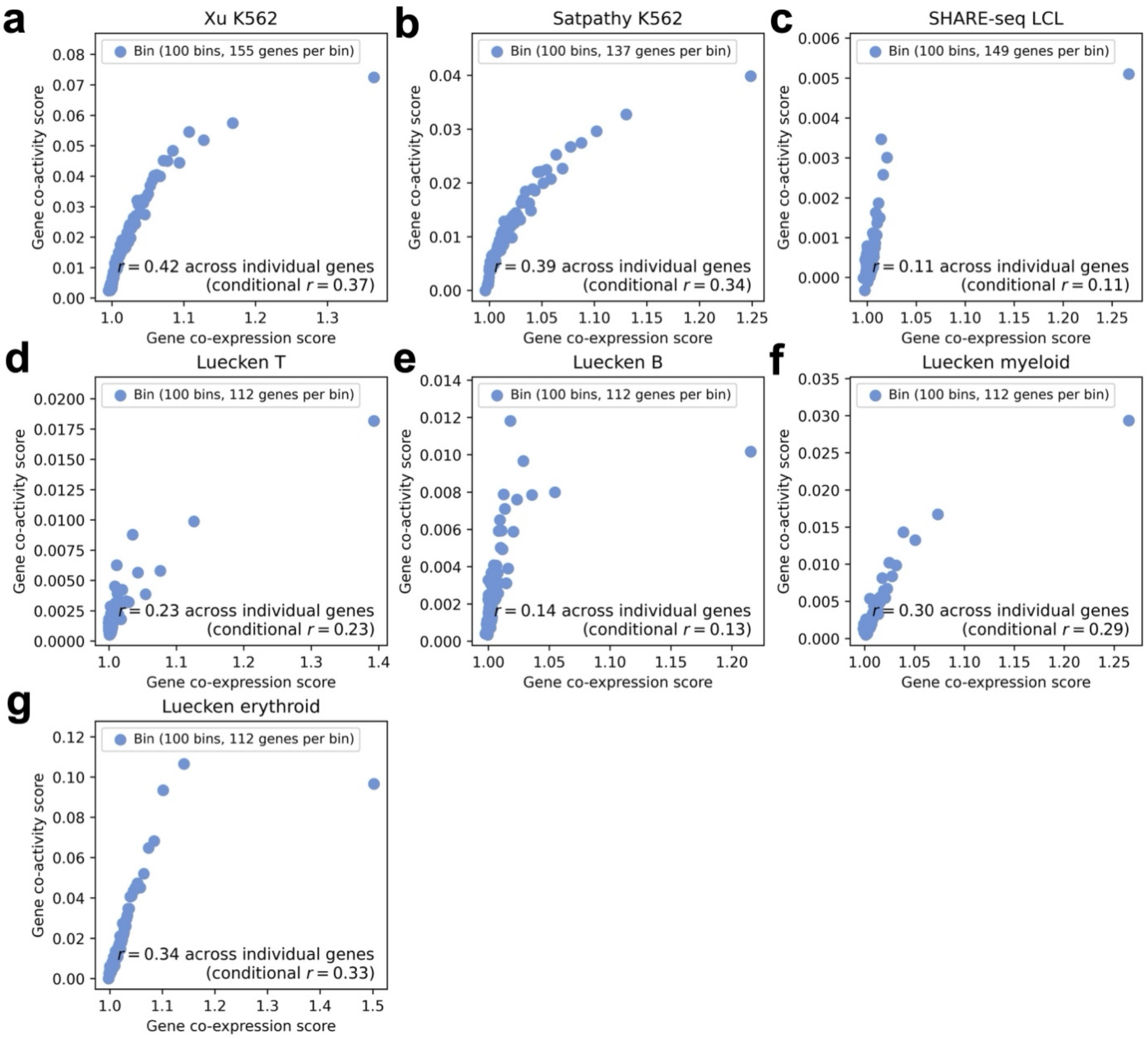


**Supplementary Figure 30. Relationship between gene co-expression score and gene co-activity score using correlations computed across single cells (instead of metacells).** Relationship between gene co-expression score and gene co-activity score across genes in the **a)** Xu K562, **b)** Satpathy K562, **c)** SHARE-seq LCL, **d)** Luecken T, **e)** Luecken B, **f)** Luecken myeloid, and **g)** Luecken erythroid data set-cell type pairs, computed using peaks and genes within the *cis* window (<1Mb) of each focal gene, with peak-peak and peak-gene correlations computed across single cells (instead of metacells). 'Conditional *r*' denotes correlation conditioned on number of peaks <1Mb from the focal gene. Genes are partitioned equally into 100 bins (each represented by 1 point) by co-expression score.

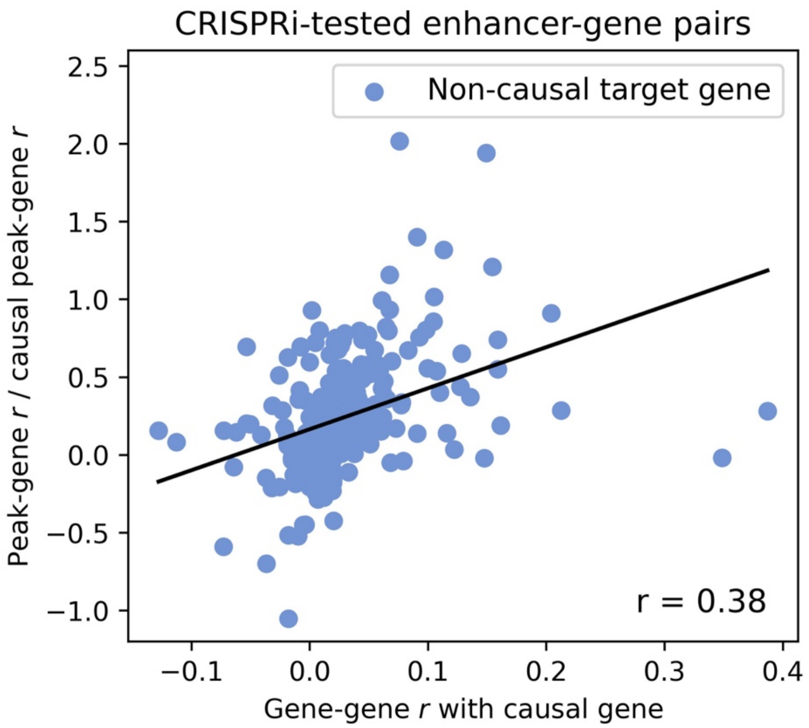


**Supplementary Figure 31. Relationship between non-causal peak-gene correlation and tagging gene correlation using correlations computed across single cells (instead of metacells).** Relationship between correlation with the CRISPR-validated causal target gene (x-axis) and correlation with the causal peak (relative to the causal target gene's correlation with the causal peak) (y-axis) across non-causal target genes tested by CRISPRi, with correlations computed across single cells (instead of metacells). We analyzed CRISPRi-tested peak-gene pairs, restricting to 86 peaks with both an experimentally validated causal (CRISPR-positive) target gene that is identified by Signac (with peak-gene correlation > 0.05 and p-value < 0.05) and at least one experimentally tested non-causal (CRISPR-negative) gene overlapping our single-cell data. We restricted this analysis to the 3 data set-cell type pairs most relevant to K562 cells: Xu K562, Satpathy K562, and Luecken erythroid. We measured the correlation of each CRISPR-negative gene with both the CRISPR-positive gene and the focal peak. Correlation with the CRISPR-positive gene was strongly correlated to correlation with the focal peak.

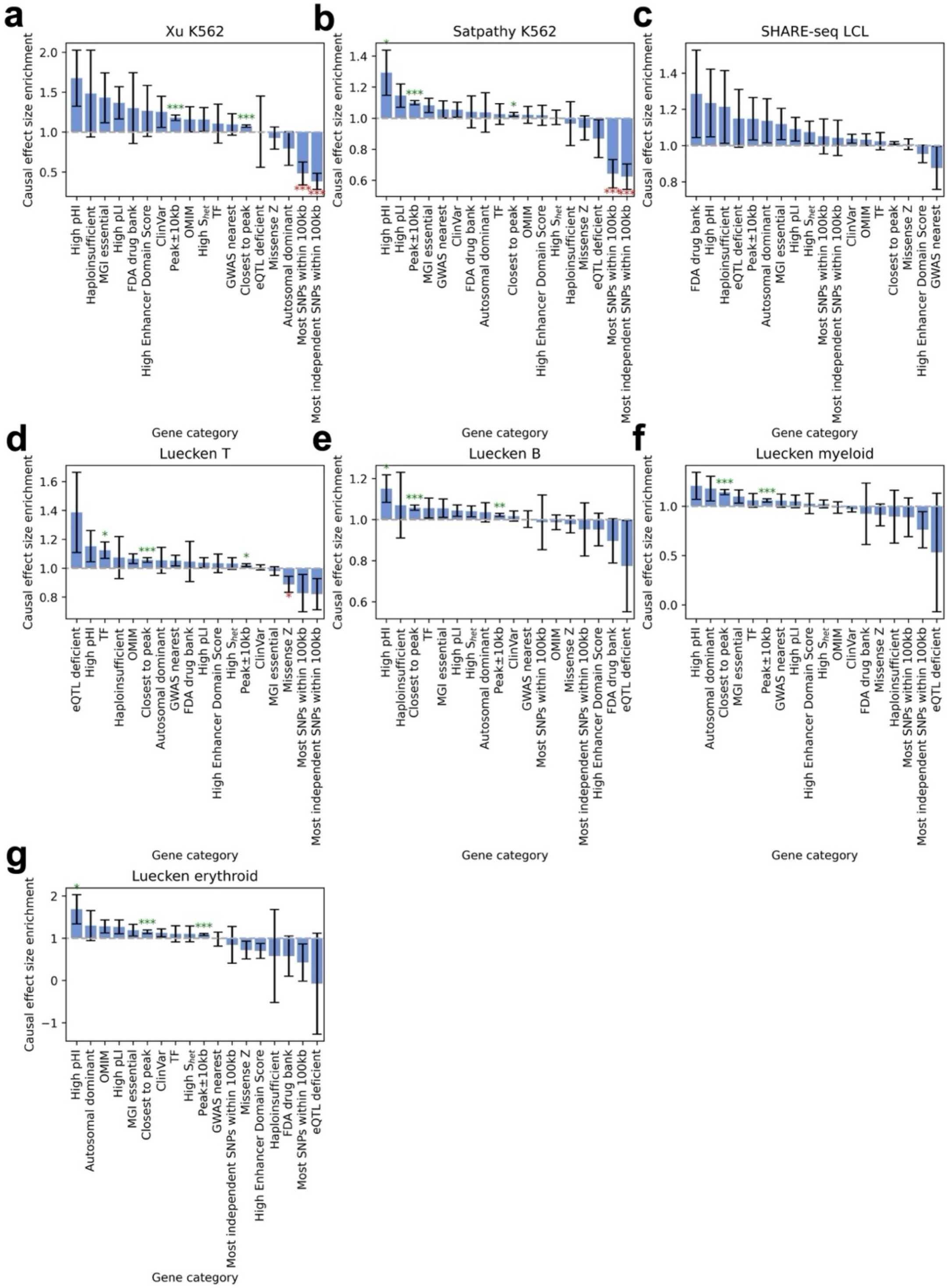


**Supplementary Figure 32. S-CESC causal effect size enrichments.** Causal effect size enrichment of 18 gene categories in stratified co-expression score regression in the **a)** Xu K562, **b)** Satpathy K562, **c)** SHARE-seq LCL, **d)** Luecken T, **e)** Luecken B, **f)** Luecken myeloid, and **g)** Luecken erythroid data set-cell type pairs. Bars and confidence intervals denote estimates and standard errors, respectively. Stars denote meta-analyzed p-values for significant enrichment (green) or depletion (red) (*: $p < 0.05$, **: $p < 0.01$, ***: $p < 0.001$). Numerical results are reported in **Supplementary Table 11**.

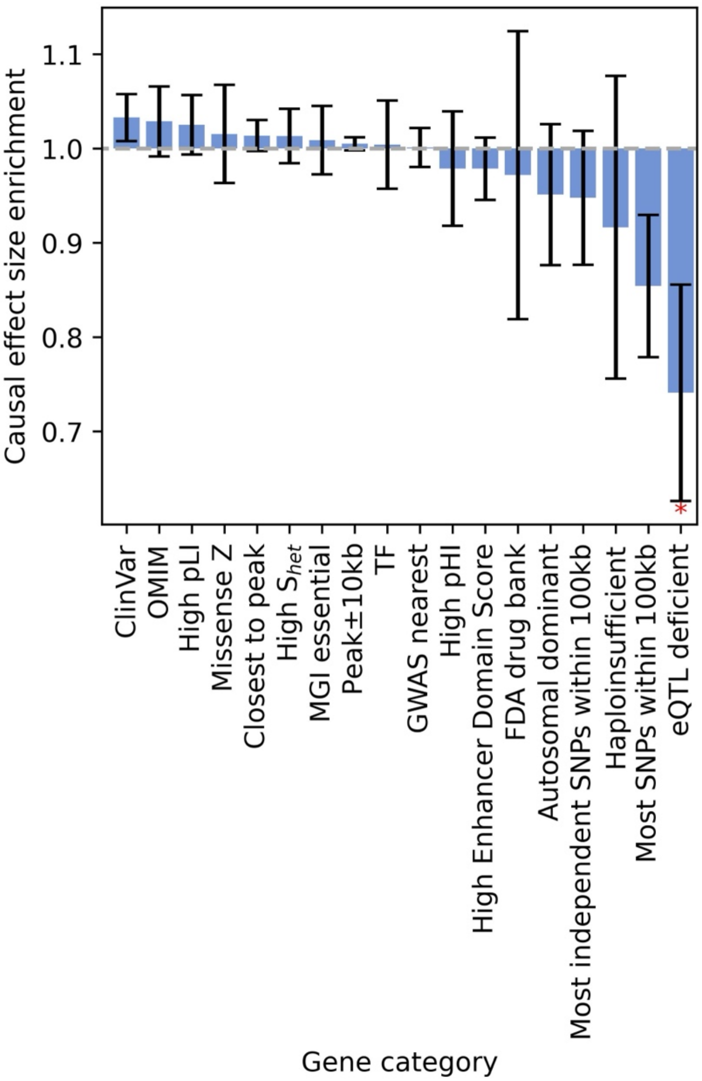


**Supplementary Figure 33. S-CESC causal effect size enrichments computed using correlations across single cells (instead of metacells).** Causal effect size enrichment of 18 gene categories in stratified co-expression score regression, implemented using gene-gene and peak-gene correlations computed across single cells (instead of metacells) and meta-analyzed across all 7 data set-cell type pairs. Bars and confidence intervals denote estimates and standard errors, respectively, meta-analyzed across data set-cell type pairs. Stars denote meta-analyzed p-values for significant enrichment (green) or depletion (red) (*: $p < 0.05$, **: $p < 0.01$, ***: $p < 0.001$).

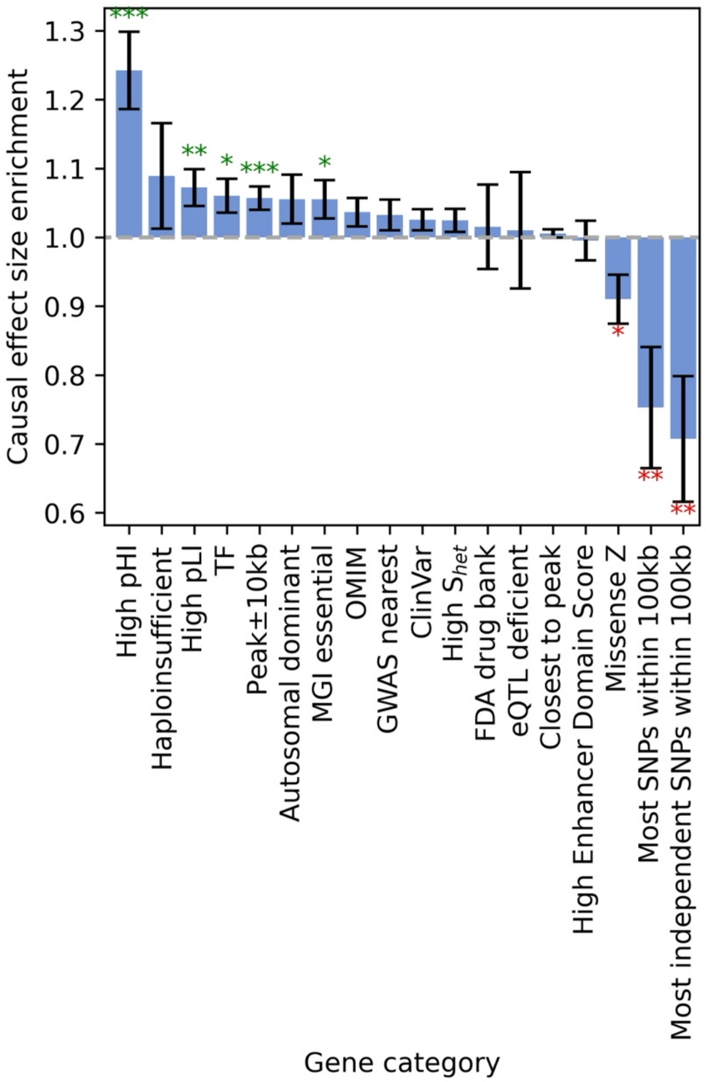


**Supplementary Figure 34. S-CESC causal effect size enrichments without conditioning on number of nearby peaks.** Causal effect size enrichment of 18 peak categories in stratified co-expression score regression across genes, implemented without the covariate for the number of peaks <1Mb from each gene and meta-analyzed across all 7 data set-cell type pairs. Results are generally similar to **Figure 4c**. Bars and confidence intervals denote estimates and standard errors, respectively, meta-analyzed across data set-cell type pairs. Stars denote meta-analyzed p-values for significant enrichment (green) or depletion (red) (*: $p < 0.05$, **: $p < 0.01$, ***: $p < 0.001$).

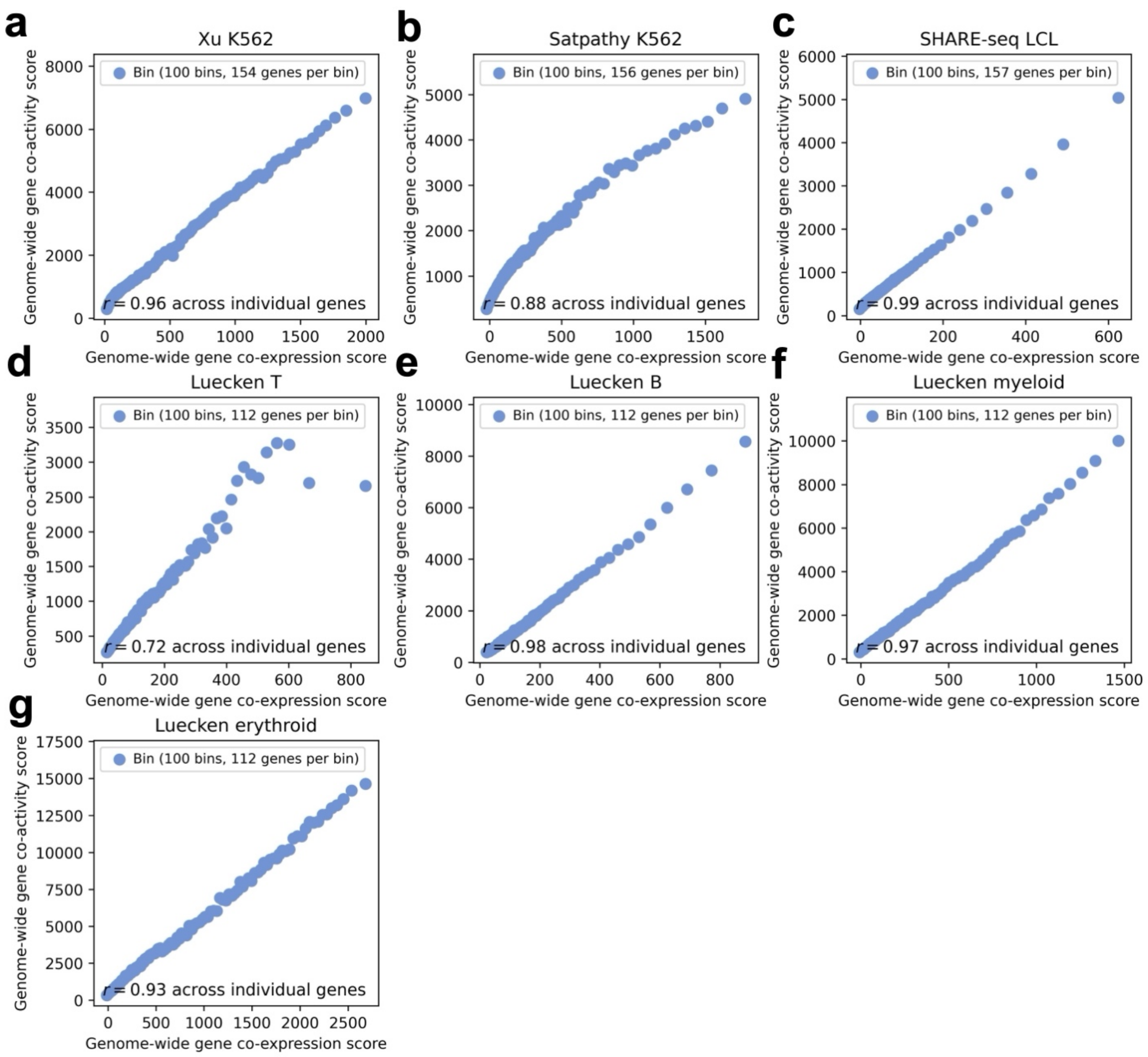


**Supplementary Figure 35. Relationship between genome-wide gene co-expression score and genome-wide gene co-activity score.** Relationship between genome-wide gene co-expression score and genome-wide gene co-activity score across peaks in the **a)** Xu K562, **b)** Satpathy K562, **c)** SHARE-seq LCL, **d)** Luecken T, **e)** Luecken B, **f)** Luecken myeloid, and **g)** Luecken erythroid data set-cell type pairs, computed using peaks and genes within the *cis* window (<1Mb) of each focal gene. 'Conditional *r*' denotes correlation conditioned on number of peaks <1Mb from the focal gene. Genes are partitioned equally into 100 bins (each represented by 1 point) by co-expression score. Numerical results are reported in **Supplementary Table 12**.

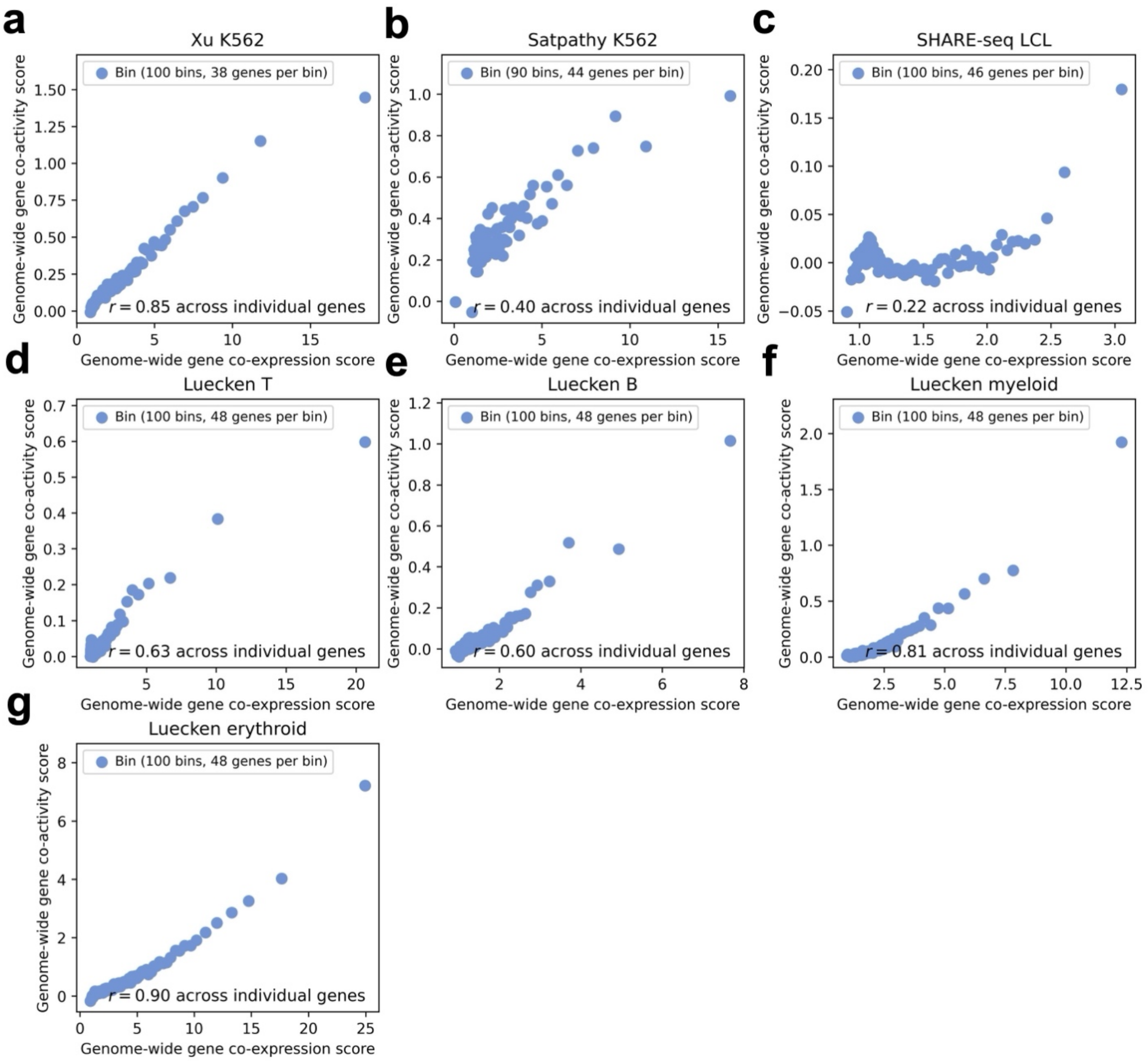


**Supplementary Figure 36. Relationship between genome-wide gene co-expression score and genome-wide gene co-activity score computed using correlations across single cells (instead of metacells).** Relationship between genome-wide gene co-expression score and genome-wide gene co-activity score (using a subset of 5000 peaks and 5000 genes) across peaks in the **a)** Xu K562, **b)** Satpathy K562, **c)** SHARE-seq LCL, **d)** Luecken T, **e)** Luecken B, **f)** Luecken myeloid, and **g)** Luecken erythroid data set-cell type pairs, computed across single cells (instead of metacells) using all peaks and genes genome-wide. We downsampled to 5000 randomly selected peaks and 5000 randomly selected genes when computing scores (due to the high computational cost of computing correlations for every peak-peak, gene-gene, and peak-gene pair genome-wide across single cells). Genes are partitioned equally into 100 bins (each represented by 1 point) by co-expression score.

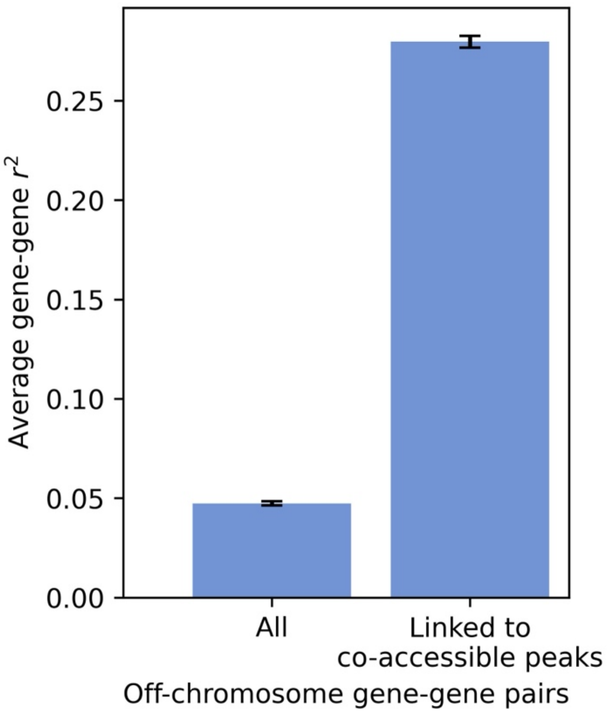


**Supplementary Figure 37. Average squared gene-gene correlation for off-chromosome pairs of genes linked to co-accessible peaks vs. all genes.** We identified 2,108 proximal peak-gene links in the Xu K562 data set (defining “proximal” as peak-gene distance <100kb and “links” as ArchR linking score ($r^2$) > 0.2025, corresponding to ArchR threshold $r$ > 0.45). From these peak-gene links, we identified 644,981 highly co-accessible pairs of off-chromosome linked peaks (involving a peak from chromosome 1 and a peak from a different chromosome, defining “highly co-accessible” as peak-peak $r^2$ > 0.2025). We then identified 124,714 pairs of genes linked to peaks forming an off-chromosome co-accessible peak-peak pair (such that each gene-gene pair also has a gene on chromosome 1 and a gene on a different chromosome). We then computed the average gene-gene $r^2$ across these gene-gene pairs (right) vs. all gene-gene pairs formed from the set of genes involved in a proximal peak-gene link (left). Average gene-gene $r^2$ for pairs of genes linked to highly co-accessible pairs of peaks was substantially higher ($r^2$ = 0.28) than for all gene-gene pairs ($r^2$ = 0.047; p-value for difference = 4e-15). Confidence intervals denote standard errors.

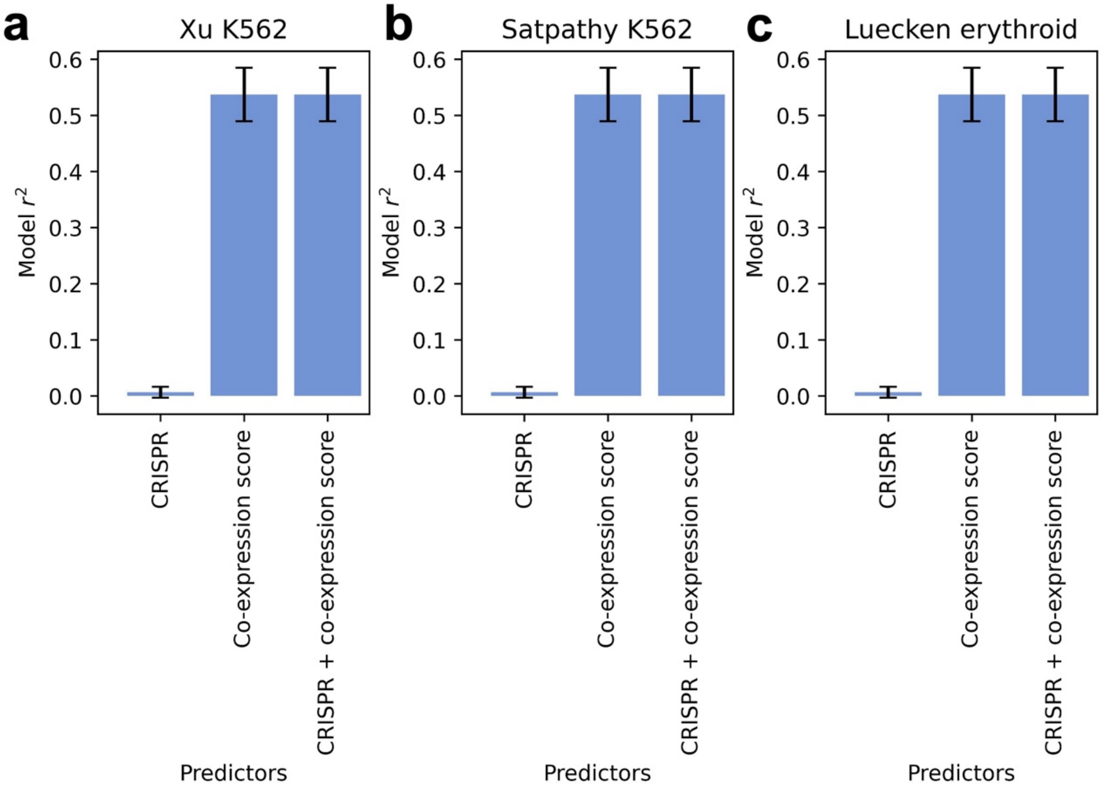


**Supplementary Figure 38. Regression of gene co-activity score on CRISPR status and gene co-expression score.** Model $r^2$ for linear regressions of gene co-activity score on CRISPR-positive status (“CRISPR”), gene co-expression score, or both, for the **a)** Xu K562, **b)** Satpathy K562, and **c)** Luecken erythroid data set-cell type pairs. Genes in more “active” regulatory regions (higher gene co-expression scores) may be more likely to be true causal target genes. To assess the role of biological causality in driving the correlation between gene co-activity score and gene co-expression score, we performed a bivariate linear regression of gene co-activity score on gene co-expression score and a binary indicator of CRISPR-positive status (1 if a gene has at least one causal peak validated by CRISPR, 0 otherwise) across genes. We also performed univariate regressions of gene co-activity score on gene co-expression score and CRISPR-positive status individually. We restricted this analysis to the 3 data set-cell type pairs most relevant to K562 cells: Xu-K562, Satpathy-K562, and Luecken-erythroid. Within each data set-cell type pair, we further restricted to peaks with ≥1 causal target gene validated by CRISPR (CRISPR-positive) and ≥1 CRISPR-tested non-target (CRISPR-negative) gene. Confidence intervals denote standard errors.

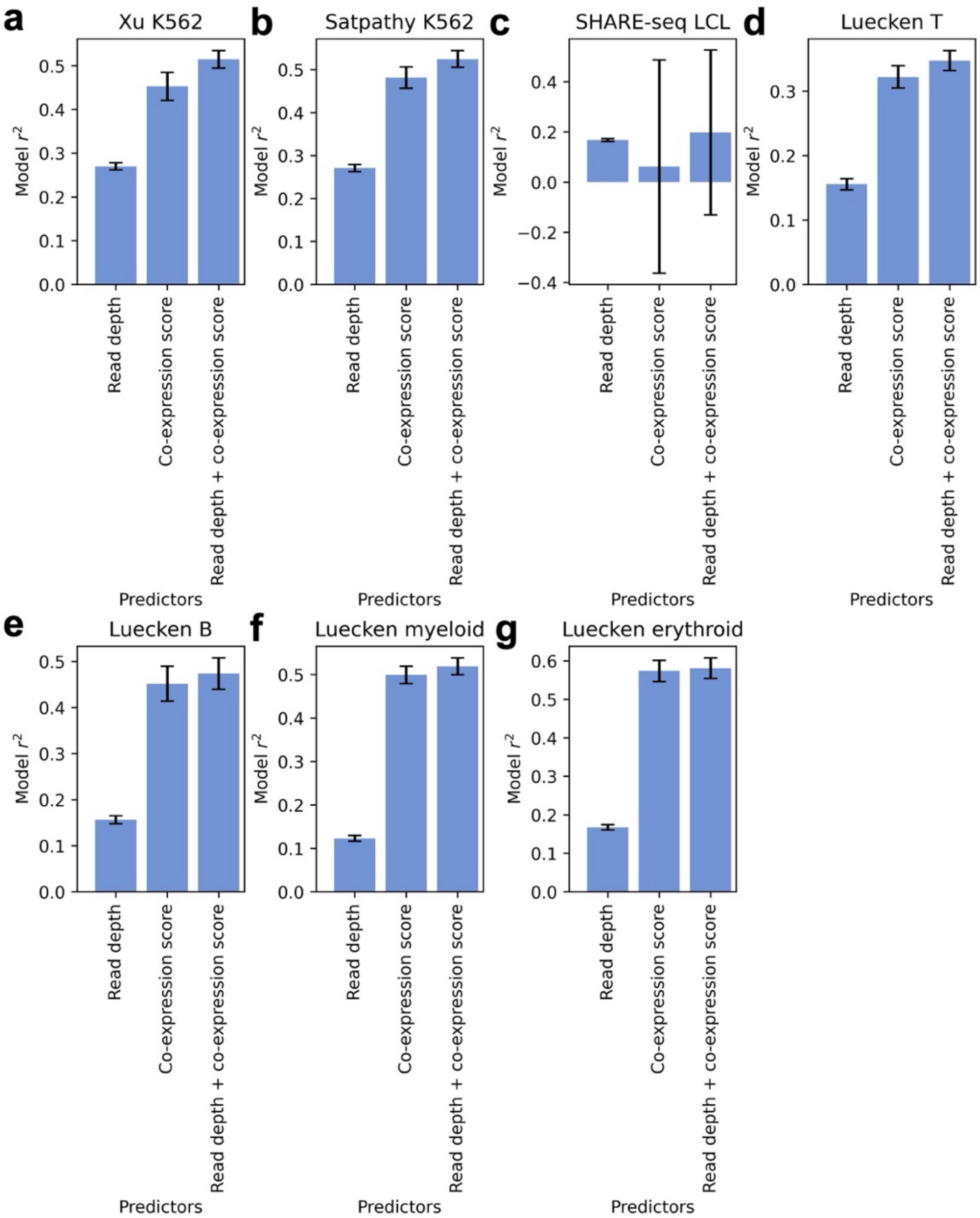


**Supplementary Figure 39. Regression of gene co-activity score on gene read depth and gene co-expression score.** Model $r^2$ for linear regressions of gene co-activity score on gene read depth, gene co-expression score, or both in the **a)** Xu K562, **b)** Satpathy K562, **c)** SHARE-seq LCL, **d)** Luecken T, **e)** Luecken B, **f)** Luecken myeloid, and **g)** Luecken erythroid data set-cell type pairs. Genes with higher read depth may have higher measured correlations with other peaks and genes, generating higher gene co-expression scores and gene co-activity scores. To assess the role of gene read depth in driving the correlation between gene co-activity score and gene co-expression scores, we performed a bivariate linear regression of gene co-activity score on gene co-expression score and log(read depth) across genes. We also performed univariate regressions of gene co-activity score on gene co-expression scores and read depth individually. Confidence intervals denote standard errors. (We note that log-transformed read depth attained higher $r^2$ than raw read depth.)

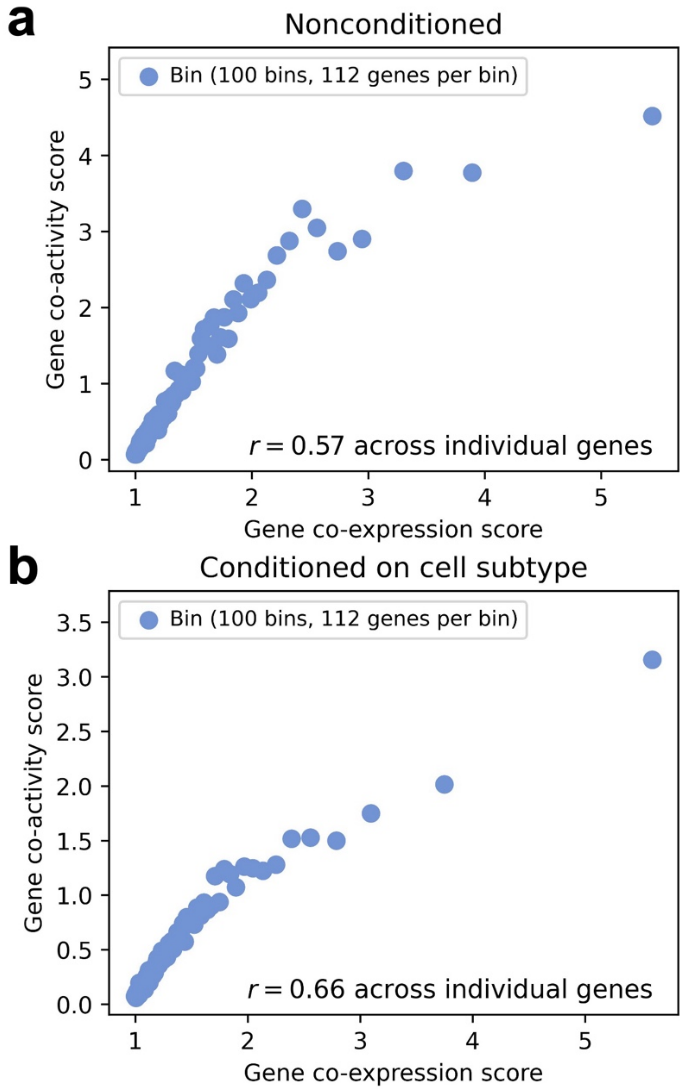


**Supplementary Figure 40. Relationship between gene co-expression score and gene co-activity score conditioned on cell subtype.** Relationship between gene co-expression score and gene co-activity score across peaks in the Luecken T cell data set computed using **a)** unconditioned peak-peak and peak-gene correlations and **b)** gene-gene and peak-gene correlations conditioned on cell subtype proportions. Cell subtypes may be associated with varying levels of average peak accessibility and gene expression, driving a correlation between gene co-expression score and gene co-activity score. To assess the role of cell subtypes, we analyzed T cells from the Luecken BMMC data set, which span 4 labeled T cell subtypes (activated CD4+, naive CD4+, CD8+, and naive CD8+; each represented by 1-12k cells). We separately residualized gene expression and ATAC peak accessibility by cell type proportions in each metacell (via linear regression across metacells), recomputed gene-gene correlations and peak-gene correlations using residualized data, and recomputed gene co-expression scores and gene co-activity scores by summing these conditional correlations. The correlation between gene co-activity score and gene co-expression score did not decrease after accounting for cell subtypes, suggesting that major cell subtypes do not fully explain the correlation. Genes are partitioned equally into 100 bins (each represented by 1 point) by gene co-expression score.

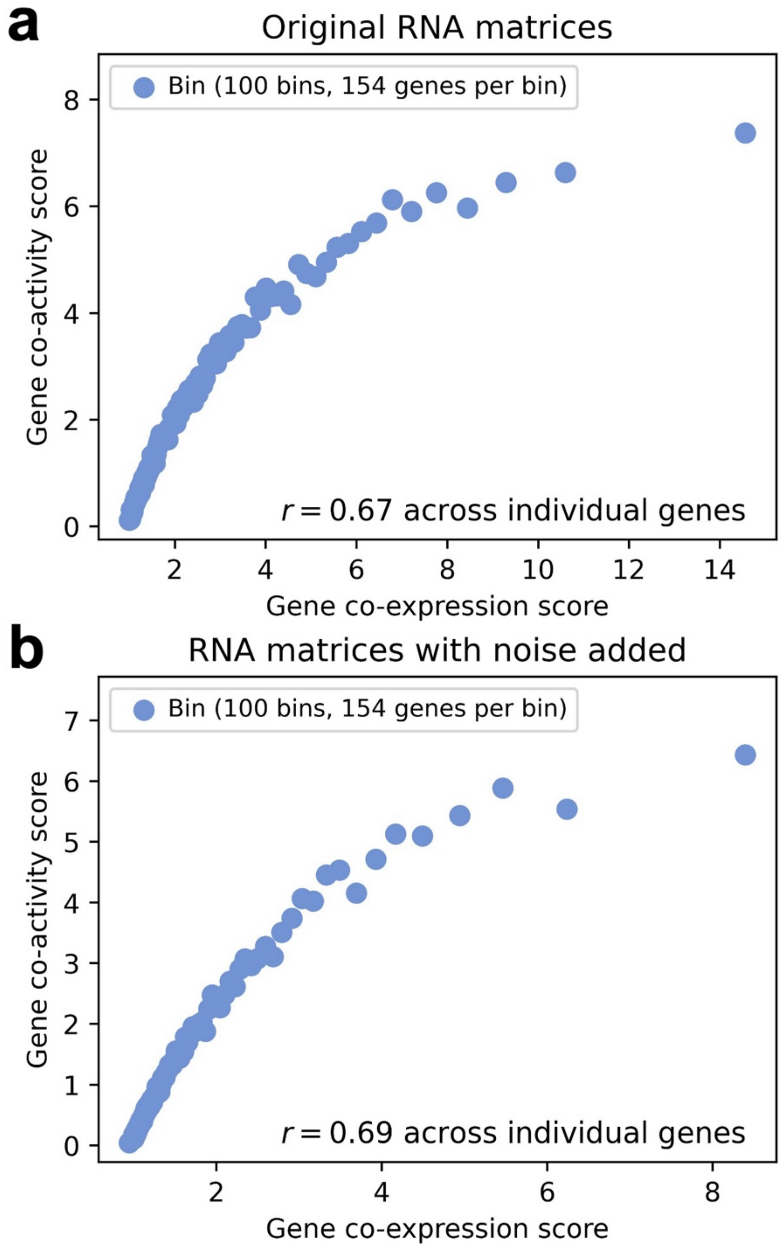


**Supplementary Figure 41**. **Relationship between gene co-expression score and gene co-activity score with RNA measurement noise added.** Relationship between gene co-expression score and gene co-activity score across peaks in the Xu K562 data set computed using **a)** the original RNA matrix and **b)** an RNA matrix with added measurement noise. Genes with higher amounts of measurement noise may have lower measured correlations with other peaks and genes, possibly driving the correlation between gene co-activity score and gene co-expression score. Because it is difficult to directly assess the amount of measurement noise in gene expression data, we introduced additional artificial noise to 50% of genes (by randomly setting 20% of entries equal to 0, retaining the original data for the remaining 50% of genes), and recomputed gene co-expression and gene co-activity scores.

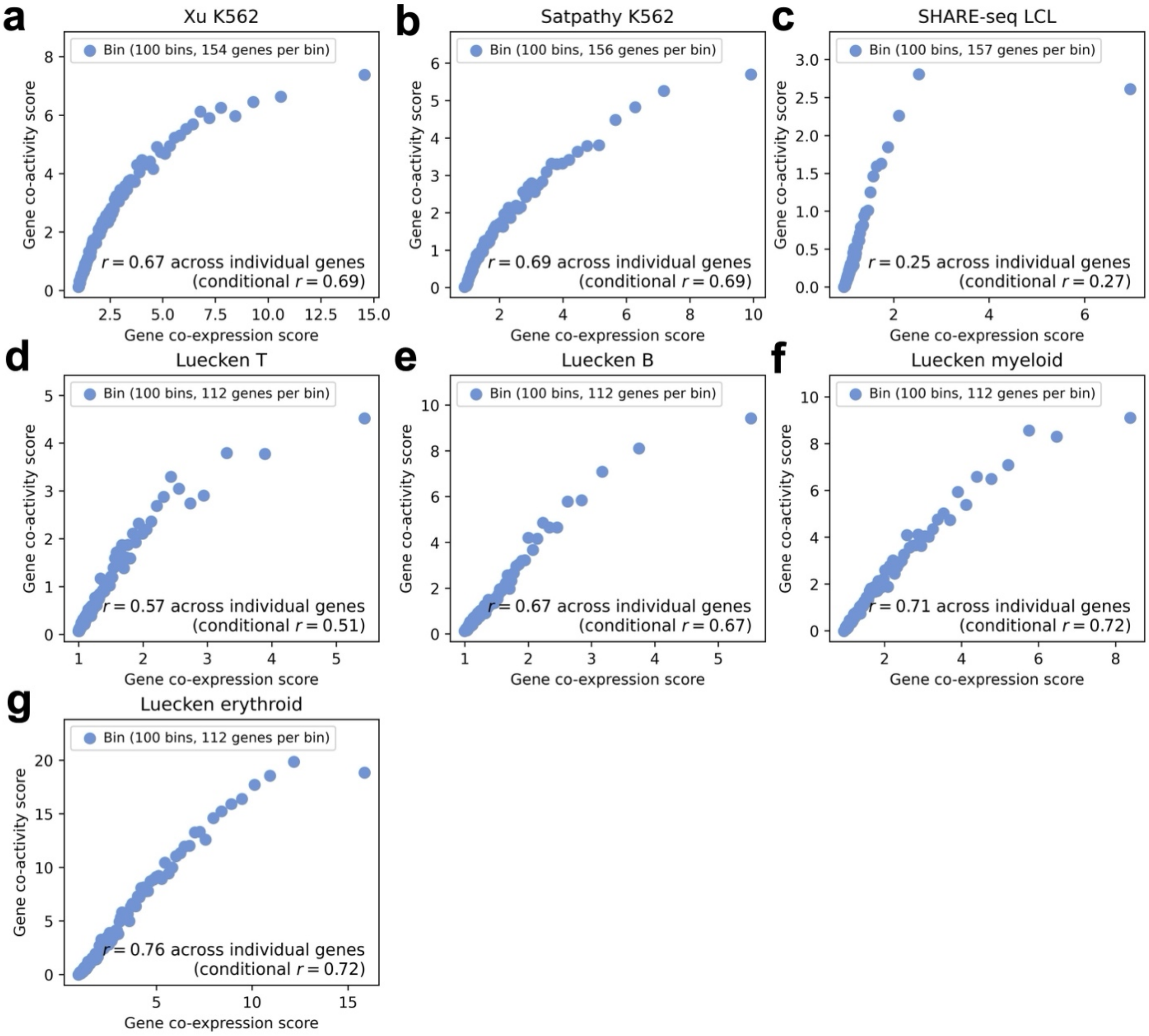


**Supplementary Figure 42. Relationship between gene co-expression score and gene co-activity score conditioned on number of nearby peaks and genes.** Relationship between gene co-expression score and gene co-activity score across genes in the **a)** Xu K562, **b)** Satpathy K562, **c)** SHARE-seq LCL, **d)** Luecken T, **e)** Luecken B, **f)** Luecken myeloid, and **g)** Luecken erythroid data set-cell type pairs, computed using peaks and genes within the *cis* window (<1Mb) of each focal gene. 'Conditional *r*' denotes correlation conditioned on both the number of peaks <1Mb from the focal gene and the number of genes <1Mb from the focal gene (instead of only the number of peaks <1Mb from the focal gene). Genes are partitioned equally into 100 bins (each represented by 1 point) by gene co-expression score.

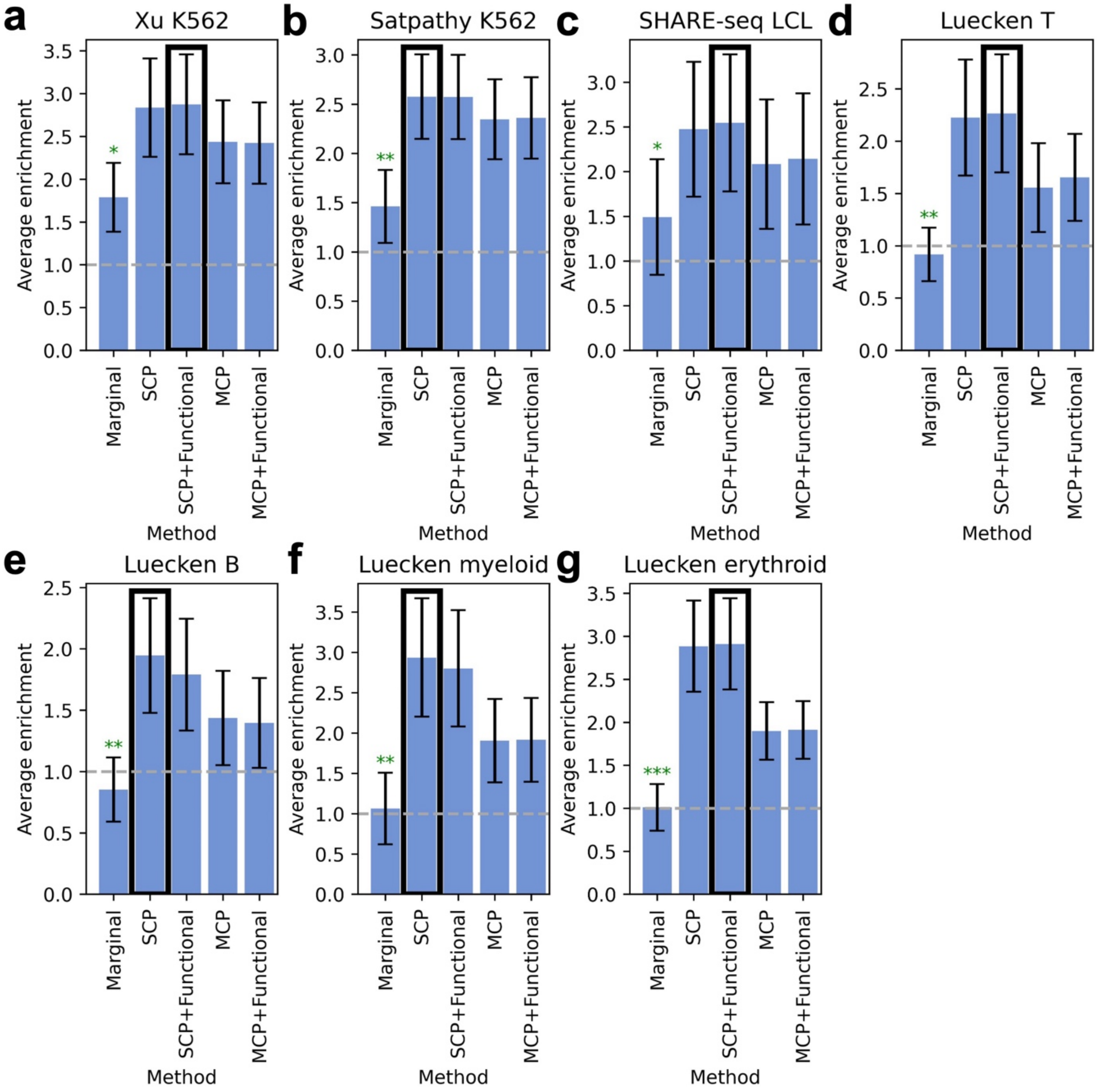


**Supplementary Figure 43. Evaluation of ArchR vs. fine-mapped ArchR scores on CRISPR data.** Average enrichment across recall values of links predicted by ArchR vs. fine-mapped ArchR scores for 448 links validated by CRISPR in the **a)** Xu K562, **b)** Satpathy K562, **c)** SHARE-seq LCL, **d)** Luecken T, **e)** Luecken B, **f)** Luecken myeloid, and **g)** Luecken erythroid data set-cell type pairs. Bars and confidence intervals denote estimates and standard errors, respectively. Green asterisks denote significant underperformance of the focal method vs. the top-performing method (denoted with a black outline) (*: $p < 0.05$, **: $p < 0.01$, ***: $p < 0.001$). Numerical results are reported in **Supplementary Table 15**. Grey horizontal dashed lines mark 1.0x enrichment. SCP, single causal peak fine-mapping; SCP + Functional, single causal peak functionally informed fine-mapping; MCP, multiple causal peak fine-mapping; MCP + Functional, multiple causal peak functionally informed fine-mapping.

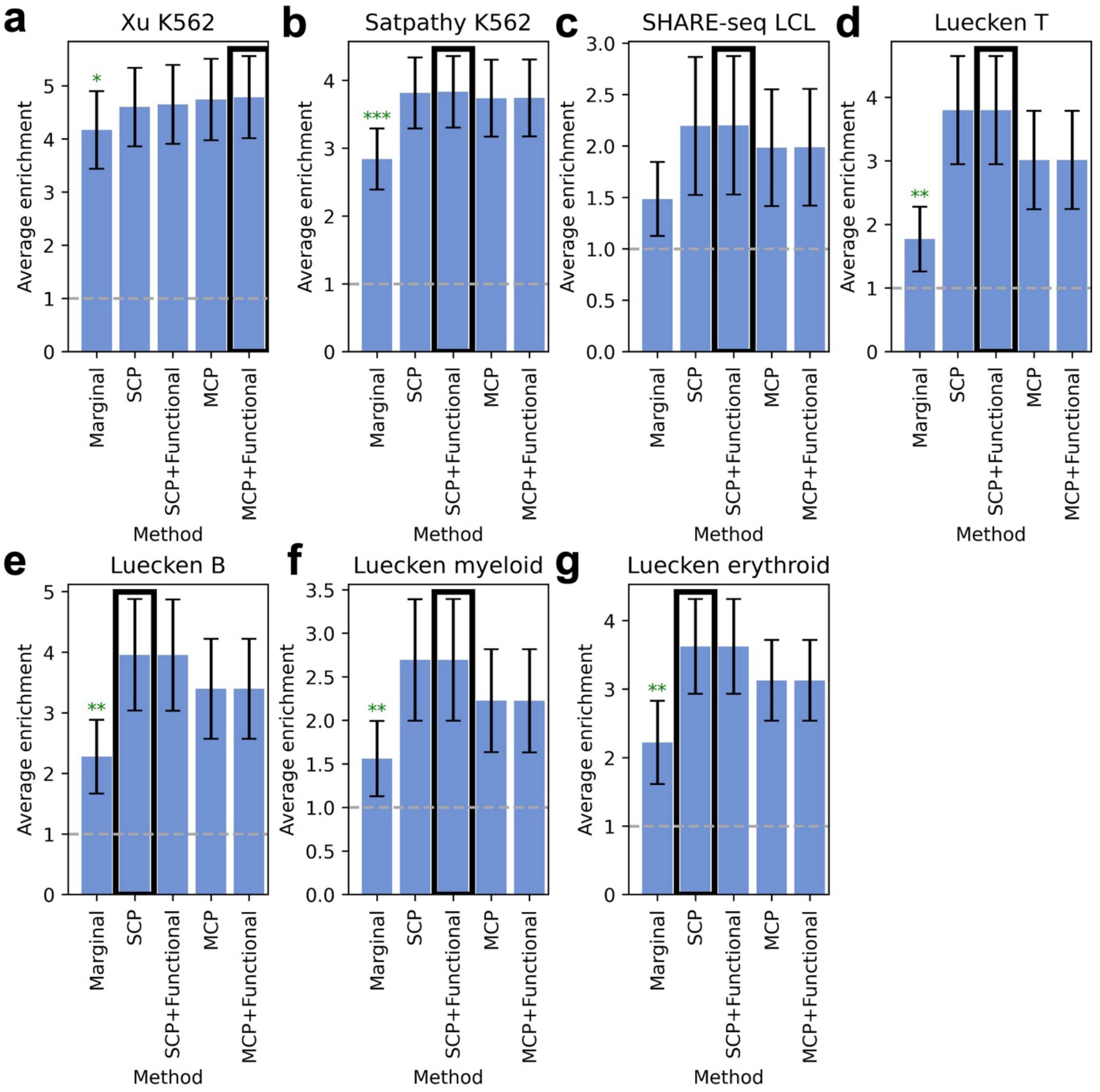


**Supplementary Figure 44. Evaluation of Signac vs. fine-mapped Signac scores on CRISPR data.** Average enrichment across recall values of links predicted by Signac vs. fine-mapped Signac scores for 448 links validated by CRISPR in the **a)** Xu K562, **b)** Satpathy K562, **c)** SHARE-seq LCL, **d)** Luecken T, **e)** Luecken B, **f)** Luecken myeloid, and **g)** Luecken erythroid data set-cell type pairs. Bars and confidence intervals denote estimates and standard errors, respectively. Green asterisks denote significant underperformance of the focal method vs. the top-performing method (denoted with a black outline) (*: $p < 0.05$, **: $p < 0.01$, ***: $p < 0.001$). Numerical results are reported in **Supplementary Table 16**. Grey horizontal dashed lines mark 1.0x enrichment. SCP, single causal peak fine-mapping; SCP + Functional, single causal peak functionally informed fine-mapping; MCP, multiple causal peak fine-mapping; MCP + Functional, multiple causal peak functionally informed fine-mapping.

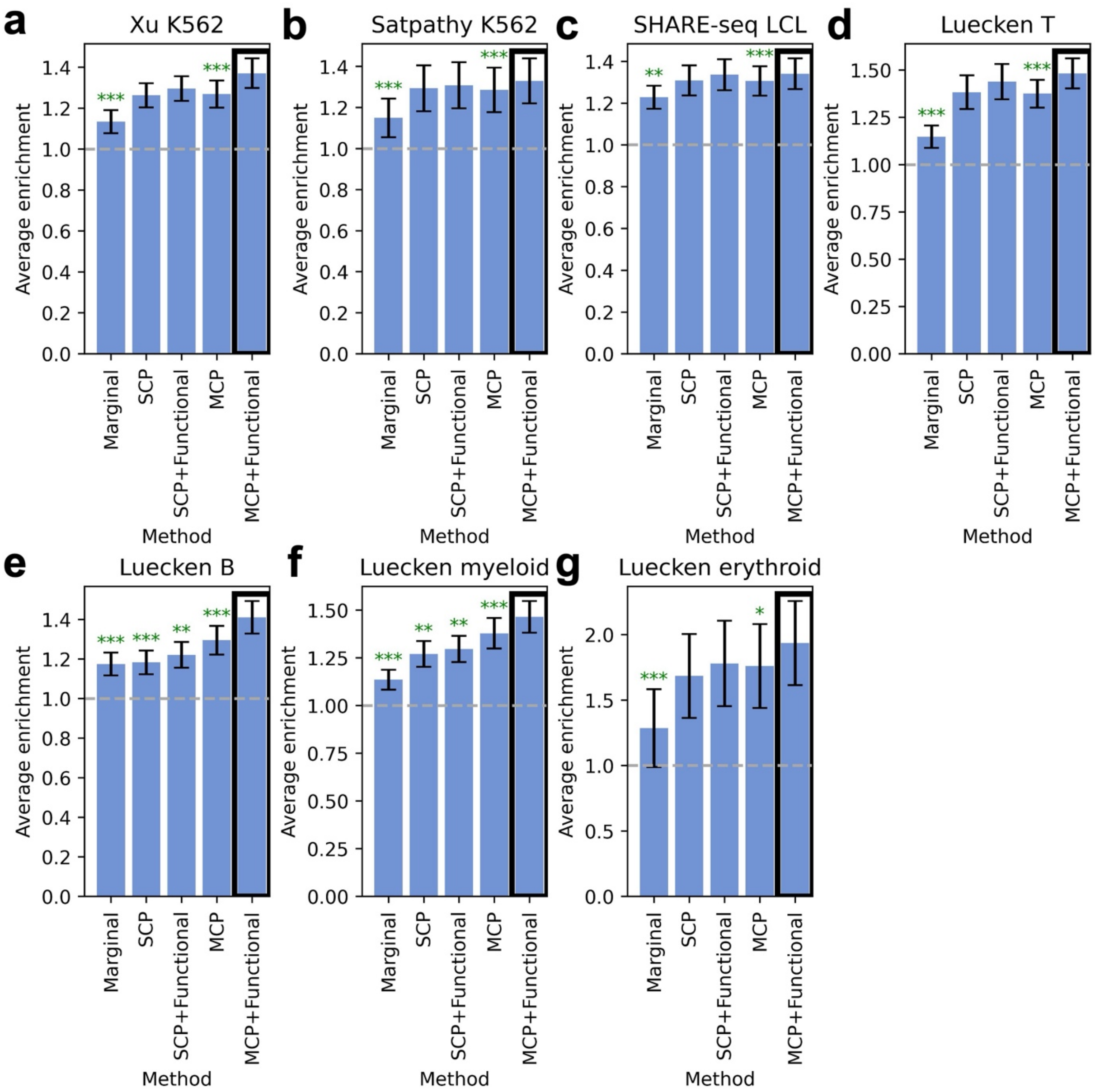


**Supplementary Figure 45. Evaluation of ArchR vs. fine-mapped ArchR scores on eQTL data.** Average enrichment across recall values of links predicted by ArchR vs. fine-mapped ArchR scores for 39,194 fine-mapped eSNP–eGene pairs attaining maximum PIP > 0.5 across GTEx tissues in the **a)** Xu K562, **b)** Satpathy K562, **c)** SHARE-seq LCL, **d)** Luecken T, **e)** Luecken B, **f)** Luecken myeloid, and **g)** Luecken erythroid data set-cell type pairs. Bars and confidence intervals denote estimates and standard errors, respectively. Green asterisks denote significant underperformance of the focal method vs. the top-performing method (denoted with a black outline) (*: $p < 0.05$, **: $p < 0.01$, ***: $p < 0.001$). Numerical results are reported in **Supplementary Table 17**. Grey horizontal dashed lines mark 1.0x enrichment. SCP, single causal peak fine-mapping; SCP + Functional, single causal peak functionally informed fine-mapping; MCP, multiple causal peak fine-mapping; MCP + Functional, multiple causal peak functionally informed fine-mapping.

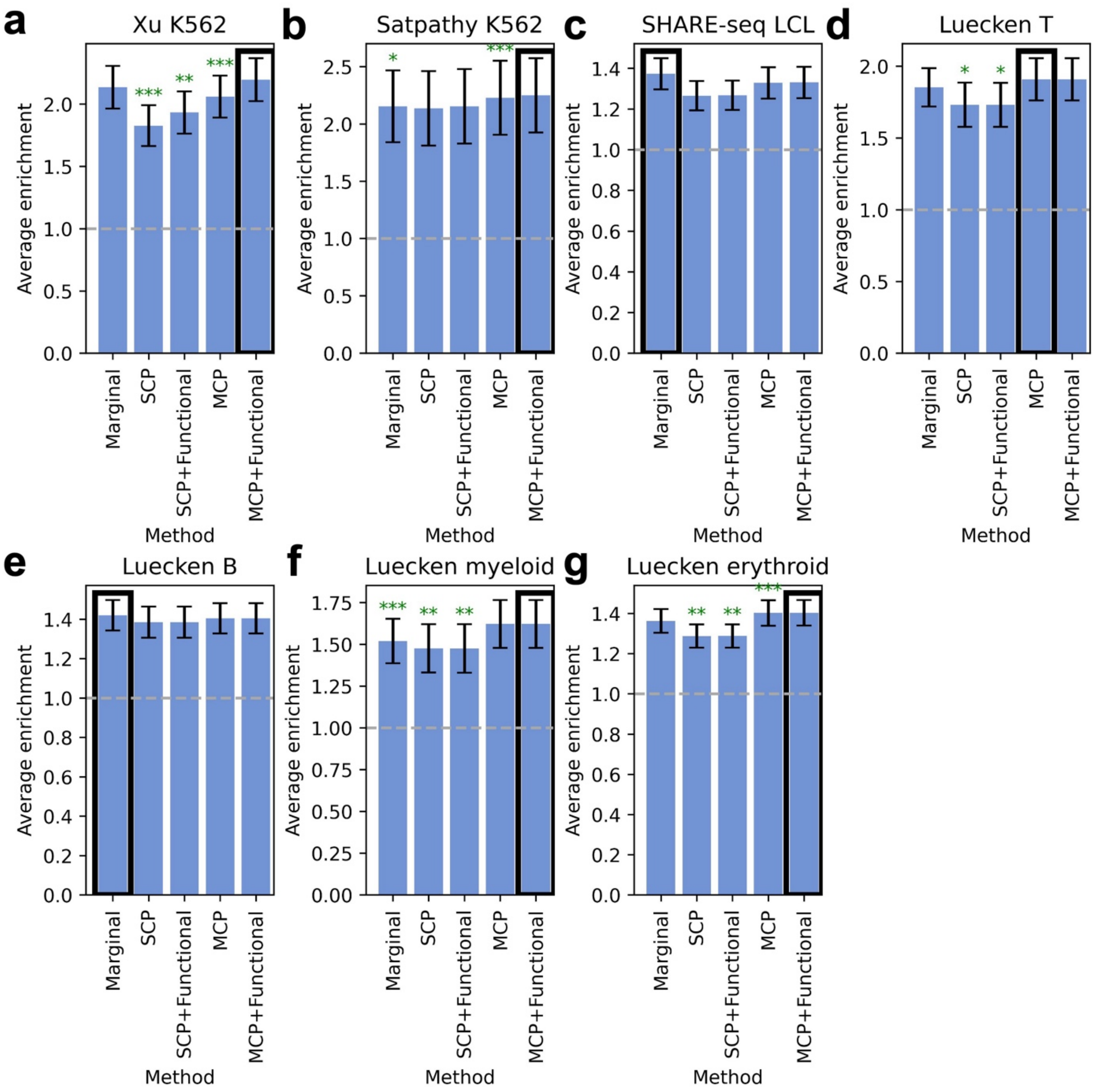


**Supplementary Figure 46. Evaluation of Signac vs. fine-mapped Signac scores on eQTL data.** Average enrichment across recall values of links predicted by Signac vs. fine-mapped Signac scores for 39,194 fine-mapped eSNP–eGene pairs attaining maximum PIP > 0.5 across GTEx tissues in the **a)** Xu K562, **b)** Satpathy K562, **c)** SHARE-seq LCL, **d)** Luecken T, **e)** Luecken B, **f)** Luecken myeloid, and **g)** Luecken erythroid data set-cell type pairs. Bars and confidence intervals denote estimates and standard errors, respectively. Green asterisks denote significant underperformance of the focal method vs. the top-performing method (denoted with a black outline) (*: $p < 0.05$, **: $p < 0.01$, ***: $p < 0.001$). Numerical results are reported in **Supplementary Table 18**. Grey horizontal dashed lines mark 1.0x enrichment. SCP, single causal peak fine-mapping; SCP + Functional, single causal peak functionally informed fine-mapping; MCP, multiple causal peak fine-mapping; MCP + Functional, multiple causal peak functionally informed fine-mapping.

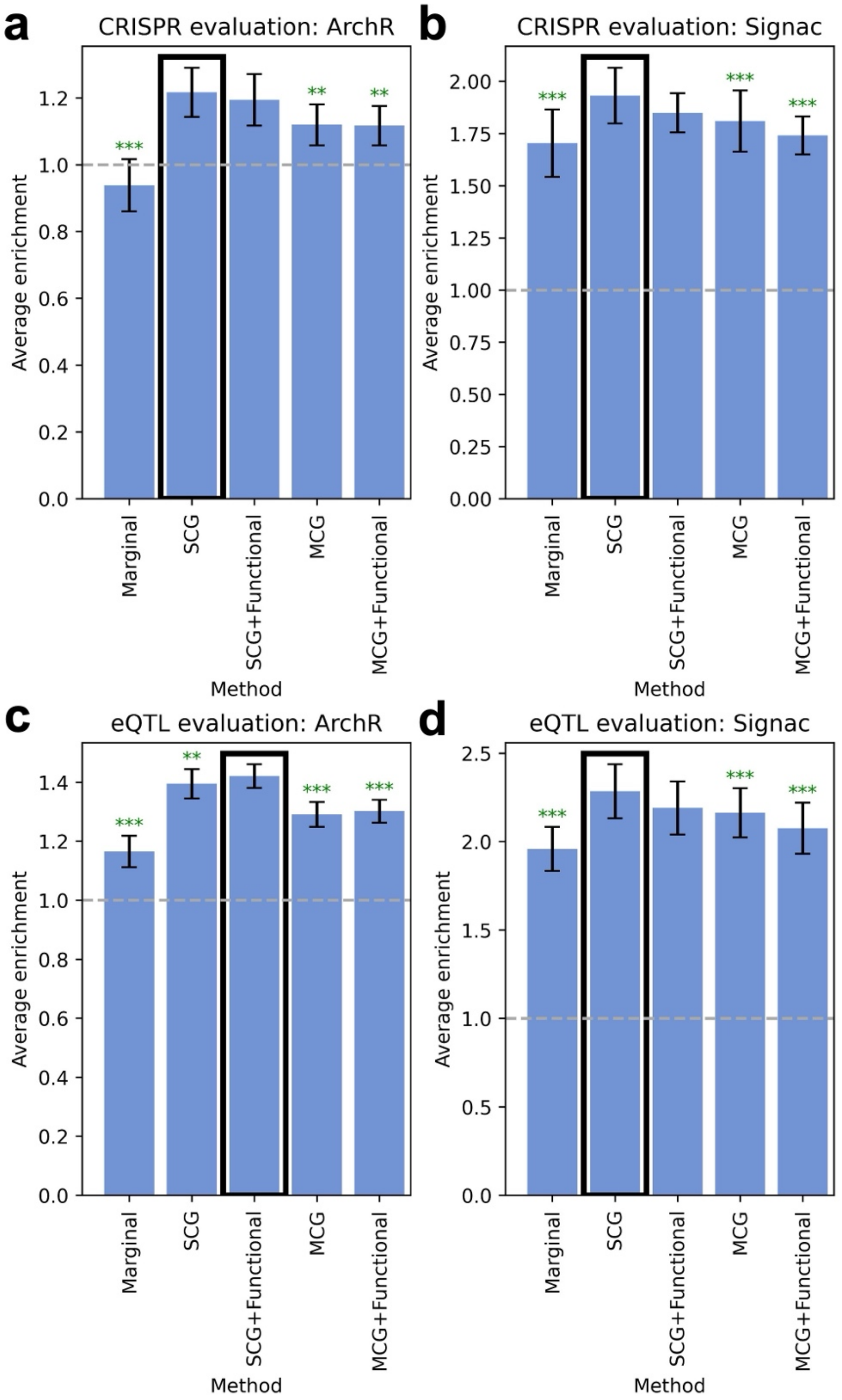


**Supplementary Figure 47. Evaluations of gene-level fine-mapping on CRISPR and eQTL data. a)** Average enrichment across recall values of links predicted by ArchR vs. gene-level fine-mapped ArchR scores for 448 links validated by CRISPR. **b)** Average enrichment across recall values of links predicted by Signac vs. gene-level fine-mapped Signac scores for 448 links validated by CRISPR. **c)** Average enrichment across recall values of links predicted by ArchR vs. gene-level fine-mapped ArchR scores for 39,194 fine-mapped eSNP–eGene pairs attaining maximum PIP > 0.5 across GTEx tissues. **d)** Average enrichment across recall values of links predicted by Signac vs. gene-level fine-mapped Signac scores for 39,194 fine-mapped eSNP–

eGene pairs attaining maximum PIP > 0.5 across GTEx tissues. Details about gene-level fine-mapping are provided in the **Methods** section. Bars and confidence intervals denote estimates and standard errors, respectively, meta-analyzed across data set-cell type pairs. Green asterisks denote significant underperformance of the focal method vs. the top-performing method (denoted with a black outline) (*: $p < 0.05$, **: $p < 0.01$, ***: $p < 0.001$). Grey horizontal dashed lines mark 1.0x enrichment. SCG, single causal gene fine-mapping; SCG + Functional, single causal gene functionally informed fine-mapping; MCG, multiple causal gene fine-mapping; MCG + Functional, multiple causal gene functionally informed fine-mapping.

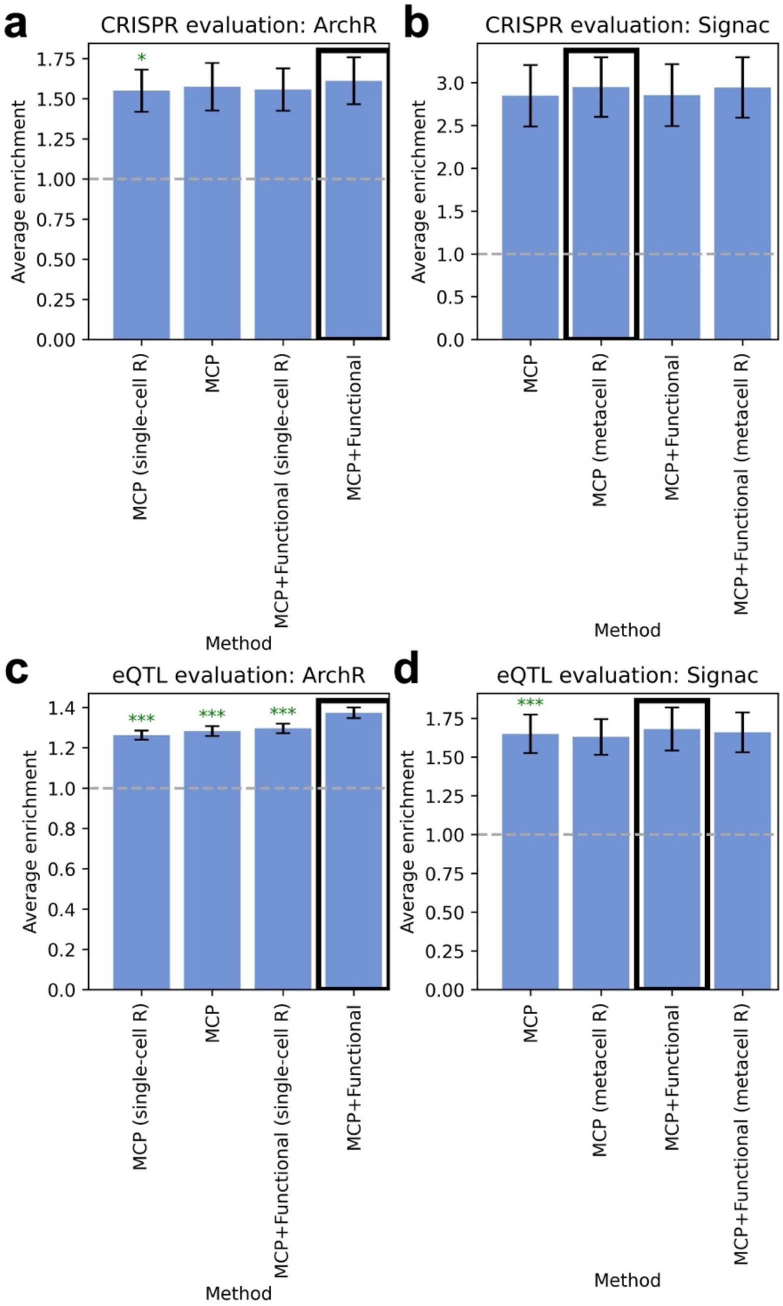


**Supplementary Figure 48. Evaluations of multiple causal peak fine-mapping using peak-peak correlations computed across single cells vs. metacells. a)** Average enrichment across recall values of links predicted by multiple causal peak fine-mapped ArchR scores using for 448 links validated by CRISPR. **b)** Average enrichment across recall values of links predicted by multiple causal peak fine-mapped Signac scores for 448 links validated by CRISPR. **c)** Average enrichment across recall values of links predicted by multiple causal peak fine-mapped ArchR scores for 39,194 fine-mapped eSNP–eGene pairs attaining maximum PIP > 0.5 across GTEx tissues. **d)** Average enrichment across recall values of links predicted by multiple causal peak fine-mapped Signac scores for 39,194 fine-mapped eSNP–eGene pairs attaining maximum PIP > 0.5 across GTEx tissues. In primary analyses, SuSiE used peak-peak correlations computed within the susie() function across metacells (for ArchR) or single cells (for

Signac) to perform multiple causal peak fine-mapping. We also assessed fine-mapping using alternatively computed peak-peak correlations (across single cells for ArchR and across metacells for Signac), which we implemented by supplying a peak correlation matrix (across single cells or metacells) to the “R” argument of the susie_rss() function. Fine-mapped scores using alternative peak-peak correlations are noted in parentheses on the x-axis (e.g. “metacell R” or “single-cell R”). Fine-mapping using alternative peak-peak correlations did not outperform fine-mapping using peak-peak correlations from the focal peak-gene linking method. Bars and confidence intervals denote estimates and standard errors, respectively, meta-analyzed across data set-cell type pairs. Green asterisks denote significant underperformance of the focal method vs. the top-performing method (denoted with a black outline) (*: $p < 0.05$, **: $p < 0.01$, ***: $p < 0.001$). Grey horizontal dashed lines mark 1.0x enrichment. MCP, multiple causal peak fine-mapping; MCP + Functional, multiple causal peak functionally informed fine-mapping.

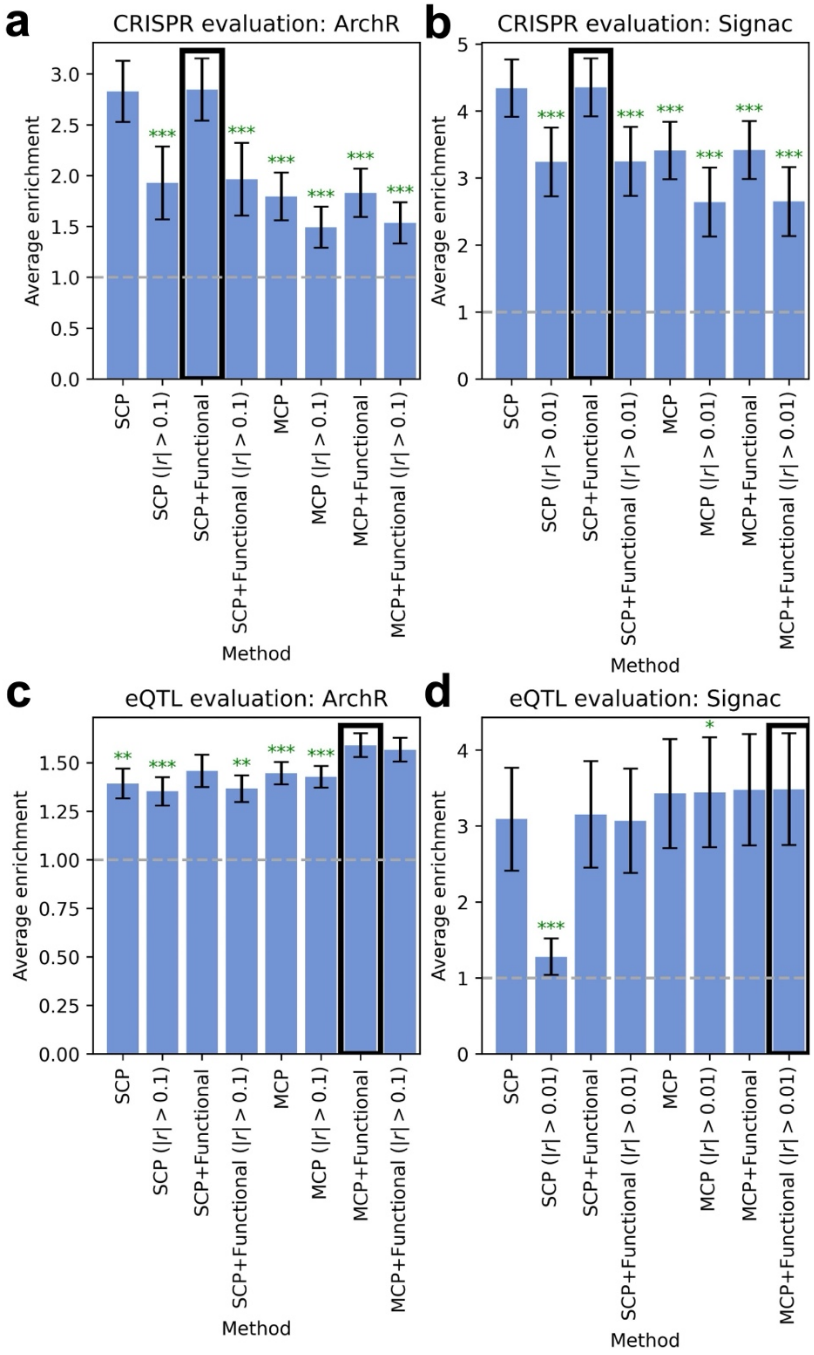


**Supplementary Figure 49. Evaluations of fine-mapping restricting to peak-gene pairs with strong marginal correlations. a)** Average enrichment across recall values of links predicted by ArchR vs. fine-mapped ArchR scores for 448 links validated by CRISPR. **b)** Average enrichment across recall values of links predicted by Signac vs. fine-mapped Signac scores for 448 links validated by CRISPR. **c)** Average enrichment across recall values of links predicted by ArchR vs. fine-mapped ArchR scores for 39,194 fine-mapped eSNP–eGene pairs attaining maximum PIP > 0.5 across GTEx tissues. **d)** Average enrichment across recall values of links predicted by Signac vs. fine-mapped Signac scores for 39,194 fine-mapped eSNP–eGene pairs attaining maximum PIP > 0.5 across GTEx tissues. For all fine-mapping methods labeled "(|*r*| > X)", we restricted the fine-mapping algorithm to candidate peak-gene pairs with marginal peak-gene |*r*|

> X (with X = 0.1 (for ArchR) and X = 0.01 (for Signac). Restricting to peak-gene pairs with strong marginal correlations yielded similar or lower average enrichments on both CRISPR and eQTL evaluation sets vs. fine-mapping without this restriction. Bars and confidence intervals denote estimates and standard errors, respectively, meta-analyzed across data set-cell type pairs. Green asterisks denote significant underperformance of the focal method vs. the top-performing method (denoted with a black outline) (*: p < 0.05, **: p < 0.01, ***: p < 0.001). Grey horizontal dashed lines mark 1.0x enrichment. SCP, single causal peak fine-mapping; SCP + Functional, single causal peak functionally informed fine-mapping; MCP, multiple causal peak fine-mapping; MCP + Functional, multiple causal peak functionally informed fine-mapping.

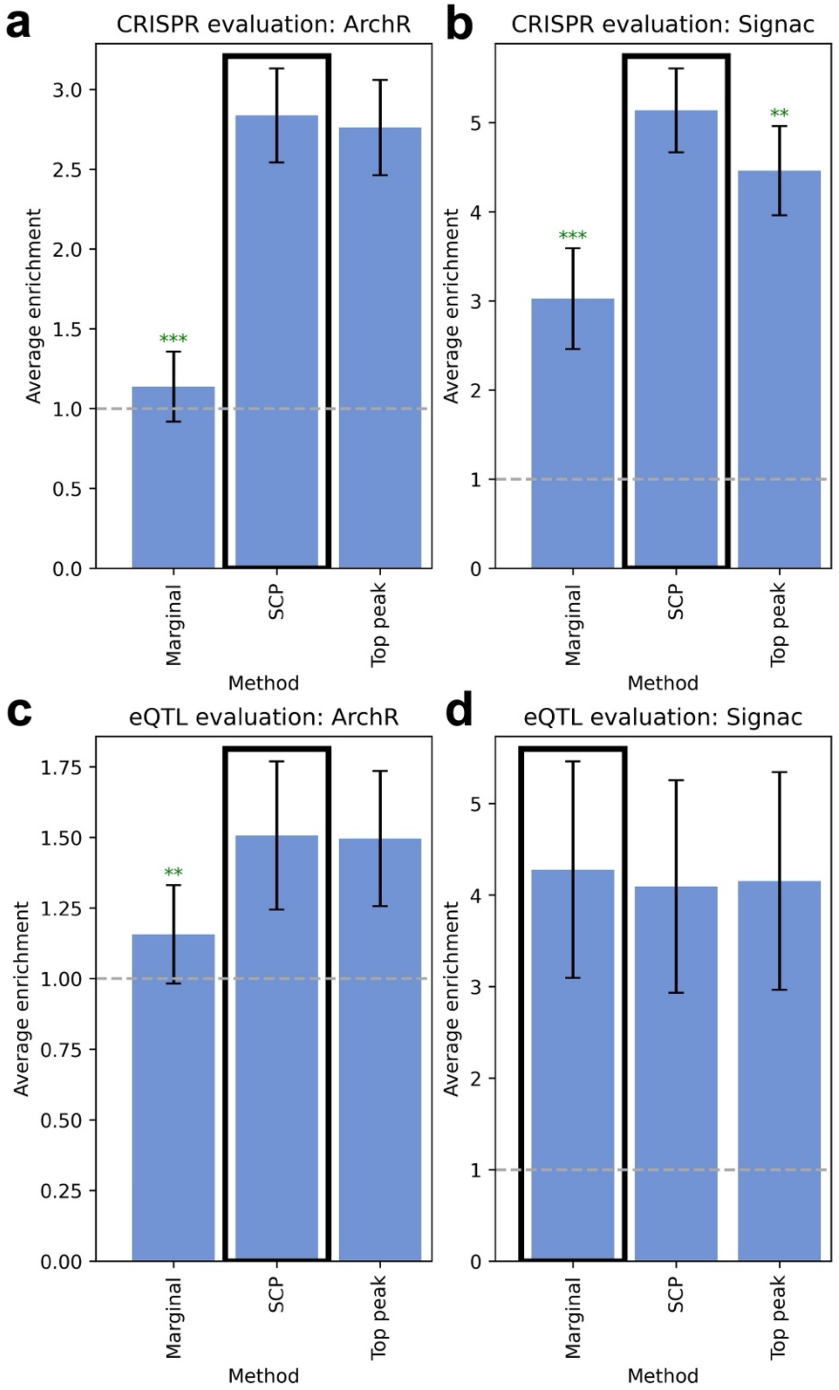


**Supplementary Figure 50. Evaluations of fine-mapping vs. a 'top peak' fine-mapping approach. a)** Average enrichment across recall values of links predicted by ArchR vs. fine-mapped ArchR scores vs. a 'top peak' fine-mapping approach for 448 links validated by CRISPR. **b)** Average enrichment across recall values of links predicted by Signac vs. fine-mapped Signac scores vs. a 'top peak' fine-mapping approach for 448 links validated by CRISPR. **c)** Average enrichment across recall values of links predicted by ArchR vs. fine-mapped ArchR scores vs. a 'top peak' fine-mapping approach for 39,194 fine-mapped eSNP–eGene pairs attaining maximum PIP > 0.5 across GTEx tissues. **d)** Average enrichment across recall values of links predicted by Signac vs. fine-mapped Signac scores vs. a ‘top peak’ fine-mapping approach for 39,194 fine-mapped eSNP–eGene pairs attaining maximum PIP > 0.5 across GTEx tissues. We defined the ‘top peak’ fine-mapping approach as follows: for each gene, we assigned the peak with the highest $r^2$ to the gene a posterior inclusion probability (PIP) of 1, and assigned PIP of 0 to all other candidate peaks. We then defined fine-mapped linking scores as PIP x marginal peak-gene linking score (as for all other fine-mapping approaches). The ‘top peak’ fine-mapping approach significantly underperformed SCP fine-mapped Signac scores on CRISPR evaluation data, but otherwise attained similar average enrichments across shared values of recall. However, ‘top peak’ fine-mapping yielded very low recall, for both the eQTL evaluation (recall 0.0053-0.025 for ‘top-peak’ fine-mapped methods vs. 0.33-0.84 for marginal and SCP fine-mapped methods) and the CRISPR evaluation (recall 0.07-0.2 for ‘top-peak’ fine-mapped methods vs. 0.35-0.98 for marginal and SCP fine-mapped methods) (because the number of peak-gene pairs with PIP > 0 is strictly equal to the number of genes); thus, ‘top-peak’ fine-mapping is a less desirable approach. Bars and confidence intervals denote estimates and standard errors, respectively, meta-analyzed across data set-cell type pairs. Green asterisks denote significant underperformance of the focal method vs. the top-performing method (denoted with a black outline) (*: $p < 0.05$, **: $p < 0.01$, ***: $p < 0.001$). Grey horizontal dashed lines mark 1.0x enrichment. SCP, single causal peak fine-mapping.

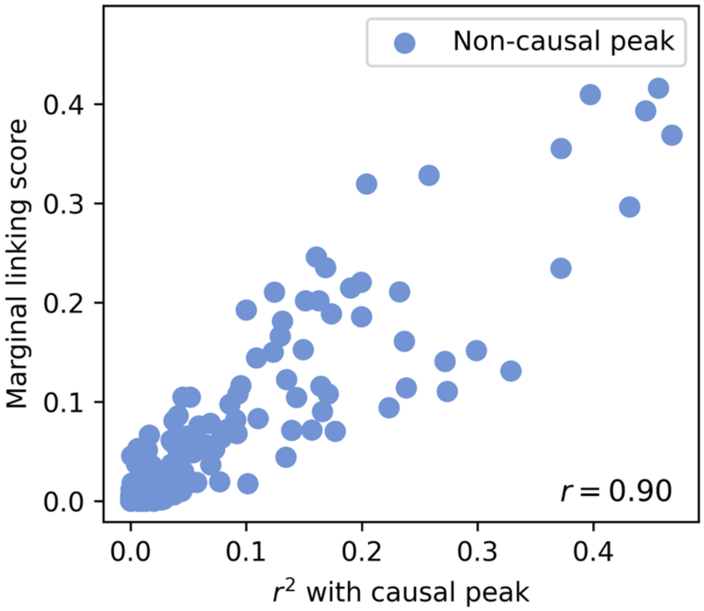


**Supplementary Figure 51. Relationship between marginal linking score and $r^2$ with causal peak across non-causal peaks near *RAP1GAP*.** Relationship between marginal linking score (squared correlation) with *RAP1GAP* and squared correlation with the "causal peak" (peak attaining highest linking score with *RAP1GAP*). Each point represents 1 "non-causal" peak (peak <1Mb from the TSS of *RAP1GAP* that does not attain the highest *RAP1GAP* linking score).

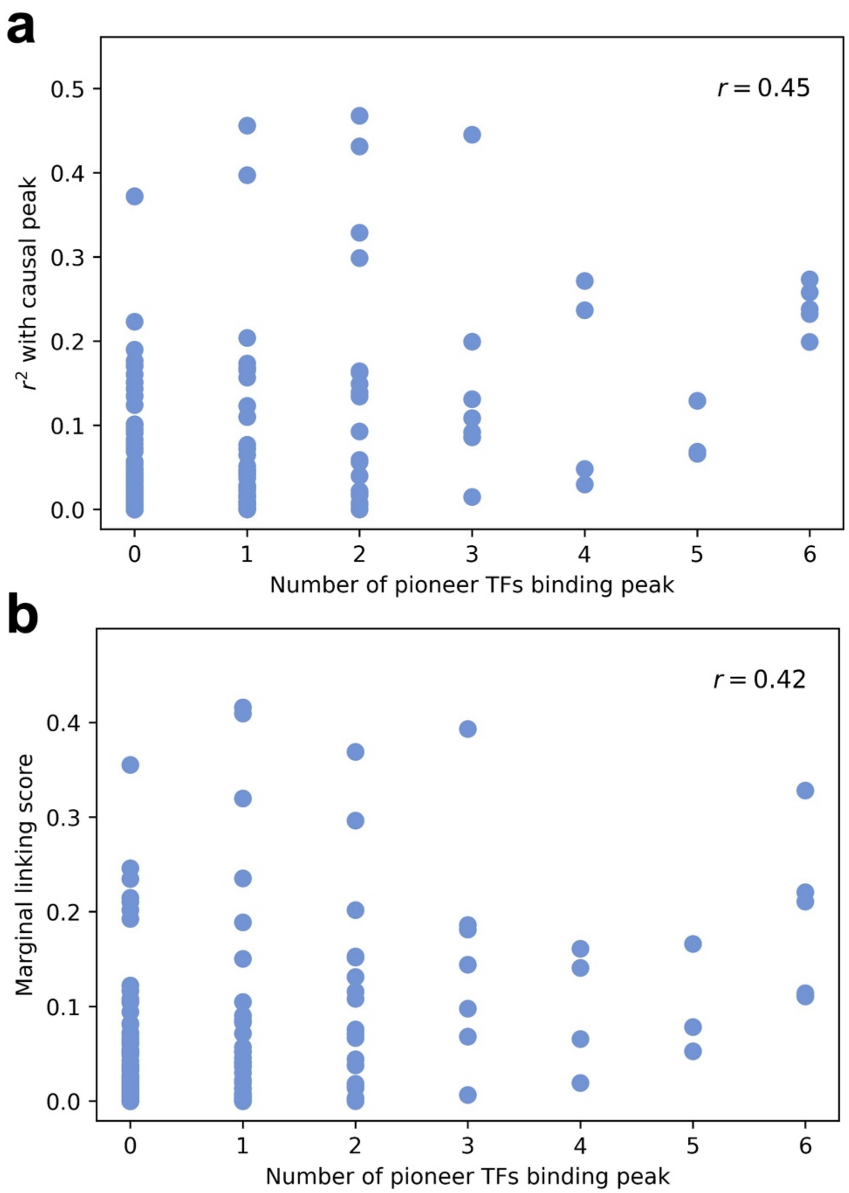


**Supplementary Figure 52. Relationship between pioneer TF activity, peak-peak co-accessibility, and peak-gene co-activity in the *cis* window of *RAP1GAP*. a)** Relationship between squared correlation with the "causal peak" and the number of pioneer TFs binding both the causal peak and the focal peak, across "non-causal" peaks. **b)** Relationship between marginal linking score (squared correlation) with *RAP1GAP* and the number of pioneer TFs binding both the focal peak and the causal peak, across "non-causal" peaks. The "causal peak" (chr1:21687033-21688350) is defined as the peak attaining the highest linking score with *RAP1GAP*, and "non-causal peaks" are defined as peaks <1Mb from the TSS of *RAP1GAP* that do not attain the highest *RAP1GAP* linking score. We measured the number of pioneer TFs that bind each non-causal peak and the causal peak (of 6 total pioneer TFs that bind the causal peak).

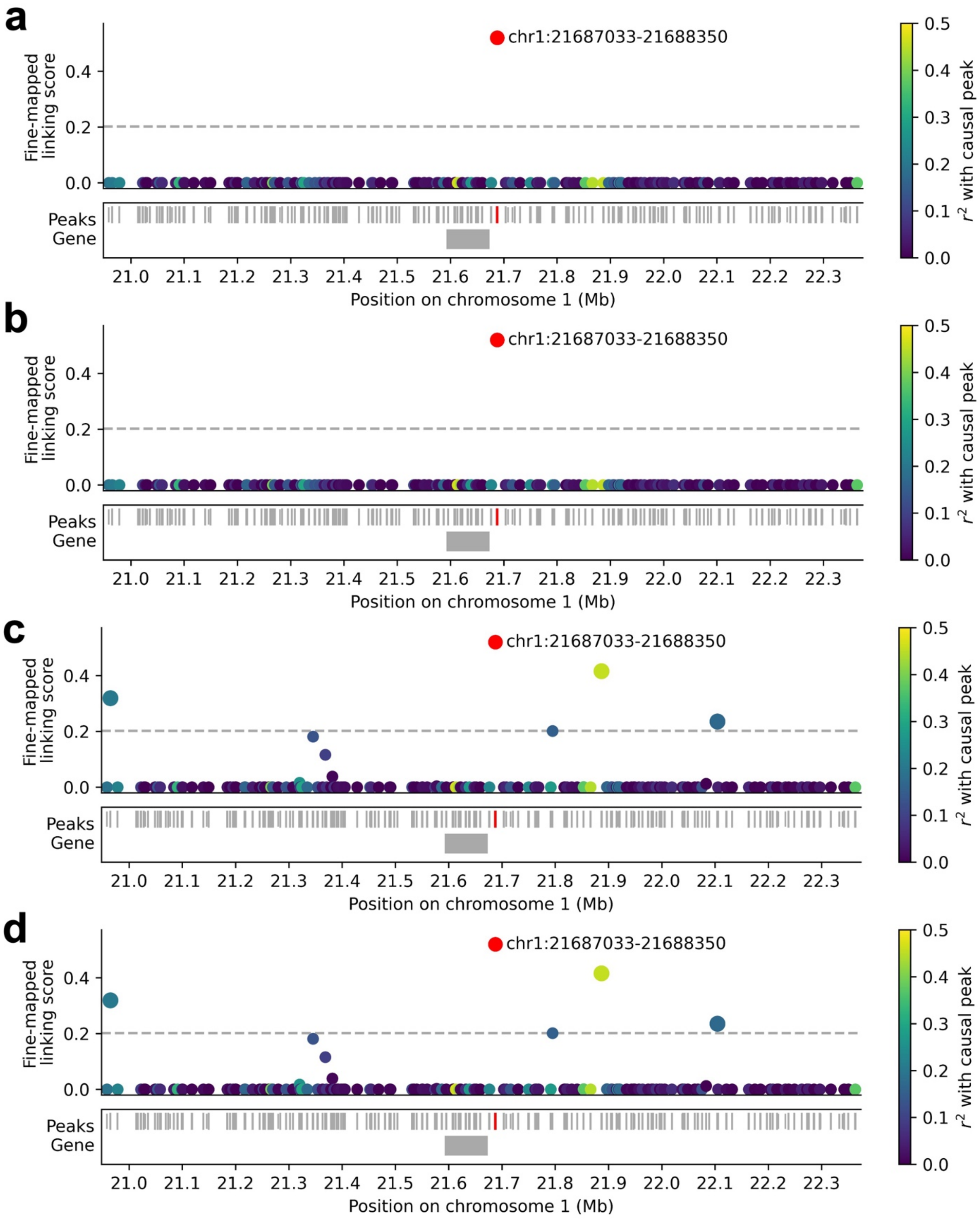


**Supplementary Figure 53. Fine-mapped linking scores for *RAP1GAP*.** Fine-mapped linking scores for each peak <1Mb from the TSS of the gene *RAP1GAP* from **a)** single causal peak fine-mapping, **b)** functionally informed single causal peak fine-mapping, **c)** multiple causal peak fine-mapping, and **d)** functionally informed multiple causal peak fine-mapping. $r^2$ values are reported as dots in the middle of each peak, with the top correlated peak colored in red and other peaks colored based on their squared correlation to the top correlated peak. In each panel, dashed line denotes linking score threshold corresponding to the ArchR linking threshold. Fine-mapping approaches assuming multiple causal peaks linked 3 additional peaks to *RAP1GAP*. None of these peaks harbored fine-mapped GWAS variants; nevertheless, these may be true causal peaks in this or other contexts. Functionally-informed approaches yielded highly similar results to non-functionally-informed approaches.

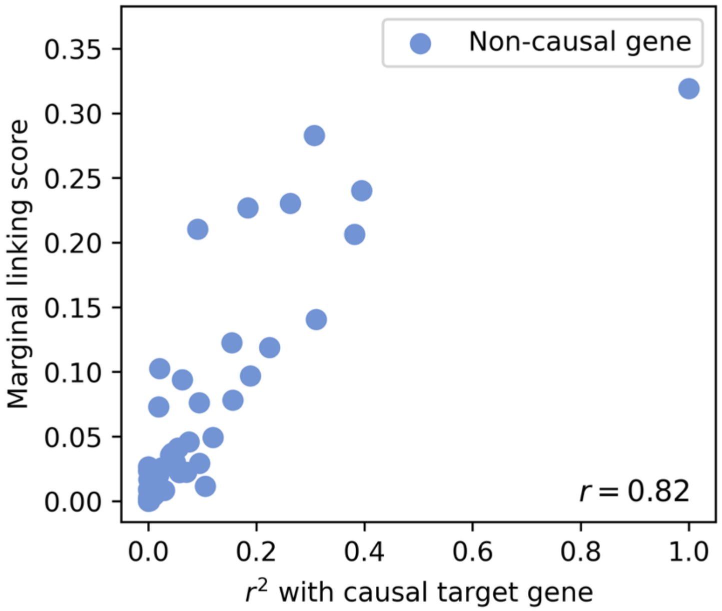


**Supplementary Figure 54. Relationship between marginal linking score and $r^2$ with causal target gene across non-causal genes near the peak chr19:8417808-8418540.** Relationship between marginal linking score (squared correlation) with the focal peak (chr19:8417808-8418540) and squared correlation with the causal target gene (gene attaining highest linking score with the focal peak), across non-causal target genes. Each point represents 1 "non-causal" gene (gene <1Mb from the focal peak that does not attain the highest linking score).

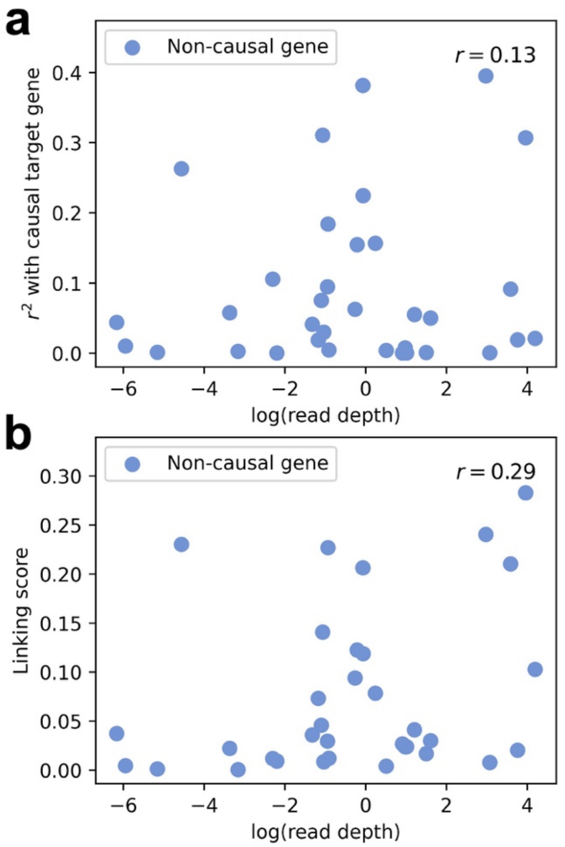


**Supplementary Figure 55. Relationship between gene read depth and gene-gene tagging for the peak chr19:8417808-8418540. a)** Relationship between squared correlation with the causal target gene (*ANGPTL4*) and log(read depth), across non-causal genes. **b)** Relationship between marginal linking score (squared correlation) with the focal peak chr19:8417808-8418540 and log(read depth) across non-causal genes. The causal gene (*ANGPTL4*) is defined as the gene attaining the highest linking score with the focal peak, and non-causal genes are defined as genes <1Mb from the focal peak that do not attain the highest linking score. We also note that *ANGPTL4* had only moderate read depth (with percentile = 0.66 among all genes <1Mb from chr19:8417808-8418540).

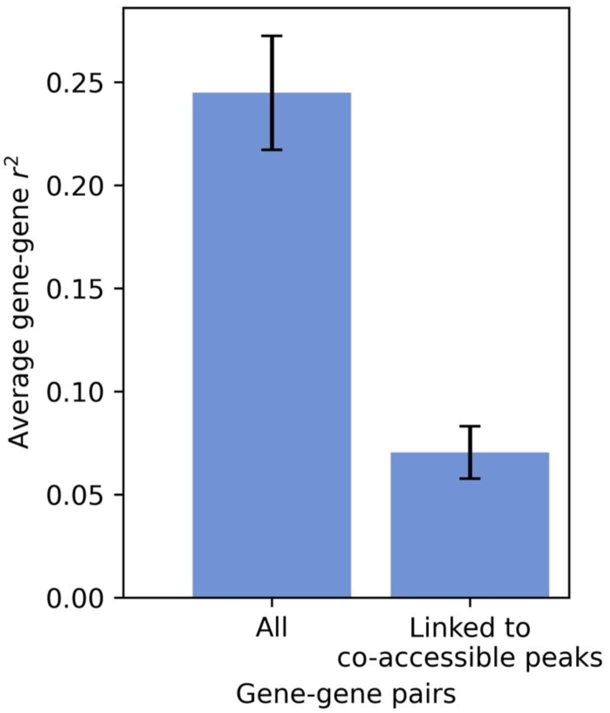


**Supplementary Figure 56. Average $r^2$ with *ANGPTL4* for genes linked to peaks co-accessible with peak chr19:8417808-8418540 vs. all *cis* genes.** We identified 51 peaks linked to any gene <1Mb from the peak chr19:8417808-8418540 (besides *ANGPTL4*) (defining "linked" as ArchR linking score ($r^2$) > 0.2025). From these peak-gene links, we identified 7 genes linked to peaks highly co-accessible with chr19:8417808-8418540 (defining "highly co-accessible" as peak-peak $r^2$ > 0.2025). We then computed the average $r^2$ between *ANGPTL4* and each of these genes (right) vs. the average $r^2$ between *ANGPTL4* and each gene <1Mb from *ANGPTL4*. Average $r^2$ with *ANGPTL4* for genes linked to peaks highly co-accessible with chr19:8417808-8418540 was substantially higher ($r^2$ = 0.24) than for all genes ($r^2$ = 0.070). Confidence intervals denote standard errors (from jackknifing across individual genes).

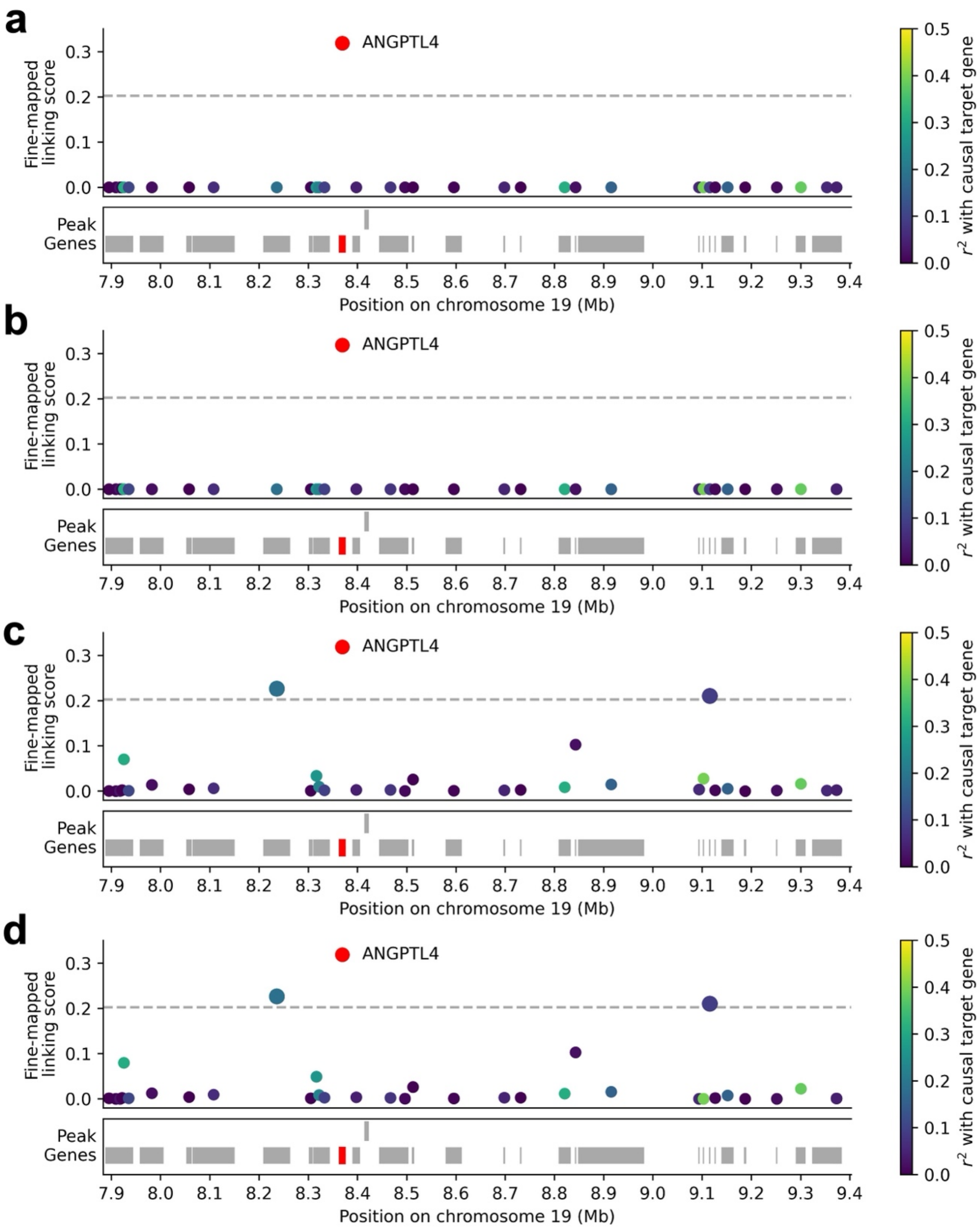


**Supplementary Figure 57. Gene-level fine-mapped linking scores for the peak chr19:8417808-8418540.** Gene-level fine-mapped linking scores for each gene <1Mb from the peak spanning chr19:8417808-8418540 from **a)** single causal gene fine-mapping, **b)** functionally informed single causal gene fine-mapping, **c)** multiple causal gene fine-mapping, and **d)** functionally informed multiple causal gene fine-mapping. $r^2$ values are reported as dots in the middle of each gene, with the top correlated gene colored in red and other peaks colored based on their squared correlation to the top correlated gene. In each panel, dashed line denotes linking score threshold corresponding to the ArchR linking threshold.
